# Efficacy and safety of biologic, biosimilars and targeted synthetic DMARDs in moderate-to-severe rheumatoid arthritis with inadequate response to methotrexate: a systematic review and network meta-analysis

**DOI:** 10.1101/2023.01.20.23284852

**Authors:** Nuttakarn Budtarad, Juthamas Prawjaeng, Pattara Leelahavarong, Songyot Pilasant, Chonticha Chanjam, Pongthorn Narongroeknawin, Tasanee Kitumnuaypong, Wanruchada Katchamart

**Affiliations:** Health Intervention and Technology Assessment Program (HITAP), Ministry of Public Health, Thailand Siriraj Hospital, Faculty of Medicine Siriraj Hospital, Mahidol University, Bangkok, Thailand; Health Intervention and Technology Assessment Program (HITAP), Ministry of Public Health, Thailand Siriraj Health Policy, Faculty of Medicine Siriraj Hospital, Mahidol University, Bangkok, Thailand; Health Intervention and Technology Assessment Program (HITAP), Ministry of Public Health, Thailand United States Agency for International Development (USAID) Regional Development Mission for Asia, Bangkok, Thailand; Health Intervention and Technology Assessment Program (HITAP), Ministry of Public Health, Thailand Puey Ungphakorn School of Development Studies, Thammasat University, 99 Moo 18 Paholyothin Road, Klong Nueng, Klong Luang, Pathumthani, 12121, Thailand; Division of Rheumatology, Department of Internal Medicine, Phramongkutklao Hospital and College of Medicine, Bangkok, Thailand; Rheumatology Unit, Department of Medicine, Rajavithi Hospital, Ministry of Public Health, Bangkok, Thailand; Division of Rheumatology, Department of Medicine, Faculty of Medicine Siriraj Hospital, Mahidol University, Bangkok, Thailand

**Author notes:** (Corresponding author)* **Corresponding author:** Dr Wanruchada Katchamart, MD, MSc (Clin Epi), Division of Rheumatology, Department of Medicine, Faculty of Medicine Siriraj Hospital, Mahidol University, Bangkok, Thailand. **Contributors:** PL, JP, NB and WK designed the study. PN, TK, and WK provided clinical opinions. PL, SP, CC, JP, and NB screened studies for inclusion, extracted data and performed quality assessment. NB and JP analysed the data and performed statistical analysis. NB, JP, PL, and WK interpreted the data. NB, JP and PL drafted the first version of the manuscript. All authors approved the final version of the manuscript. WK and NB are the guarantors. **Ethical approval:** Not required.

**Keywords:** Antirheumatic Agents, Arthritis, Rheumatoid with inadequate responses to conventional synthetic DMARDs, serious adverse events, network meta-analysis

## Abstract

**Objective:** To assess the comparative efficacy and safety of approved biologic disease-modifying antirheumatic drugs (bDMARDs), biosimilars, and targeted synthetic disease-modifying antirheumatic drugs (tsDMARDs) for patients with rheumatoid arthritis (RA) who had inadequate responses to methotrexate (MTX).

**Results:** 53 eligible studies were identified and 44 studies were included in a network meta-analysis. Using Surface Under the Cumulative Ranking Curve (SUCRA), tofacitinib (10 mg bid) with MTX [Relative risk (RR) 95% confidence interval (CI) 4.65 (2.98-7.27)] and tofacitinib (10 mg bid) [RR (95%CI)1.96 (1.27-3.03)] were ranked highest among tsDMARDs for increasing remission rate at 24-26 weeks and 48-52 weeks, respectively. For bDMARDs, tocilizumab (8 mg/kg) with MTX was ranked with highest treatment effect for remission at both 24-26 and 48-52 weeks [RR (95%CI) 3.06 (2.27-4.12); RR (95%CI) 2.52 (1.94-3.28)]. For safety, baricitinib (4 mg) and tofacitinib (5 mg bid) with MTX likely showed an increased risk of HZ with statistical significance [for baricitinib, RR (95%CI) 3.52 (1.38-9.02) at 24-26 weeks, and RR (95%CI) 4.20 (1.22-14.48) at 48-52 weeks, and for tofacitinib, RR (95%CI) 5.38 (1.00-28.91) at 48-52 weeks]. No statistically significant safety concerns for serious infection, tuberculosis (TB), cancer, and cardiovascular (CV) events were identified.

**Conclusions:** For RA patients who failed MTX, bDMARDs, biosimilars, and tsDMARDs monotherapy and combination therapy with MTX provided better treatment outcomes than MTX monotherapy with modest safety concerns within 24-52 weeks. A scarcity of longer-term effects and post-market surveillance necessitates further analyses using long-term patient-level data to improve the medication profile.

**Rheumatology key messages:**

- For RA patients who failed MTX and other conventional DMARDs, different types of DMARDs are available.
- At dose- and time point-specific levels, tofacitinib (10 mg bd) showed the highest probability to be the most effective in achieving remission at 24-26 weeks.
- An increased risk of herpes zoster was found for baricitinib (4 mg) and tofacitinib (5 mg bid) with MTX.

## Introduction

Rheumatoid arthritis (RA) is the most common idiopathic inflammatory arthritis (1). RA can lead to pain, functional disability, reduced quality of life, and premature mortality (2). The current recommendations for management of RA include non-steroidal anti-inflammatory drugs or corticosteroids for pain relief, as well as conventional synthetic disease-modifying antirheumatic drugs (csDMARDs), biologic DMARDs (bDMARDs), or targeted synthetic DMARDs (tsDMARDs) to target the pathophysiological basis of the disease (3).

Treat-to-target aiming at remission is the mainstay strategy that leads to favourable outcomes (4). Methotrexate (MTX) is recommended as the first line csDMARDs (5–7). However, in patients who still have moderate to high disease activity despite maximum tolerated doses of MTX, a combination of csDMARDs, a bDMARD, or a tsDMARD, is conditionally recommended over triple therapy (i.e., MTX with the addition of sulfasalazine and hydroxychloroquine) (7). MTX in combination with a bDMARD or tsDMARD also has greater persistence when compared to triple therapy (8, 9). This recommendation is based on very low-certainty evidence and no difference was found in long-term outcomes across both treatment strategies (10–12). When the target of therapy is not achieved, switching between different classes of bDMARDs or tsDMARDs is conditionally recommended over switching to the same class for patients taking a bDMARD or tsDMARD. Very low-certainty evidence also suggests greater improvement in disease activity and drug survival. Although safety data indicating an increased risk of serious cardiovascular (CV) events and cancer (13, 14) has been reported by the USFDA, there is no current recommendation on selecting between bDMARDs and tsDMARDs for RA patients.

There are several bDMARDs, biosimilars and tsDMARDs available in the market. However, there are few head-to-head direct comparisons, hence indirect comparisons are a necessity for clinical practice and policy decisions (15). This systematic review and network meta-analysis was conducted to compare and rank the efficacy and safety of 12 patented bDMARDs and tsDMARDs, divided into five main groups: 1) anti-TNF-α i.e., adalimumab, certolizumab pegol, etanercept, golimumab and infliximab; 2) anti-IL6 i.e., sarilumab and tocilizumab; 3) anti-CD20 i.e., rituximab; 4) anti-cytotoxic T-lymphocyte-associated antigen (CTLA)-4 i.e., abatacept, and 5) JAK inhibitor i.e., baricitinib, filgotinib, and tofacitinib. In addition, nine biosimilars of these originators were included in this study: 1) two adalimumab biosimilars i.e., ABP 501 and SB5; 2) three etanercept biosimilars i.e., HD203, LBEC0101 and SB4; 3) three infliximab biosimilar i.e., CT-P13, PF-06438179/GP1111 and SB2; and 4) rituximab biosimilar i.e., CT-P10. All selected bDMARDs and tsDMARDs in combination with MTX were investigated.

By comparing desirable clinical outcomes of bDMARDs, biosimilars and tsDMARDs, this study focuses on remission using the disease activity score 28-erythrocyte sedimentation (DAS28-ESR) and well-known serious adverse events (SAEs), including serious infection, tuberculosis (TB), herpes zoster (HZ), thromboembolism, cancer, and cardiovascular (CV) events. This study aims to compare clinical outcomes and safety of bDMARDs and tsDMARDs and provide a comprehensive evidence base to enable clinicians to select the most advantageous medicines for RA patients who have inadequately responded to MTX. This review was initially conducted to generate data for input into a model-based economic evaluation of bDMARDs and tsDMARDs. The results derived from all syntheses (i.e., network meta-analysis and the economic evaluation) were used to inform the policy decision-making process of the Subcommittee of Development of the Thai National List of Essential Medicine (16).

## Methods

### Search strategy

A systematic literature search was conducted using the adapted search strategy from a previous study (17) that combined MeSH terms, keywords, and text words for “rheumatoid arthritis” including comparative medicines “methotrexate, abatacept, adalimumab, certolizumab pegol, etanercept, golimumab, infliximab, sarilumab, tocilizumab, rituximab, baricitinib, filgotinib, tofacitinib” and “randomised controlled trials” to search in MEDLINE, EMBASE, and the Cochrane Library from the inception date to November 30, 2021, without language restrictions. We also reviewed the reference lists of all eligible studies to ensure no relevant trials were missed (Supplementary 1).

### Study selection

We included published studies with the following characteristics: 1) randomized controlled trials (RCTs) of at least 24 weeks’ duration in adults (age ≥18 years) with RA who had inadequate responses to MTX; 2) comparisons between either bDMARDs or tsDMARDs with a common comparator for a network meta-analysis and biological medicines monotherapy and combined with MTX (Supplementary 2); and 3) investigations of at least one of relevant outcome i.e., the proportion of remission (DAS28-ESR<2.6) and serious adverse events (SAEs) including serious infection, TB, HZ, thromboembolism, cancer, and CV events. Post-hoc analyses of RCT and extension trials with treatment switching trials prior to week 24 were excluded. Four review authors (NB, JP, SP, CC) independently screened titles or abstracts and full-text articles to identify published RCTs that matched the predefined inclusion criteria. Disagreements were resolved by consensus or discussion with the fifth review author (PL).

### Data extraction

Four review authors worked in pairs (NB and CC; JP and SP) to independently extract the data from eligible studies into an extraction form. The following information was extracted: 1) publication information (i.e., author, published year, trial name); 2) compared interventions (i.e., generic name, dosage regimen, route of administration); 3) total number and participant demographics of each intervention (i.e., mean age, percentage of females and baseline DAS28-ESR); 4) measured outcomes (i.e., the proportion of patients who achieved remission (DAS28-ESR<2.6) at week 24-26 or 48-52; and 5) number of patients who experienced SAEs at week 24-26 or 48-52. Data presented only in graphical format were digitally estimated and numbers were extracted. Discrepancies in the extracted data including missing data were resolved by discussion with the fifth review author (PL).

Medications and dosages were defined as follows: abatacept (125 mg) was defined as abatacept 125 mg subcutaneously injection (SC) once weekly, abatacept (10 mg/kg) was defined as abatacept 10 mg/kg intravenous infusion (IV) every 4 weeks, adalimumab (20 or 40 mg) was defined as adalimumab 20 or 40 mg SC injection every other week, certolizumab pegol (400 mg) was defined as certolizumab pegol 400 mg SC every 4 weeks, etanercept (25 or 50 mg) was defined as etanercept 25 or 50 mg SC once weekly, golimumab (50 or 100 mg) was defined as golimumab 50 or 100 mg SC every 4 weeks, infliximab (3 mg/kg) was defined as infliximab 3 mg/kg IV every 8 weeks, sarilumab (200 mg) was defined as sailumab 200 mg SC every 2 week, tocilizumab (4 or 8 mg/kg) was defined as tocilizumab 4 or 8 mg/kg IV every 4 weeks, rituximab (500 or 1,000 mg) was defined as rituximab 500 or 1,000 mg IV 2 courses (15 days apart) every 24 weeks, baricitinib (2 or 4 mg) was defined as baricitinib 2 or 4 mg orally once daily, filgotinib (200 mg) was defined as filgotinib 200 mg orally once daily, and tofacitinib (5 or 10 mg) was defined as tofacitinib 5 or 10 mg orally twice daily. A summary table of all medications included can be found in Supplementary 2.

### Risk of bias assessment

Version 2 of the Cochrane risk-of-bias tool for randomized trials (RoB 2) was used to assess the methodological quality of included studies by four independent review authors (NB, JP, SP, CC), with disagreements resolved through discussion (18, 19). The RoB2 assessments were performed through the Excel spreadsheets developed by the RoB2 development group (20). The following methodological domains were assessed: randomization process, deviations from the intended interventions, missing outcome data, measurement of the outcomes, and selection of the reported results. The response options comprised yes, probably yes, probably no, no, and no information. A risk-of-bias judgement arising from each domain was assigned by an algorithm to grade as having a “low” or “high” risk of bias, or “some concerns”. An overall risk-of-bias judgement of included studies was graded according to the judgement across five domains (Supplementary 3).

### Data synthesis and statistical approach

A network meta-analysis corresponded to a generalised linear model and was conducted from a set of data that can be linked by a potential common comparator (i.e., MTX) with a sufficient number of patients with DAS28-ESR or predefined SAEs of at least 24-weeks duration. Our network meta-analysis was conducted according to a frequentist approach using STATA (StataCorp. 2019. *Stata Statistical Software*: *Release 16*. College Station, TX: StataCorp LLC) program command (21). Results of our analyses are presented as relative risk (RR) with 95% confidence intervals (CI) for each regimen versus MTX monotherapy. Network plot was generated to demonstrate the relationship for each medication for each outcome. A hierarchy of medications was generated using the Surface Under the Cumulative Ranking Curve (SUCRA), ranging from 1-100%, according to their likelihood to be ranked in a particular order. A higher SUCRA value indicates a higher likelihood of such intervention being ranked at a higher order in the hierarchy and vice versa (22). SUCRA accounts for both the location and the variance of relative risk of all regimens (23).

### Similarity, homogeneity & consistency assessment

Baseline characteristics of the study population, including the severity of RA, age, prior use of csDMARDs, and study duration, were assessed qualitatively for similarity. The similarity of trial design and characteristics of each included study were also assessed using the same approach. Our model assumes heterogeneity of data and the results were analysed using a random-effects model as a default. If no heterogeneity was found, a fixed-effects model was employed (24). A global test was used to assess consistency between indirect and direct data sets for each outcome. Details on consistency (significance at the 0.05 level) are available in Supplement 4.

### Public and patient involvement

No patients were involved in setting the research question or the outcome measures, nor were they involved in the design or implementation of the study. Two stakeholder meetings comprising clinicians, public persons, pharmaceutical company representatives, and methodologists were held to verify and establish consensus on the study results. The results of this study were reviewed by external reviewers and incorporated into the economic evaluation report submitted to the Health Economic Working Group under the Subcommittee of Development of the Thai National List of Essential Medicine (16).

## Results

### Search results and description of eligible studies

From 2,889 individual records, we included 53 studies meeting our inclusion criteria (n = 26,113), and 44 studies were included in our network meta-analysis after assessing for network connectivity (Figure 1). We included 22 medications with 39 regimens (Supplementary 2). The mean age of subjects was 51.89 ±2.91 years and 80.52 % were female (data not shown). We included 41 studies for efficacy outcomes and 33 studies for safety outcomes (Table 1). Patients in all studies were diagnosed using either the 1987 American College of Rheumatology (ACR) (25) or the 2010 ACR/European League Against Rheumatism (EULAR) criteria (26). The studies were initiated between 2000 and 2019, and 47 of 53 studies were double-blinded (Table 1). Most studies reported remission and safety outcomes at 24-26 weeks.

**Table 1.**
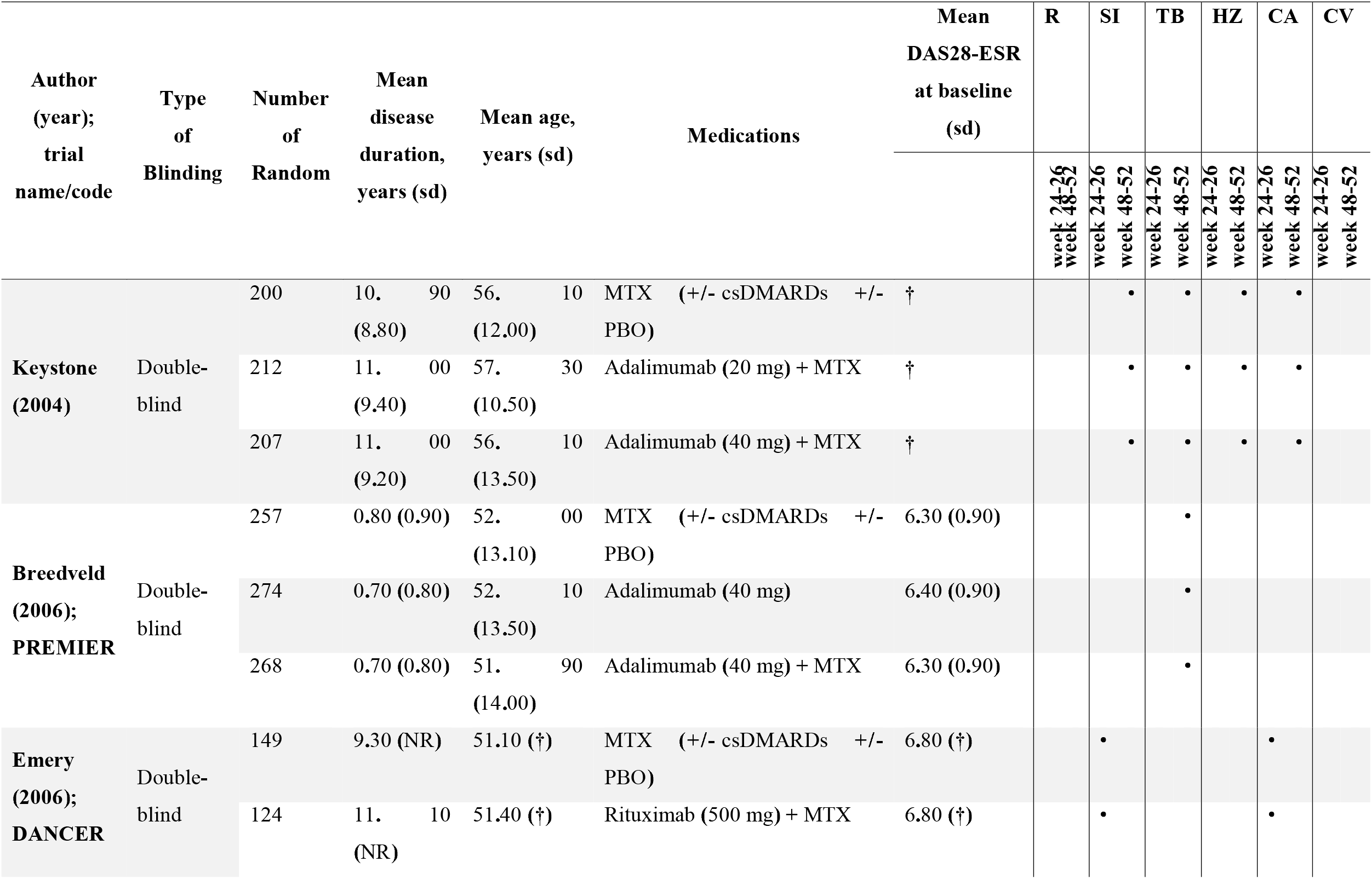
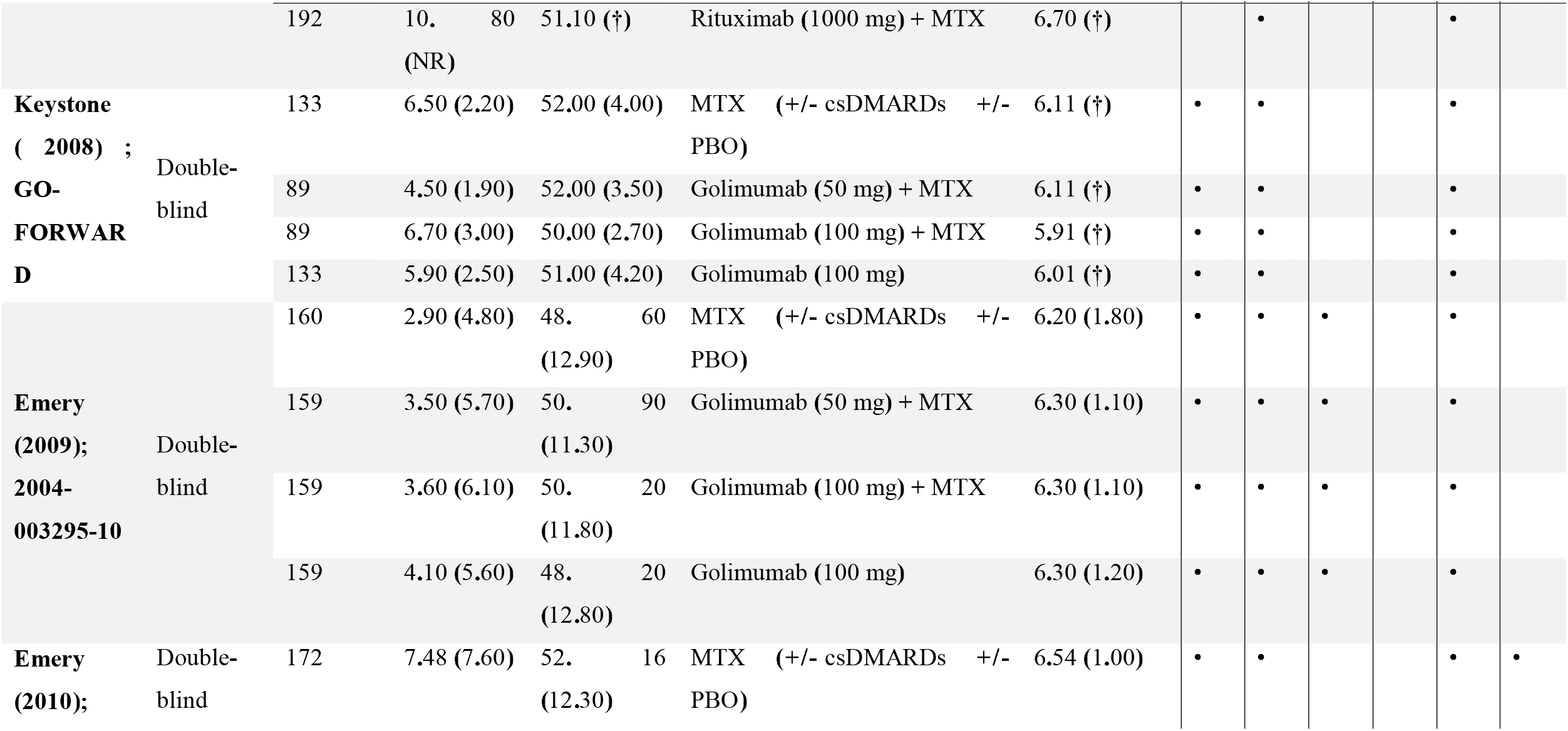
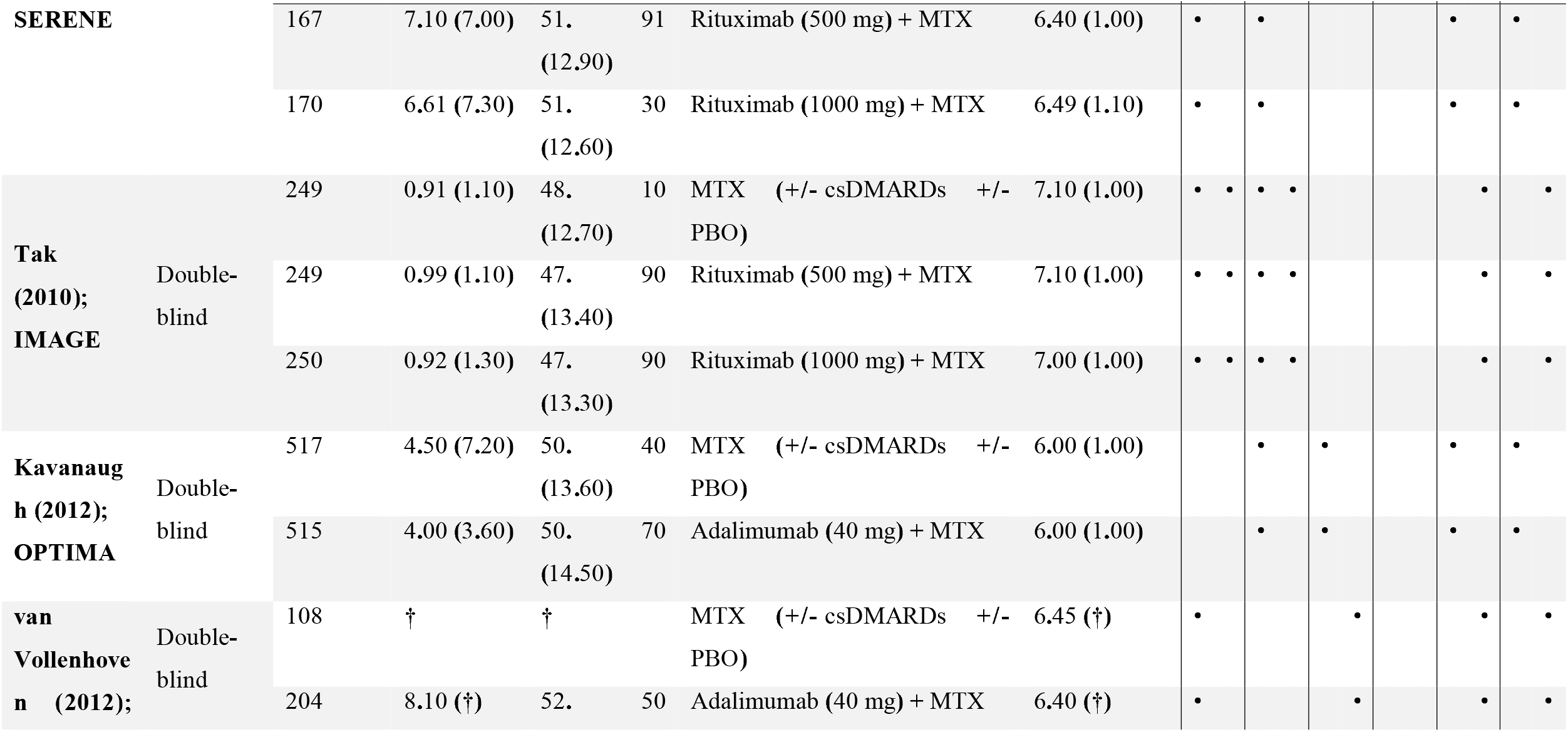
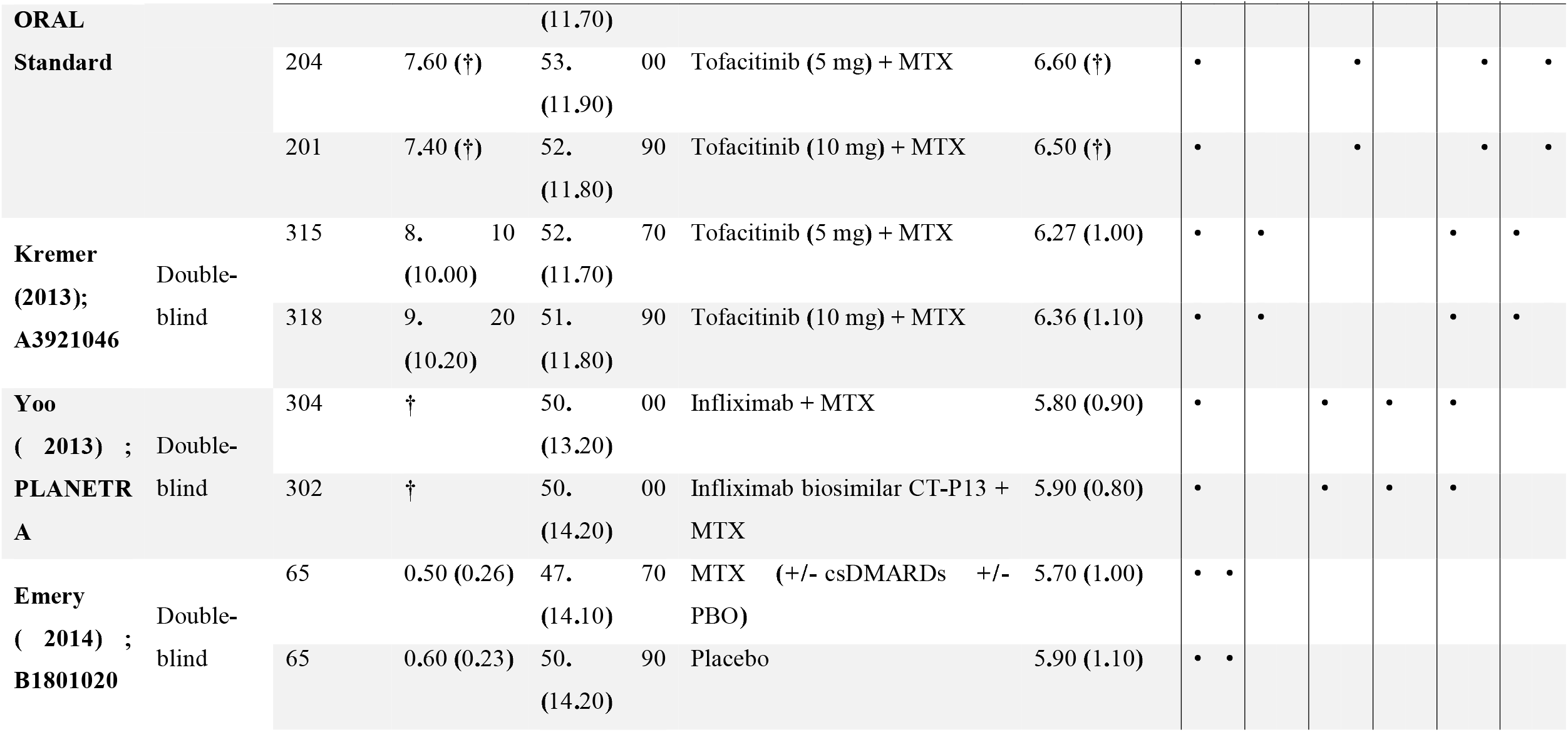
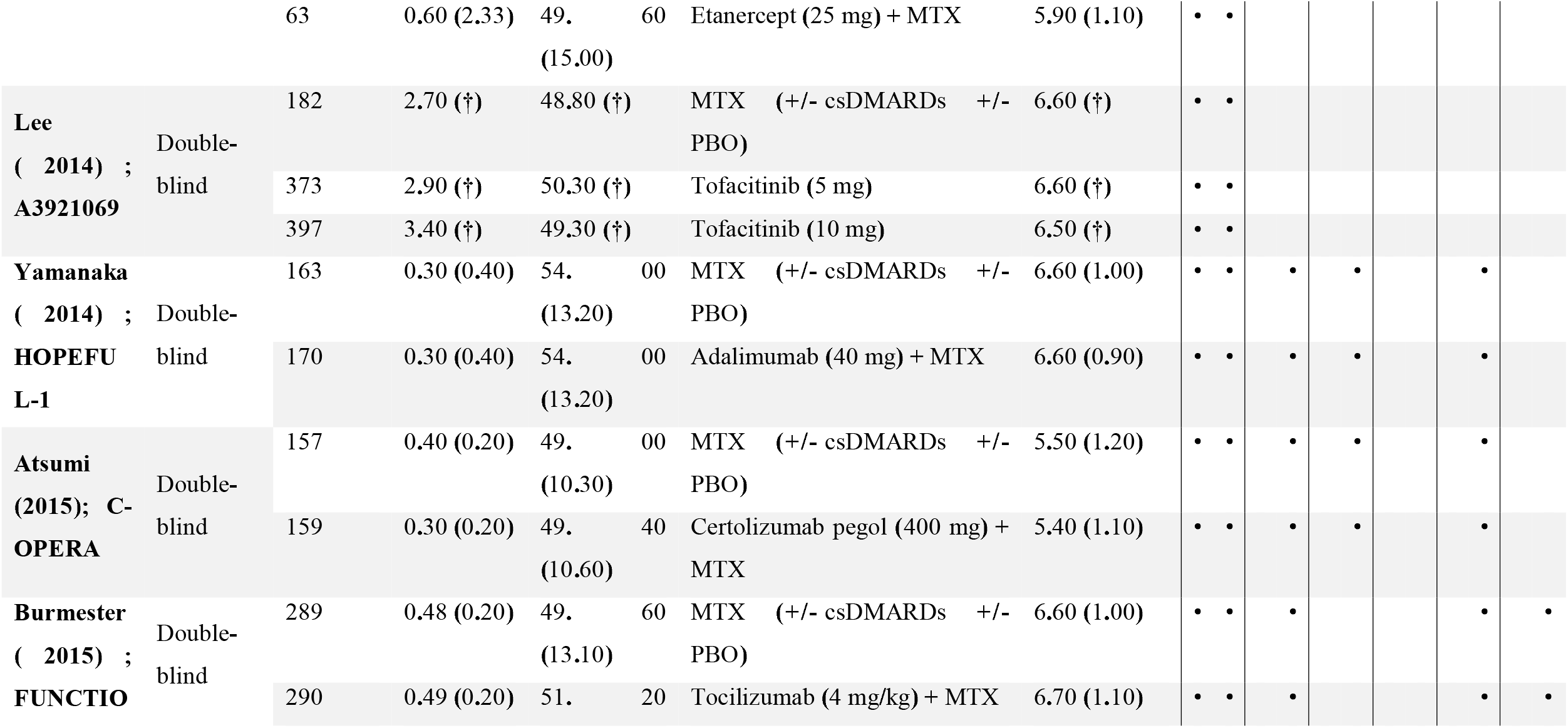
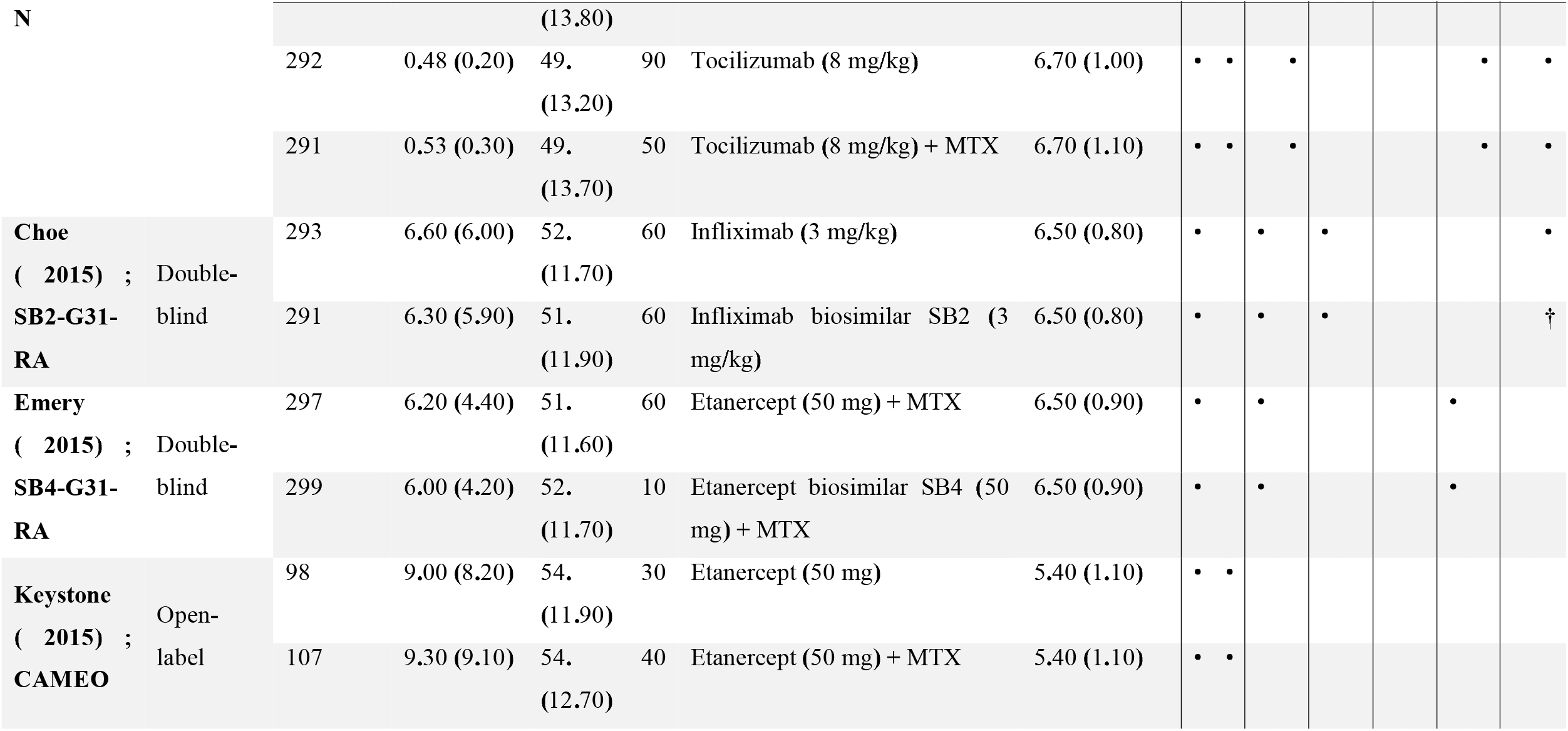
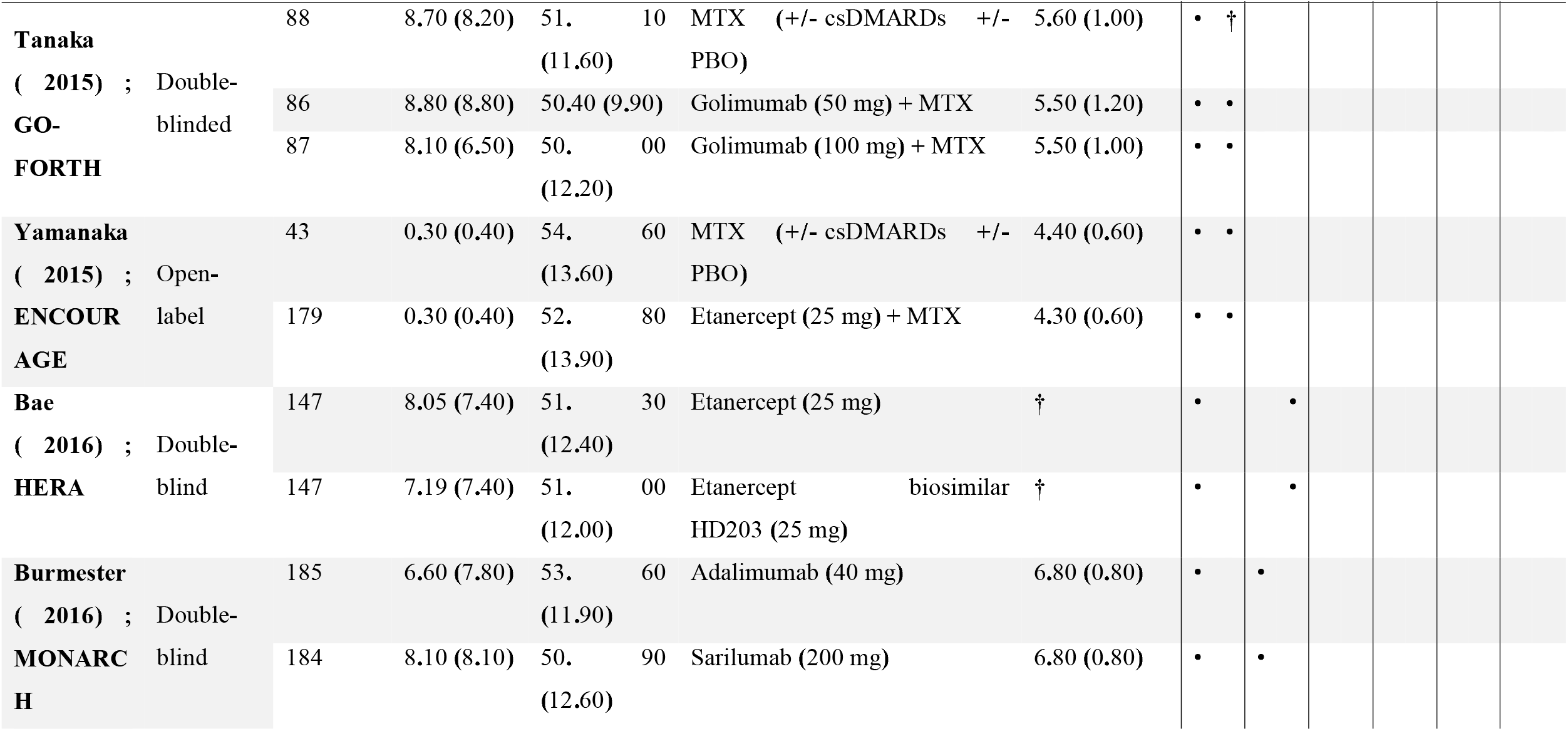
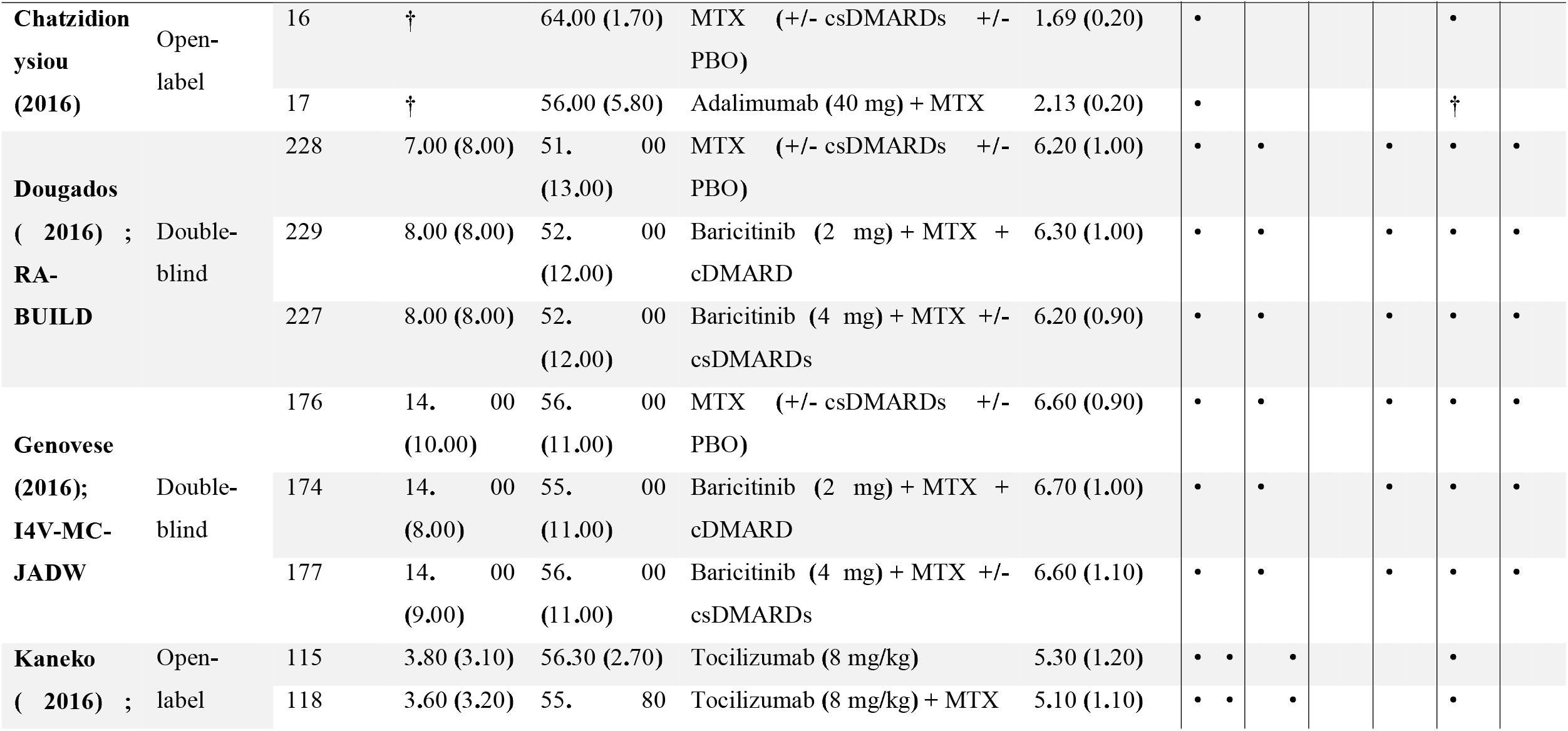
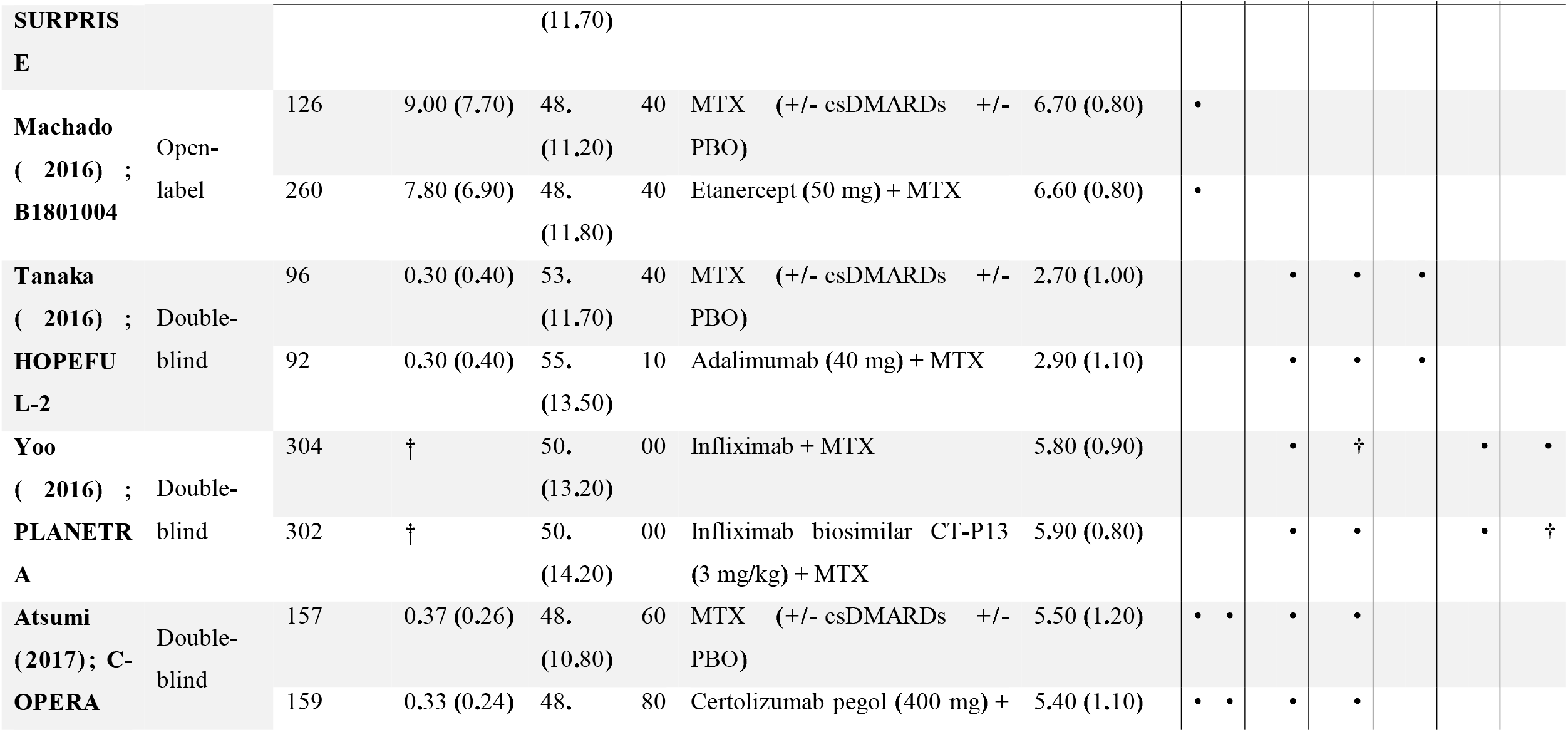
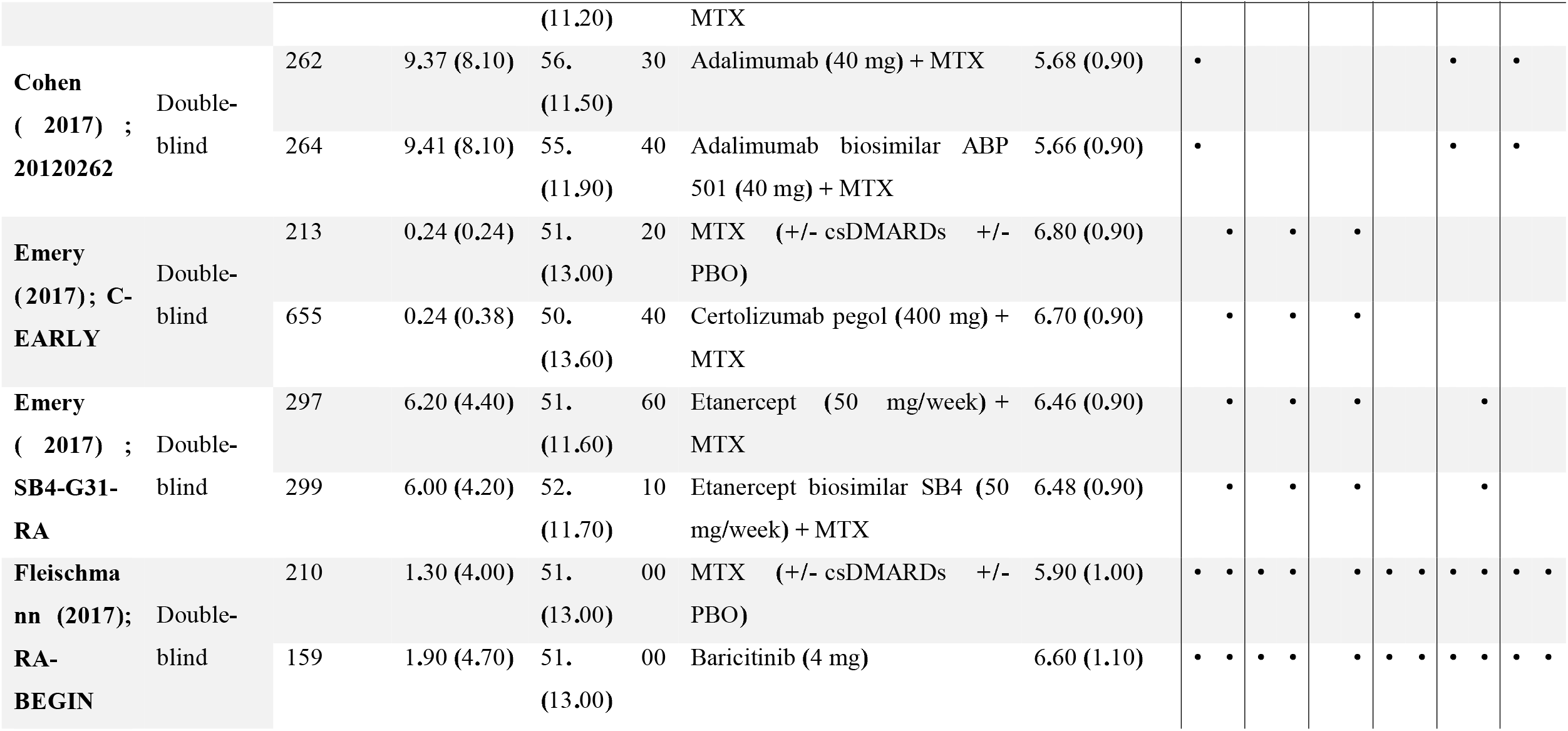
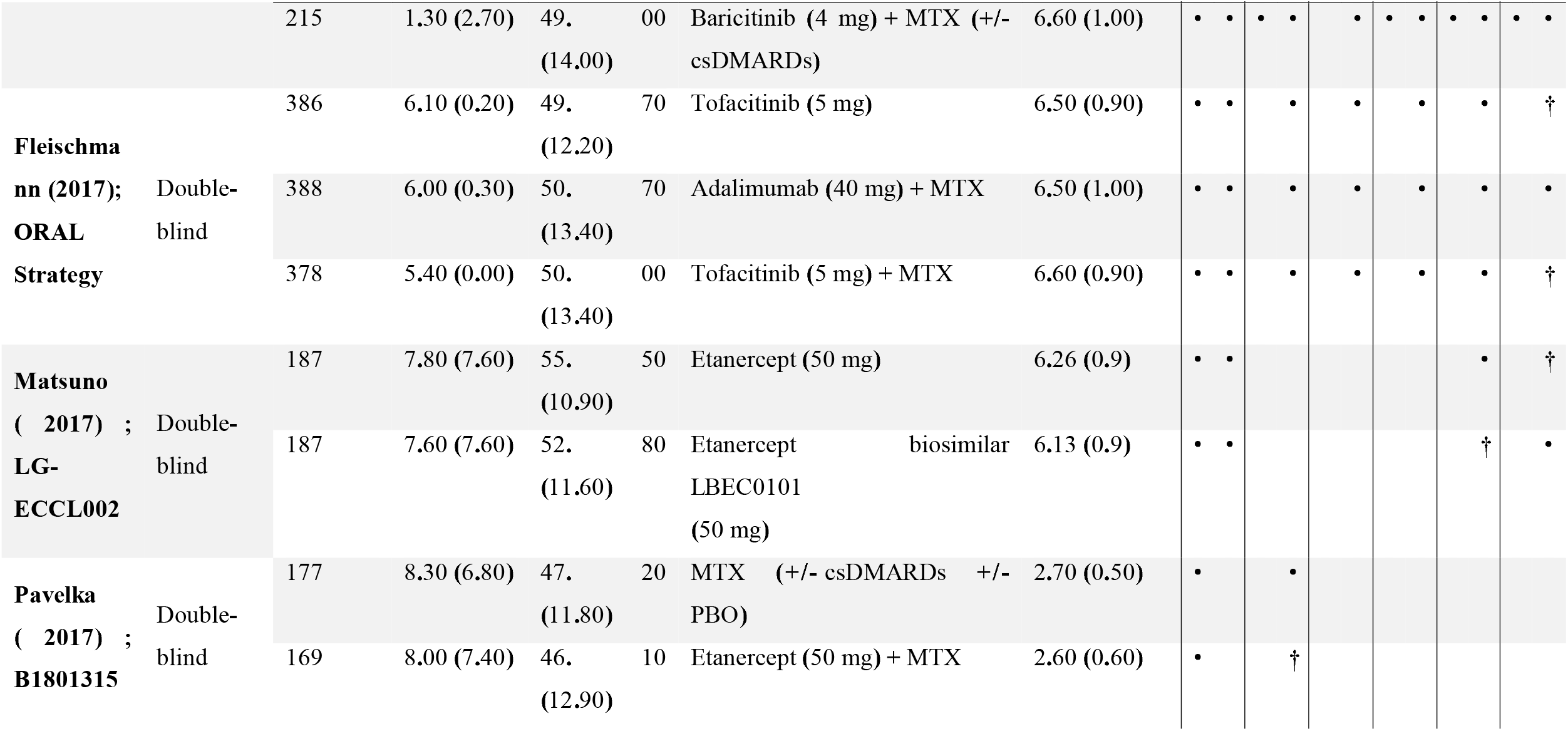
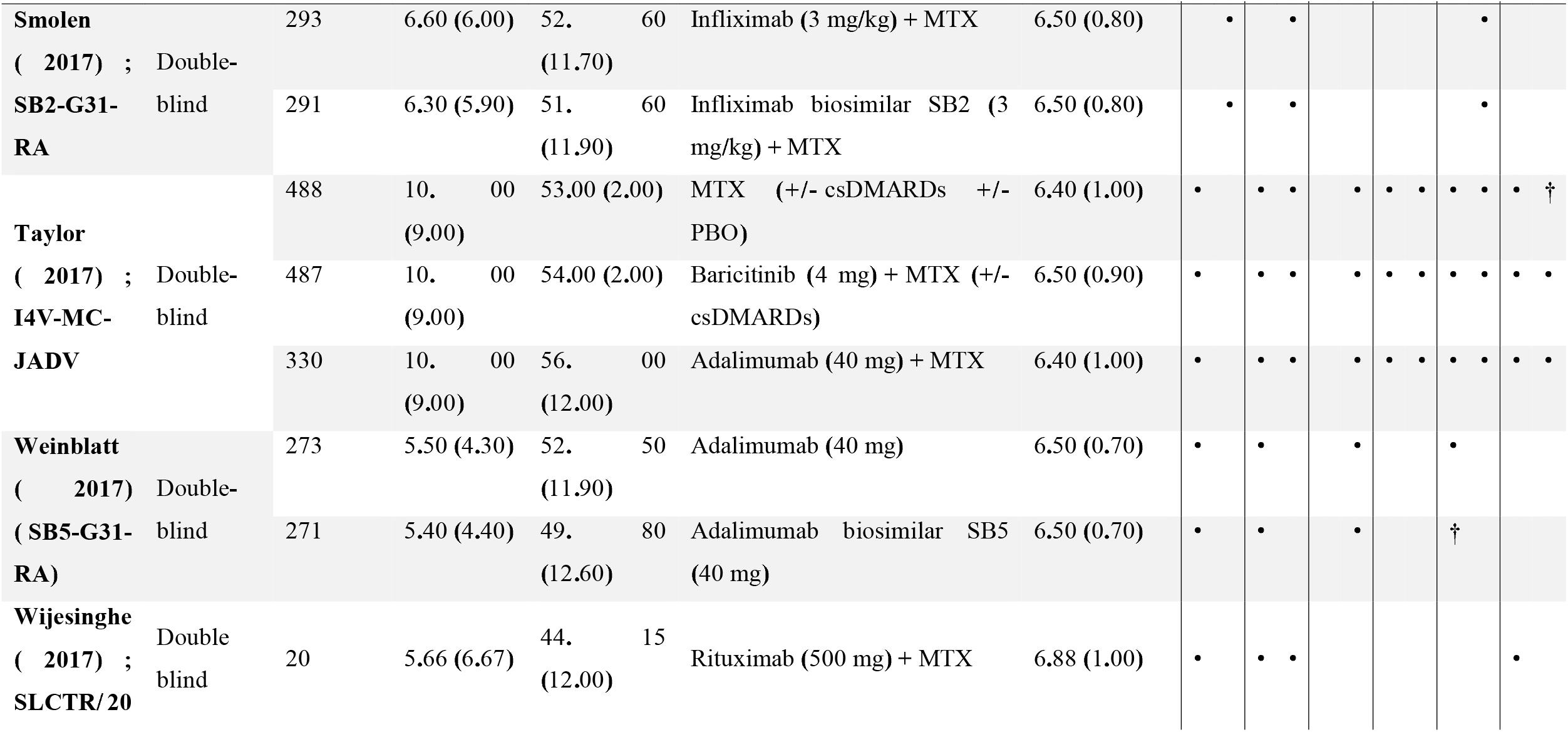
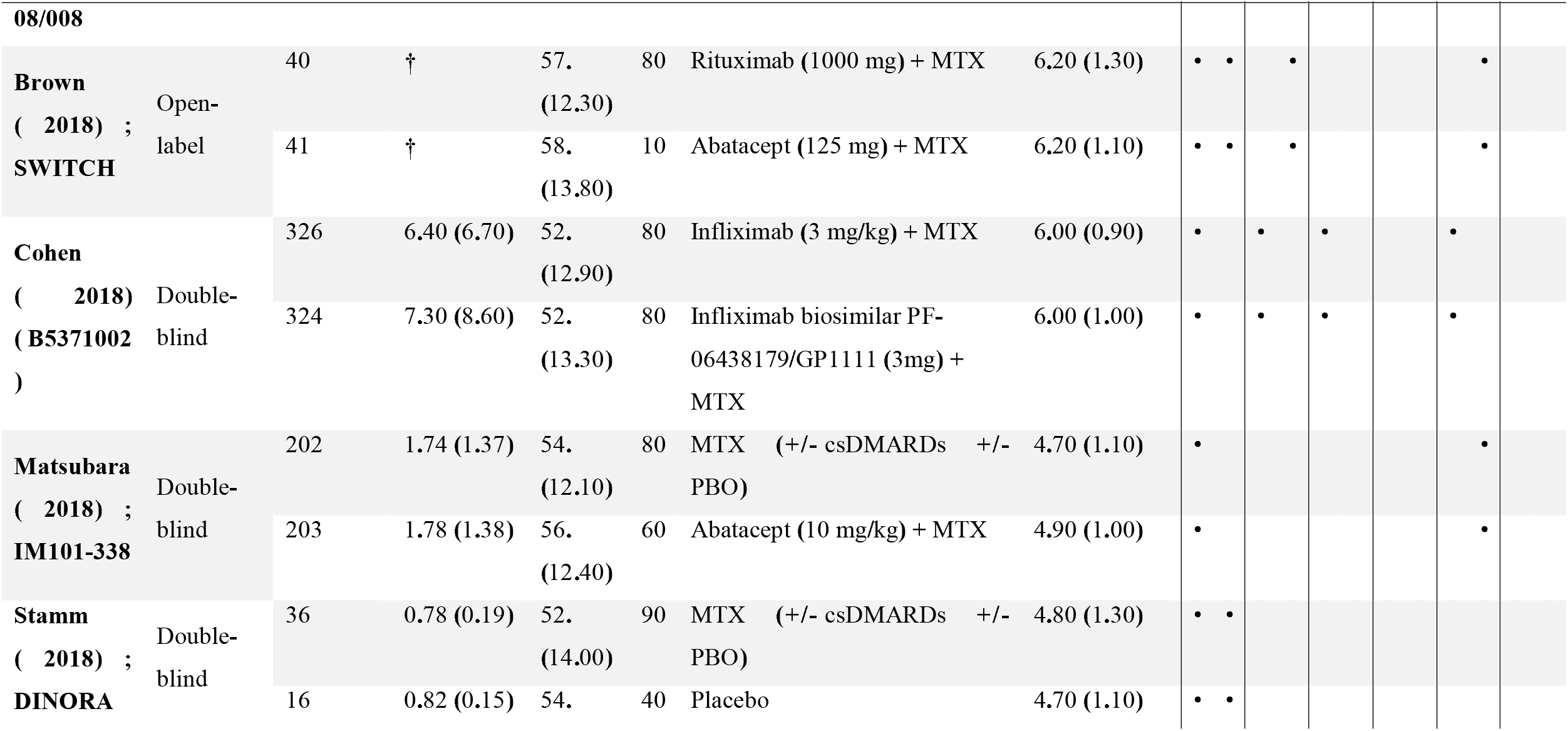
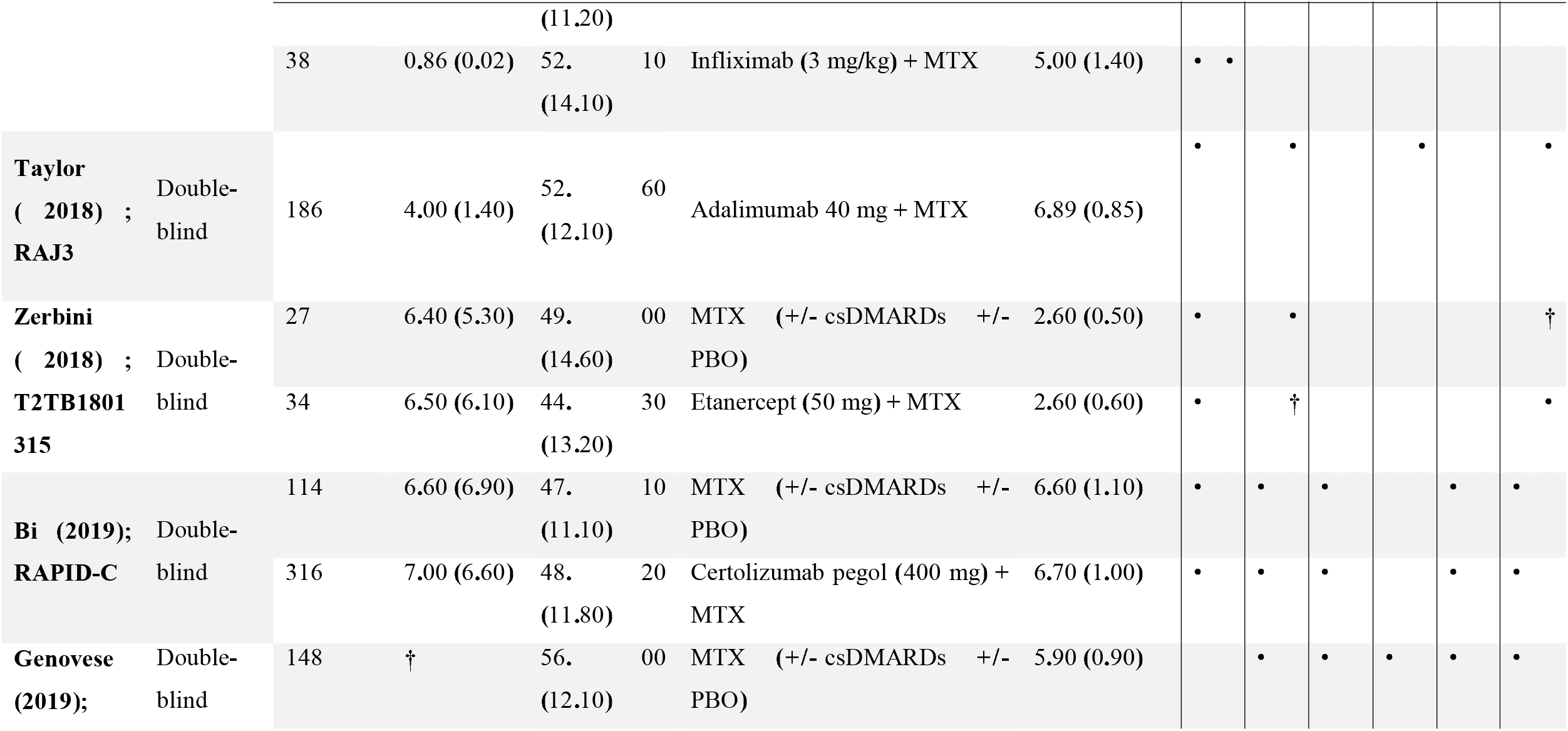
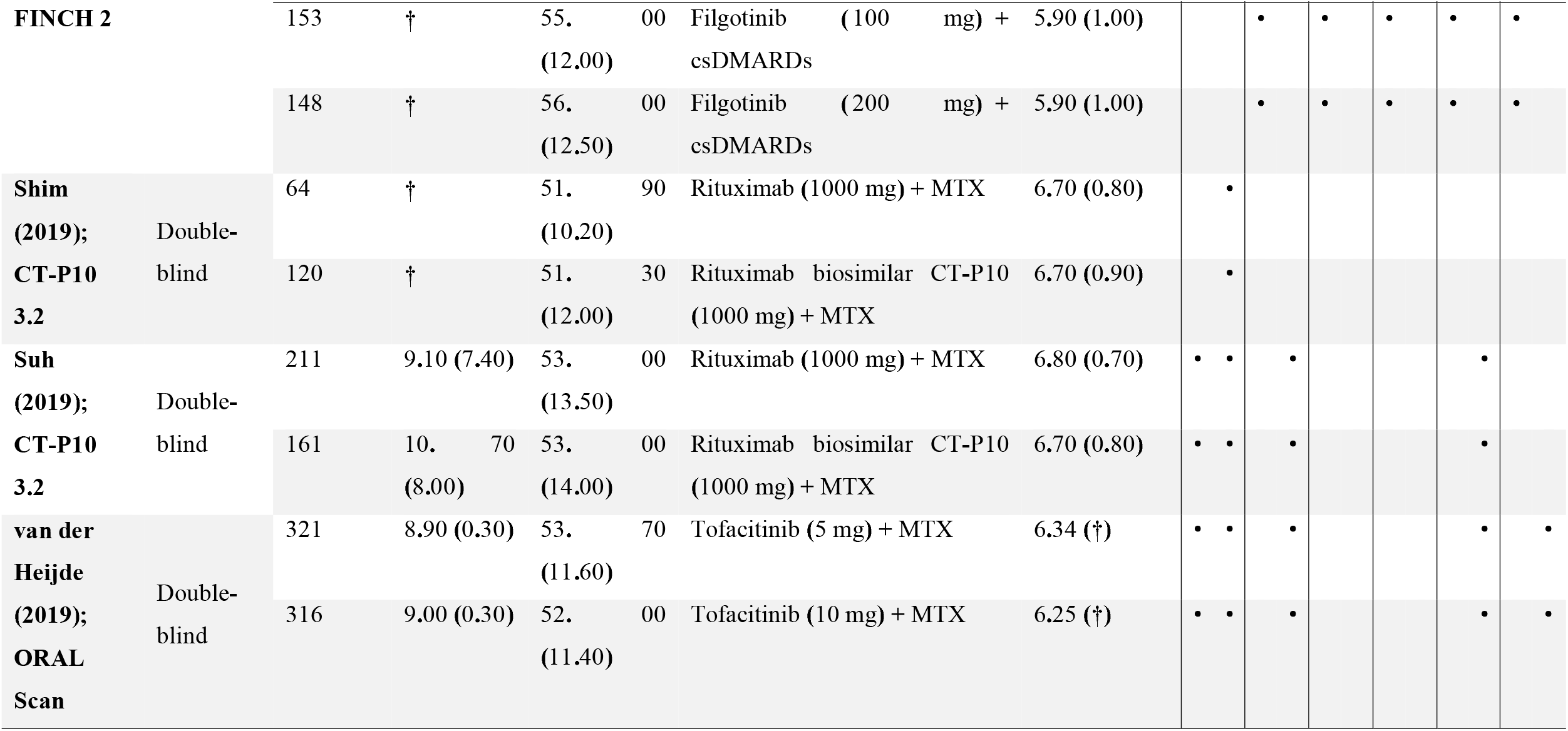
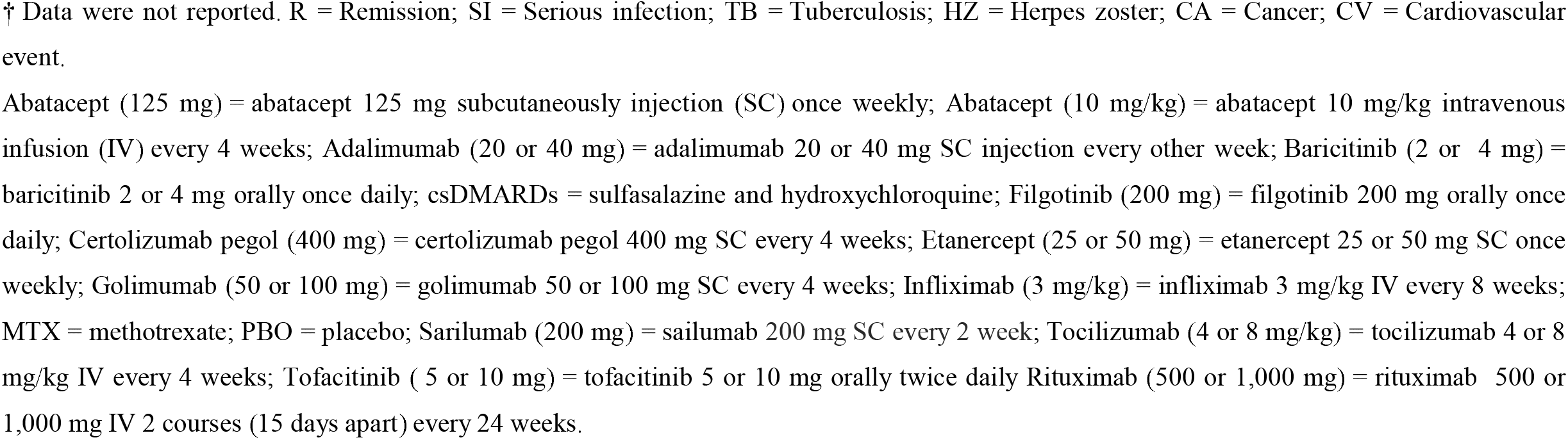
Study and patient characteristics included in the systematic review.

**Figure 1.**
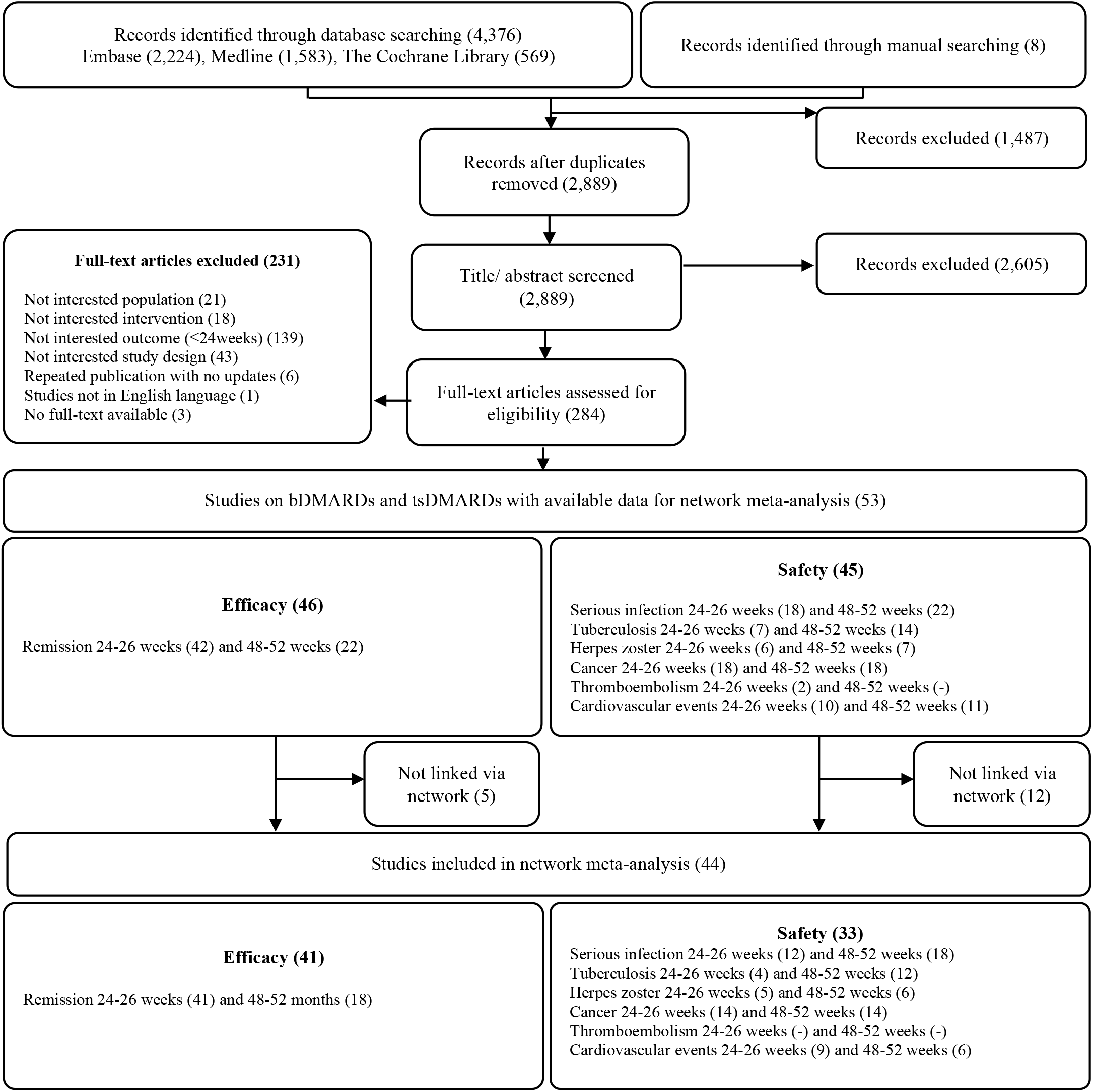
Evidence search and study selection.

### Network characteristics

For similarity and consistency assessment of network meta-analysis, 5 studies were excluded from efficacy analyses and 12 studies were excluded from safety outcomes (Figure 1). Fifty-three studies showed available data from which 44 studies were included in the final network meta-analysis for all outcomes. Of 46 studies included in the network meta-analysis for efficacy outcomes, 41 studies were included in remission at 24-26 weeks and 18 studies for 48-52 weeks. Of 45 studies included in network meta-analysis for safety outcomes, 33 were included for all safety outcomes. Remission outcomes at 24-26 and 48-52 weeks were analysed using a random-effects model. SAEs of TB at 24-26 weeks, HZ at both time points, cancer at 48-52 weeks, and CV events at both time points were analysed using a fixed-effects model. For efficacy outcomes, results for remission at 24-26 weeks were obtained using a consistency model (*p-value*=0.323). An inconsistency model was used for remission at 48-52 weeks (*p-value*=0.006). All safety outcomes were performed using a consistency model (*p-value* >0.05) (Supplementary 4). The available data were insufficient to perform a network meta-analysis for thromboembolism.

### Comparative effect of DMARDs

#### Efficacy

Remission at 24-26 weeks was obtained from 43 studies (n = 19,354) investigating 34 regimens (i.e., nine bDMARDs, two tsDMARDs, and nine biosimilars) (Table 2; Figure 2A; and Supplementary table 8). According to SUCRA ranking (Supplementary figure 1), the regimen with statistical significance that was ranked highest was tofacitinib (10 mg) + MTX [RR (95% CI), 4.65 (2.98-7.27)], followed by abatacept (10 mg/kg) + MTX [RR (95% CI), 3.74 (2.60-5.38)], and tocilizumab (8 mg/kg) + MTX [RR (95% CI) 3.06 (2.27-4.12)].

**Table 2.** Remission at 24-26 weeks

| **Ranking** | **Medication (dose)** | **Number of**<br>**study**<br>**(patients)** | **RR (95% CI)** |
| --- | --- | --- | --- |
| **1** | Tofacitinib (10 mg) + MTX | 3 (835) | **4.65** (2.98-7.27) |
| **2** | Abatacept (10 mg/kg) + MTX | 1 (203) | **3.74** (2.60-5.38) |
| **3** | Tocilizumab (8 mg/kg) + MTX | 2 (409) | **3.06** (2.27-4.12) |
| **4** | Tofacitinib (10 mg) | 1 (397) | **3.05** (2.03-4.57) |
| **5** | Golimumab (50 mg) + MTX | 3 (334) | **2.96** (2.03-4.32) |
| **6** | Etanercept biosimilar LBEC0101 + MTX | 1 (187) | **3.07** (1.87-5.05) |
| **7** | Etanercept biosimilar SB4 + MTX | 1 (299) | **2.95** (1.75-4.98) |
| **8** | Etanercept (50 mg) + MTX | 7 (1,054) | **2.85** (1.96-4.16) |
| **9** | Adalimumab biosimilar SB5 (40 mg) + MTX | 1 (271) | **2.81** (1.85-4.26) |
| **10** | Baricitinib (4 mg) + MTX +/- csDMARDs | 4 (1,106) | **2.71** (2.13-3.44) |
| **11** | Adalimumab (40 mg) + MTX | 8 (1,830) | **2.58** (2.03-3.28) |
| **12** | Infliximab biosimilar CT-P13 + MTX | 1 (302) | **2.62** (1.25-5.46) |
| **13** | Tocilizumab (8 mg/kg) | 2 (407) | **2.54** (1.88-3.44) |
| **14** | Tofacitinib (5 mg) + MTX | 4 (1,218) | **2.49** (1.70-3.65) |
| **15** | Etanercept biosimilar HD203 + MTX | 1 (147) | **2.38** (1.39-4.08) |
| **16** | Etanercept (50 mg) | 1 (98) | **2.37** (1.23-4.53) |
| **17** | Infliximab biosimilar PF-06438179/GP1111 + MTX | 1 (324) | **2.25** (1.17-4.33) |
| **18** | Baricitinib (2 mg) + MTX +/- csDMARDs | 2 (403) | **2.26** (1.49-3.43) |
| **19** | Etanercept (25 mg) + MTX | 3 (389) | **2.23** (1.64-3.04) |
| **20** | Golimumab (100 mg) + MTX | 3 (335) | **2.19** (1.47-3.27) |
| **21** | Adalimumab biosimilar ABP-501 + MTX | 1 (264) | **2.22** (1.58-3.12) |
| **22** | Tocilizumab (4 mg/kg) + MTX | 1 (290) | **2.13** (1.54-2.95) |
| **23** | Baricitinib (4 mg) | 1 (159) | **2.11** (1.42-3.14) |
| **24** | Tofacitinib (5 mg) | 2 (759) | **2.08** (1.46-2.97) |
| **25** | Golimumab (100 mg) | 1 (292) | **1.98** (1.30-3.03) |
| **26** | Infliximab (3 mg/kg) + MTX | 4 (961) | **1.95** (1.11-3.43) |
| **27** | Infliximab biosimilar SB2 + MTX | 1 (291) | 1.75 (0.89-3.47) |
| **28** | Certolizumab-pegol (400 mg) + MTX | 3 (634) | **1.75** (1.44-2.13) |
| **29** | Abatacept (125 mg) + MTX | 1 (41) | 1.15 (0.26-5.13) |
| **30** | Rituximab (1000 mg) + MTX | 4 (671) | **1.58** (1.04-2.40) |
| **31** | Rituximab (500 mg) + MTX | 3 (436) | 1.51 (0.99-2.31) |
| **32** | Placebo | 2 (81) | **1.45** (1.01-2.08) |
| **33** | Rituximab biosimilar CT-P10 + MTX | 1 (161) | 1.22 (0.60-2.51) |
| **34** | MTX | 24 (3,766) | Comparator |

Significant RR (CI) are in bold.

**Figure 2.**
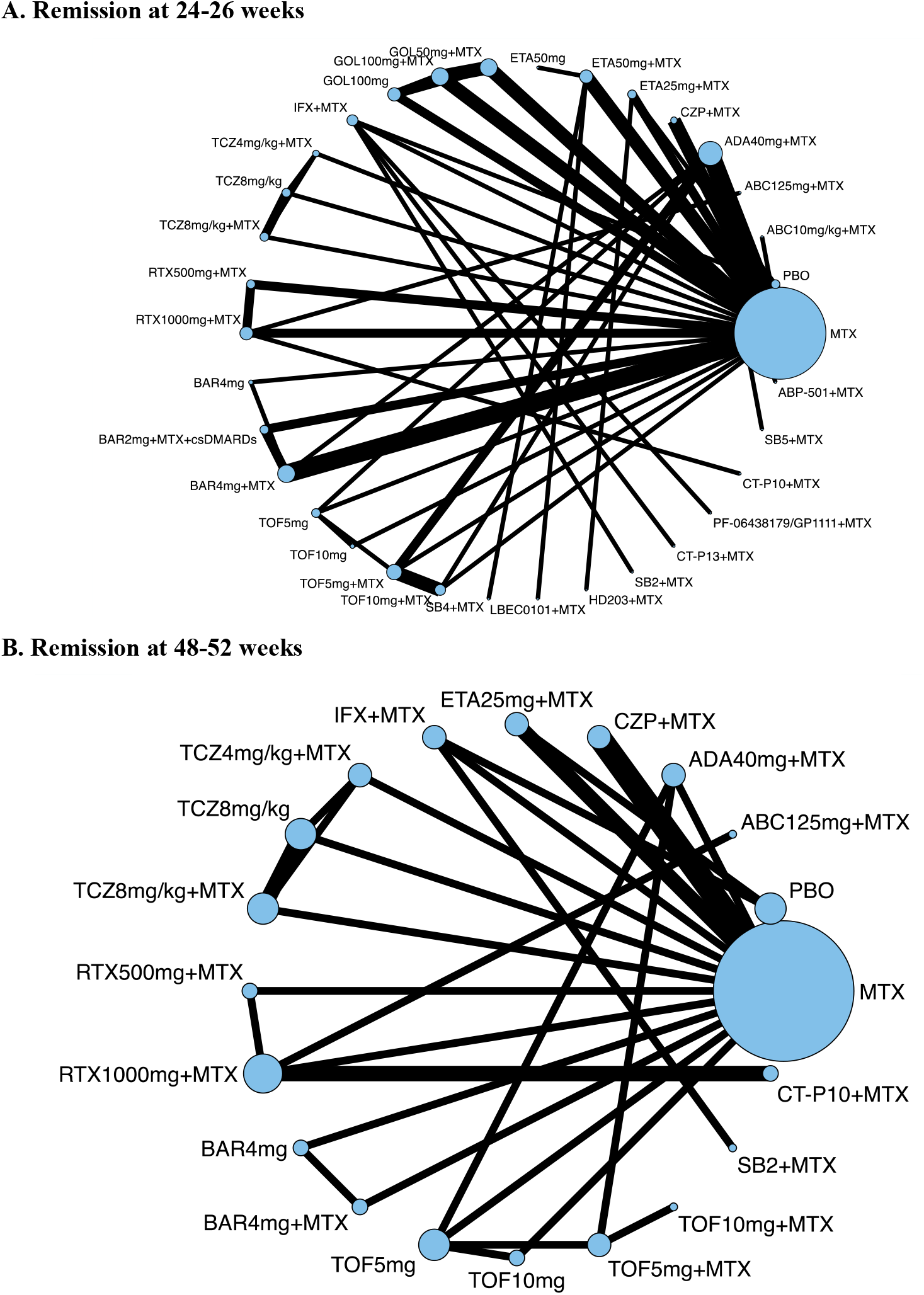
Network plots of treatment comparisons for remission at 24-26 weeks (A) and 48-52 weeks (B). MTX = methotrexate, PBO = Placebo; ABC125mg/kg+MTX = abatacept 125 mg subcutaneously injection (SC) once weekly + MTX; ABC10mg/kg+MTX = abatacept 10 mg/kg intravenous infusion (IV) every 4 weeks + MTX; ADA40mg+MTX = adalimumab 40 mg SC injection every other week + MTX; ABP-501+MTX = Adalimumab biosimilar 40 mg; BAR4mg = baricitinib 4 mg orally once daily; BAR2mg+MTX+csDMARDs = baricitinib 2 mg orally once daily + MTX + csDMARDs; BAR4mg+MTX = baricitinib 4 mg orally once daily + MTX; CT-P10+MTX = Biosimilar rituximab 1000 mg + MTX; CT-P13+MTX = Infliximab biosimilar 3 mg/kg + MTX; CZP+MTX = certolizumab pegol 400 mg SC every 4 weeks + MTX; ETA25mg+MTX = etanercept 25 mg SC once weekly + MTX, ETA50mg+MTX = etanercept 50 mg SC once weekly + MTX, ETA50mg = etanercept 50 mg SC once weekly, GOL50mg+MTX = golimumab 50 mg SC every 4 weeks + MTX; GOL100mg+MTX = golimumab 100 mg SC every 4 weeks + MTX; GOL100mg = golimumab 100 mg SC every 4 weeks; HD203+MTX = Etanercept biosimilar 25 mg twice weekly SC; IFX+MTX = infliximab 3 mg/kg IV every 8 weeks; LBEC0101+MTX = Etanercept biosimilar 50 mg once weekly SC + MTX; PF-06438179/GP1111+MTX = infliximab biosimilar 3 mg/kg + MTX; RTX500mg+MTX = rituximab 500 mg IV 2 courses (15 days apart) every 24 weeks + MTX; RTX1000mg+MTX = rituximab 500 mg IV 2 courses (15 days apart) every 24 weeks + MTX; SB2+MTX = Infliximab biosimilar 3 mg/kg + MTX; SB4+MTX = Etanercept biosimilar 50 mg SC once weekly + MTX; SB5+MTX = SB5 Adalimumab biosimilar 40 mg every other week SC + MTX; TCZ4mg/kg+MTX = tocilizumab 4 mg/kg IV every 4 weeks + MTX, TCZ8mg/kg = tocilizumab 8 mg/kg IV every 4 weeks, TCZ8mg/kg+MTX = tocilizumab 8 mg/kg IV every 4 weeks + MTX; TOF5mg = tofacitinib 5 mg orally twice daily, TOF10mg = tofacitinib 10 mg orally twice daily; TOF5mg+MTX = tofacitinib 5 mg orally twice daily + MTX; TOF10mg+MTX = tofacitinib 10 mg orally twice daily + MTX.

Remission at 48-52 weeks was obtained from 31 studies (n = 9,027) for a total of 20 regimens (i.e., seven bDMARDs and two tsDMARDs in different doses and two biosimilars) (Table 3; Figure 2B; Supplementary Table 8). According to the SUCRA ranking (Supplementary figure 2), the regimen with statistical significance with the highest ranking was tocilizumab (8 mg/kg) + MTX [RR (95% CI) 2.52 (1.94-3.28)], followed by rituximab (1000 mg) + MTX [RR (95% CI) 2.38 (1.65-3.45), tocilizumab (8 mg/kg) monotherapy [RR (95% CI) 2.03 (1.54-2.68), and tofacitinib (10 mg) [RR (95% CI) 1.96 (1.27-3.03).

**Table 3.**
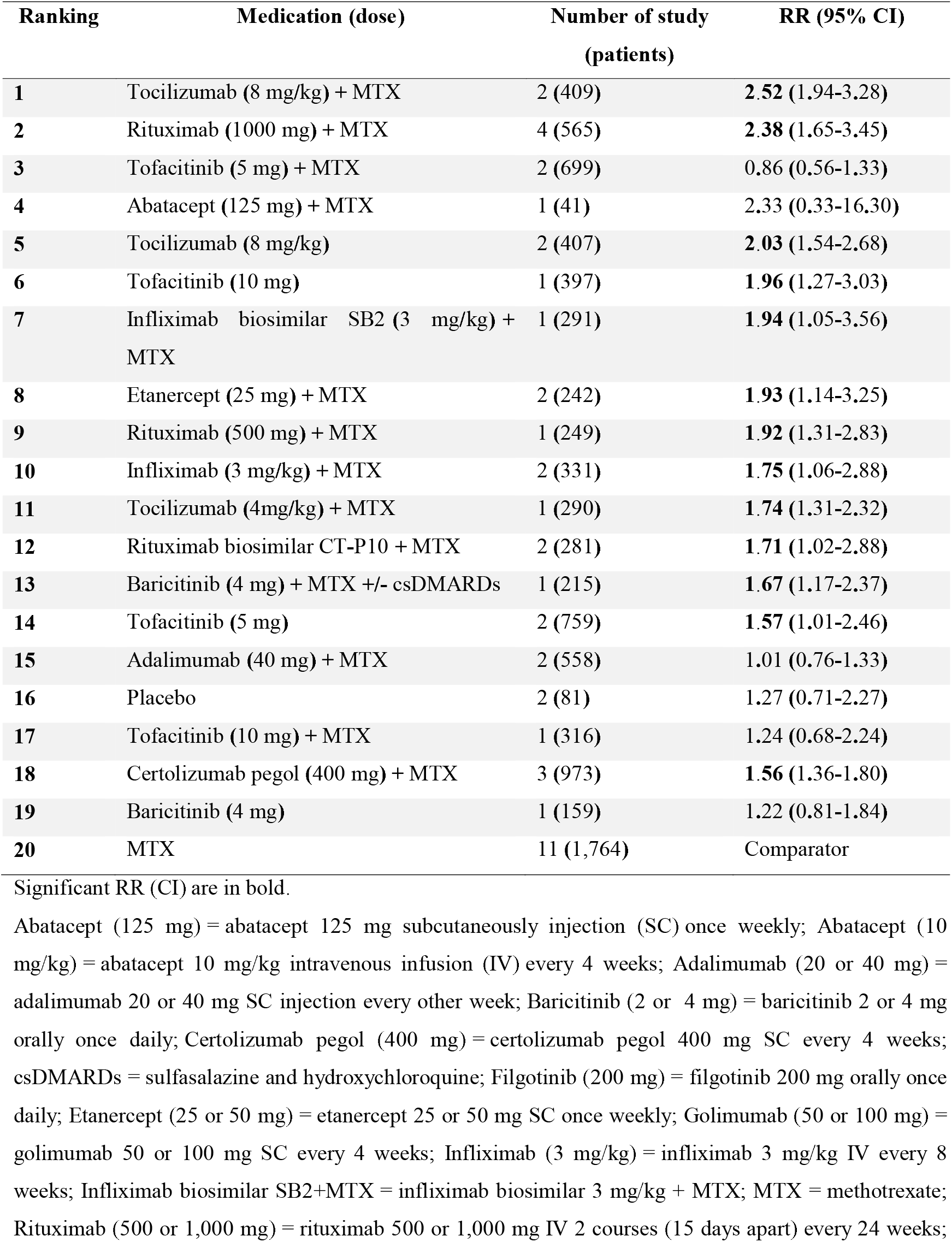
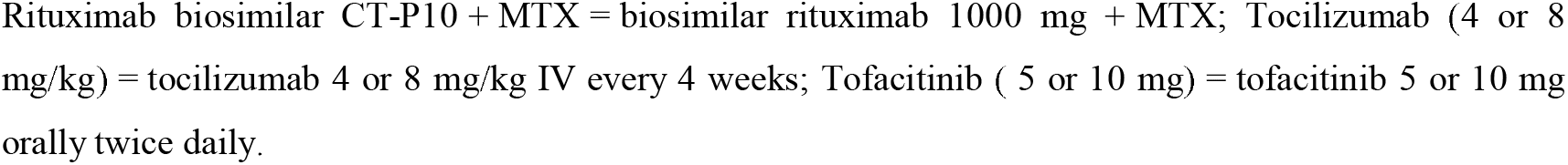
Remission at 48-52 weeks

| Ranking | Medication (dose) | Number of study<br>(patients) | RR (95% CI) |
| --- | --- | --- | --- |
| **1** | Tocilizumab (8 mg/kg) + MTX | 2 (409) | **2.52** (1.94-3.28) |
| **2** | Rituximab (1000 mg) + MTX | 4 (565) | **2.38** (1.65-3.45) |
| **3** | Tofacitinib (5 mg) + MTX | 2 (699) | 0.86 (0.56-1.33) |
| **4** | Abatacept (125 mg) + MTX | 1 (41) | 2.33 (0.33-16.30) |
| **5** | Tocilizumab (8 mg/kg) | 2 (407) | **2.03** (1.54-2.68) |
| **6** | Tofacitinib (10 mg) | 1 (397) | **1.96** (1.27-3.03) |
| **7** | Infliximab biosimilar SB2 (3 mg/kg) + MTX | 1 (291) | **1.94** (1.05-3.56) |
| **8** | Etanercept (25 mg) + MTX | 2 (242) | **1.93** (1.14-3.25) |
| **9** | Rituximab (500 mg) + MTX | 1 (249) | **1.92** (1.31-2.83) |
| **10** | Infliximab (3 mg/kg) + MTX | 2 (331) | **1.75** (1.06-2.88) |
| **11** | Tocilizumab (4mg/kg) + MTX | 1 (290) | **1.74** (1.31-2.32) |
| **12** | Rituximab biosimilar CT-P10 + MTX | 2 (281) | **1.71** (1.02-2.88) |
| **13** | Baricitinib (4 mg) + MTX +/- csDMARDs | 1 (215) | **1.67** (1.17-2.37) |
| **14** | Tofacitinib (5 mg) | 2 (759) | **1.57** (1.01-2.46) |
| **15** | Adalimumab (40 mg) + MTX | 2 (558) | 1.01 (0.76-1.33) |
| **16** | Placebo | 2 (81) | 1.27 (0.71-2.27) |
| **17** | Tofacitinib (10 mg) + MTX | 1 (316) | 1.24 (0.68-2.24) |
| **18** | Certolizumab pegol (400 mg) + MTX | 3 (973) | **1.56** (1.36-1.80) |
| **19** | Baricitinib (4 mg) | 1 (159) | 1.22 (0.81-1.84) |
| **20** | MTX | 11 (1,764) | Comparator |

Significant RR (CI) are in bold.
Abatacept (125 mg) = abatacept 125 mg subcutaneously injection (SC) once weekly; Abatacept (10 mg/kg) = abatacept 10 mg/kg intravenous infusion (IV) every 4 weeks; Adalimumab (20 or 40 mg) = adalimumab 20 or 40 mg SC injection every other week; Baricitinib (2 or 4 mg) = baricitinib 2 or 4 mg orally once daily; Certolizumab pegol (400 mg) = certolizumab pegol 400 mg SC every 4 weeks; csDMARDs = sulfasalazine and hydroxychloroquine; Filgotinib (200 mg) = filgotinib 200 mg orally once daily; Etanercept (25 or 50 mg) = etanercept 25 or 50 mg SC once weekly; Golimumab (50 or 100 mg) = golimumab 50 or 100 mg SC every 4 weeks; Infliximab (3 mg/kg) = infliximab 3 mg/kg IV every 8 weeks; Infliximab biosimilar SB2+MTX = infliximab biosimilar 3 mg/kg + MTX; MTX = methotrexate; Rituximab (500 or 1,000 mg) = rituximab 500 or 1,000 mg IV 2 courses (15 days apart) every 24 weeks;
Rituximab biosimilar CT-P10 + MTX = biosimilar rituximab 1000 mg + MTX; Tocilizumab (4 or 8 mg/kg) = tocilizumab 4 or 8 mg/kg IV every 4 weeks; Tofacitinib ( 5 or 10 mg) = tofacitinib 5 or 10 mg orally twice daily.

#### Safety

The results from our network meta-analysis are shown in Figures 3 and 4. Serious infection at 24-26 and 48-52 weeks was obtained from 12 studies (n = 7,610) and 18 studies (n = 9,304), respectively. A total of 14 regimens (i.e., four bDMARDs, two tsDMARDs, and one biosimilar), and 16 regimens (i.e., five bDMARDs, two tsDMARDs, and one biosimilar) were investigated for 48-52 weeks. No regimen was found to have statistically significant concerns of an increased risk of serious infection when compared to MTX at both time points (Supplementary Table 9, 10 and 19; Supplementary figure 3-4 for SUCRA ranking).

**Figure 3.**
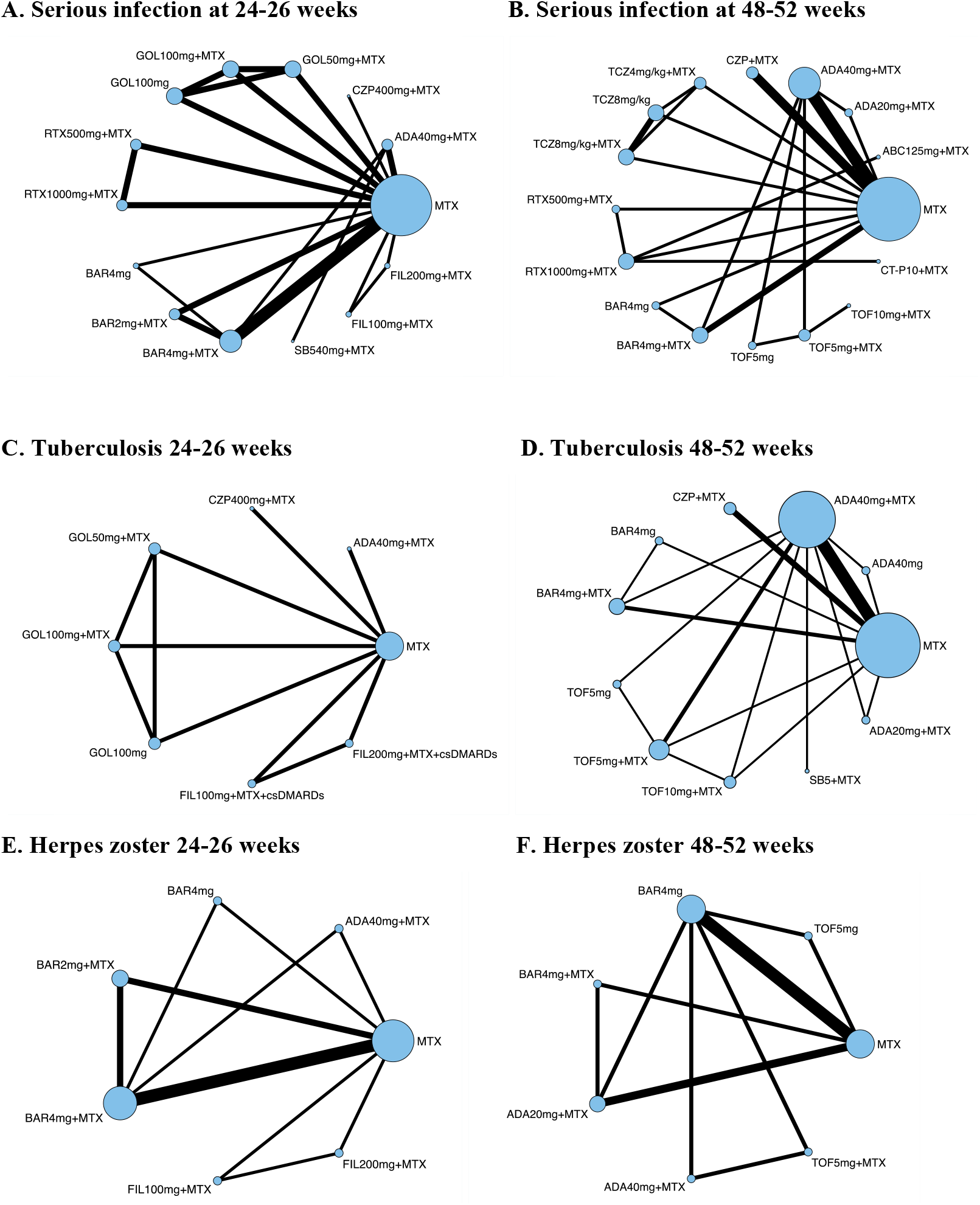
Network plots of treatment comparisons for serious infection, TB and HZ at 24-26 weeks and 48-52 weeks HZ = herpes zoster; TB = tuberculosis. ABP-501+MTX = Adalimumab biosimilar ABP-501 40 mg; ADA40mg+MTX = adalimumab 40 mg SC injection every other week + MTX; BAR4mg = baricitinib 4 mg orally once daily; BAR2mg+MTX = baricitinib 2 mg orally once daily; CZP+MTX = certolizumab pegol 400 mg SC every 4 weeks + MTX; FIL100mg = filgotinib 100 mg orally once daily; FIL200mg = filgotinib 200 mg orally once daily; GOL50mg+MTX = golimumab 50 mg SC every 4 weeks + MTX; GOL100mg+MTX = golimumab 100 mg SC every 4 weeks + MTX; GOL100mg = golimumab 100 mg SC every 4 weeks; MTX = methotrexate; RTX500mg+MTX = rituximab 500 mg IV 2 courses (15 days apart) every 24 weeks + MTX; RTX1000mg+MTX = rituximab 1000 mg IV 2 courses (15 days apart) every 24 weeks + MTX; SB5+MTX = Adalimumab biosimilar SB5 40 mg every other week SC + MTX; TCZ4mg/kg+MTX = tocilizumab 4 mg/kg IV every 4 weeks + MTX; TCZ8mg/kg = tocilizumab 8 mg/kg IV every 4 weeks; TCZ8mg/kg+MTX = tocilizumab 8 mg/kg IV every 4 weeks + MTX; TOF5mg = tofacitinib 5 mg orally twice daily; TOF10mg = tofacitinib 10 mg orally twice daily; TOF5mg+MTX = tofacitinib 5 mg orally twice daily + MTX; TOF10mg+MTX = tofacitinib 10 mg orally twice daily + MTX.

**Figure 4.**
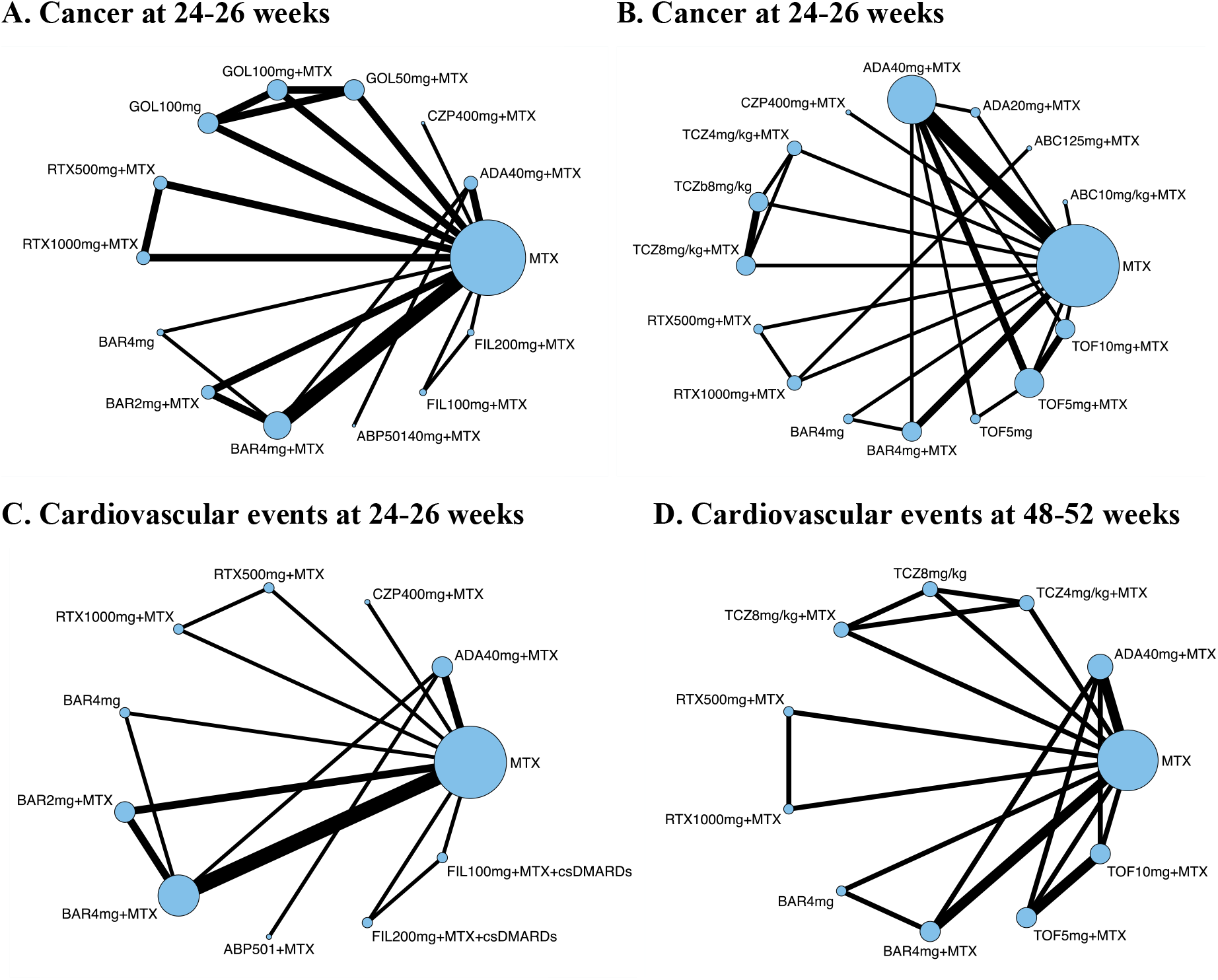
Network plots of treatment comparisons for cancer and CV events at 24-26 weeks and 48-52 weeks CV = cardiovascular. ABC10mg/kg+MTX = abatacept 10 mg/kg intravenous infusion (IV) every 4 weeks + MTX; ABC125mg/kg+MTX= abatacept 125 mg subcutaneously injection (SC) once weekly + MTX; ABP-501+MTX = Adalimumab biosimilar 40 mg; ADA40mg+MTX = adalimumab 40 mg SC injection every other week + MTX; BAR4mg = baricitinib 4 mg orally once daily; BAR2mg+MTX = baricitinib 2 mg orally once daily + MTX + csDMARDs; BAR4mg+MTX = baricitinib 4 mg orally once daily + MTX; CZP+MTX = certolizumab pegol 400 mg SC every 4 weeks + MTX; ETA25mg+MTX = etanercept 25 mg SC once weekly + MTX; ETA50mg+MTX = etanercept 50 mg SC once weekly + MTX, ETA50mg = etanercept 50 mg SC once weekly; FIL100mg = filgotinib 100 mg orally once daily, FIL200mg = filgotinib 200 mg orally once daily; GOL50mg+MTX = golimumab 50 mg SC every 4 weeks + MTX; GOL100mg+MTX = golimumab 100 mg SC every 4 weeks + MTX, GOL100mg = golimumab 100 mg SC every 4 weeks; IFX+MTX = infliximab 3 mg/kg IV every 8 weeks; MTX = methotrexate; PBO= Placebo; TCZ4mg/kg+MTX = tocilizumab 4 mg/kg IV every 4 weeks + MTX; TCZ8mg/kg = tocilizumab 8 mg/kg IV every 4 weeks; TCZ8mg/kg+MTX = tocilizumab 8 mg/kg IV every 4 weeks + MTX; RTX500mg+MTX = rituximab 500 mg IV 2 courses (15 days apart) every 24 weeks + MTX; RTX1000mg+MTX = rituximab 500 mg IV 2 courses (15 days apart) every 24 weeks + MTX; TOF5mg = tofacitinib 5 mg orally twice daily; TOF10mg = tofacitinib 10 mg orally twice daily; TOF5mg+MTX = tofacitinib 5 mg orally twice daily + MTX; TOF10mg+MTX = tofacitinib 10 mg orally twice daily + MTX;

TB at 24-26 and 48-52 weeks were obtained from four studies (n = 2,548) and 12 studies (n = 7,741), respectively. A total of eight regimens (i.e., three bDMARDs and one tsDMARD) were investigated for 24-26 weeks, and 10 regimens (i.e., three bDMARDs, two tsDMARDs and one biosimilar) were investigated for 48-52 weeks. No regimen significantly increased the risk of TB at both time points (Supplementary Table 11, 12 and 20; Supplementary figure 5-6 for SUCRA ranking).

HZ at 24-26 and 48-52 weeks was obtained from five studies (n = 3,549) and six studies (n = 4,034), respectively. A total of seven regimens (i.e., one bDMARDs and two tsDMARDs) were investigated for 24-26 weeks, and seven regimens (i.e., one bDMARDs and two tsDMARDs) were investigated for 48-52 weeks. Baricitinib (4 mg) + MTX showed an increased risk of HZ [RR (95%CI) 3.52 (1.38-9.02)] at 24-26 weeks (Supplementary Table 13 and 21) and at 48-52 weeks [RR (95%CI) 4.20 (1.22-14.48)] (Supplementary Table 14). Tofacitinib (5 mg) + MTX [RR (95%CI) 5.38 (1.00-28.91)] was also found to increase the risk HZ at 48-52 weeks (Supplementary Table 13, 14 and 21; Supplementary figure 7-8 for SUCRA ranking).

Cancer at 24-26 and 48-52 weeks was obtained from 14 studies (n = 7,881) and 13 studies (n = 8,292), respectively. A total of 14 regimens (i.e., four bDMARDs, two tsDMARDs and one biosimilar) were investigated for 24-26 weeks, and 16 regimens (i.e., six bDMARDs and two tsDMARDs) were investigated for 48-52 weeks. No regimen significantly increased the risk of cancer at both time points (Supplementary table 15, 16 and 22; Supplementary figure 9-10 for SUCRA ranking).

CV events at 24-26 and 48-52 weeks were obtained from nine studies (n = 6,046) and six studies (n = 5,153), respectively. A total of 11 regimens (i.e., three bDMARDs, two tsDMARDs and one biosimilar) were investigated for 24-26 weeks, and 11 regimens (i.e., three bDMARDs and two tsDMARDs) were investigated for 48-52 weeks. For both time points, none of the regimens was found to increase the risk of CV events (Supplementary table 17, 18 and 23; Supplementary figure 11-12 for SUCRA ranking).

## Discussion

This systematic review and network meta-analysis showed that bDMARDs and tsDMARDs monotherapy, as well as with MTX, had superior treatment effects compared to MTX alone. Based on SUCRA, tofacitinib (10 mg) + MTX was ranked with the highest efficacy at remission 24-26 weeks. Tocilizumab (8 mg/kg) + MTX was the third rank for efficacy for remission at 24-26 weeks, and the highest at 48-52 weeks. Safety outcomes showed an increased risk of HZ compared between bDMARDs/ tsDMARDs and MTX including baricitinib (4 mg) + MTX and tofacitinib (5 mg) + MTX. There were no statistically significant safety concerns for other SAEs.

Due to the use of different clinical outcomes in previous studies (17, 27, 28), direct comparison of treatment effect values among studies is not appropriate. However, compared to a study by Hazlewood et al. (17), we found the treatment effects of most regimens to be lower. These discrepancies could be attributed specifically to differences in dosage and the time points analysed, which in our case are strictly specified. Other factors may include differences in the selected common comparator and the analysis approach used (Bayesian vs. frequentist methods) (21, 23), as well as trial duration and study population (MTX naïve vs. MTX-inadequate responders). Nevertheless, tocilizumab (8 mg/ kg) + MTX showed the highest treatment effect in their study, the same as in our remission outcome at 48-52 weeks.

Comparing our SAEs results with other studies confirmed no statistically significant safety concerns for serious infection, TB, cancer and CV events associated with the use of DMARDs (28). We observed no differences in SAEs results between 24-26 weeks and 48-52 weeks, which may be because we separately analysed safety outcomes according to these time points, resulting in lower number of events and participants available for analyses. Moreover, the association between bDMARDs/tsDMARDs and an increased risk of cancer and CV events remained inconclusive. Our network meta-analysis did not demonstrate a risk of these SAEs, as previously reported by the USFDA (13, 14). This could be explained by the USFDA’s long-term real-world monitoring of the drug’s performance.

### Strengths and limitations of this study

Previous studies focused on comparing treatment effects among bDMARDs (27, 28), and tofacitinib was the only tsDMARD considered in a previous network meta-analyses (17). To our knowledge, this network meta-analysis was the first to compare a wide range of both bDMARDs, biosimilars and tsDMARDs, including baricitinib and filgotinib. We also analysed each medication in a dose- and time point-specific manner, generating a specific profile of each medication. While other studies assessed efficacy based on ACR response criteria (commonly used in clinical trials and defined as a joint involvement index) (17, 27) and Clinical Disease Activity Index (CDAI) (28), our analyses used DAS28-ESR remission. As DAS28-ESR Remission is a widely used outcome measure in routine practice (3, 29–31), it is a practical choice and applicable for monitoring treatment effects in a clinical practice setting.

The previously reported hierarchy of treatment effect of DMARDs was different among studies. While others report only point estimates of the mean relative or absolute treatment effects (17, 27, 28), our study used SUCRA, which indicates which medication is ranked better over the largest fraction of competitors (32). The hierarchy generated by SUCRA depends on the rankings of all treatments considered and the ranking of medications with high uncertainty is not under- or over-estimated (32). Our results involve outcomes with a wide range of CI and therefore SUCRA was the chosen method to generate a more impartial ranking as it takes the uncertainty of the effect size estimates and their precision into account (32).

We performed network meta-analyses on five substantial SAEs, of which TB, HZ and CV events have not been reported in other network meta-analyses before. By ranking DMARDs according to both treatment effect and safety profile, a more comprehensive overview of each medication’s performance can be compared. This is important information when choosing the most appropriate regimens for this group of RA patients.

Nevertheless, this network meta-analysis has several limitations. There were nine of 53 studies excluded from our analysis after assessing for network connectivity since there was no common comparator (i.e., MTX), which reduced the data available for analyses. RCTs of biosimilars with the defined outcomes were also limited. Data related to long-term safety outcomes beyond 52 weeks were insufficient for analyses. This study focused on only RA patients who failed MTX and had high disease activity, so the results should not be directly applied to RA patients who were MTX-naïve, failed MTX, sulfasalazine and hydroxychloroquine, or had moderate disease activity. Although this network meta-analysis could demonstrate additional indirect comparisons among multiple interventions that aim for similar treatment outcomes where direct comparisons are not available, network meta-analysis cannot compensate for a lack of direct comparisons that are needed and a priority for future research.

### Conclusion and policy recommendation

For treatment of RA patients who had inadequate responses to MTX, bDMARDs, tsDMARDs, and their biosimilars, both monotherapy or in combination with MTX, had better treatment outcomes compared to MTX monotherapy with modest safety concerns at 24-52 weeks. However, long-term efficacy and safety information should be rigorously monitored in routine healthcare services as well as real-time post-market surveillance to produce additional safety reports for all utilised products. For further health technology assessment, this synthesis of the evidence through network meta-analysis can provide important data for cost-effectiveness analysis and could be used to inform future policy decisions.

## Supporting information

PRISMA_2020_abstract_checklist

PRISMA_2020_checklist

## Data Availability

Main data produced in the present work are contained in the manuscript. Additional data produced in the present work are available upon reasonable request to the authors.

## Acknowledgement

We would like to thank Chatkamol Pheerapanyawaranun for her contribution in formatting the supplement sections and Disorn Kulpokin for his contribution in generating the Risk of Bias section.

