## Supplementary material for "Efficacy and safety of biologic, biosimilars and targeted synthetic DMARDs in moderate-to-severe rheumatoid arthritis with inadequate response to methotrexate: a systematic review and network meta-analysis"

### **Methods Supplement**

#### **Supplementary 1 Search strategy**

##### *Supplement table 1 Search term Embase*

| No. | Search term |
| --- | --- |
| 1 | exp AND ('arthritis,'/exp OR 'arthritis,' OR 'arthritis,'/exp OR arthritis,) AND rheumatoid:ab,kw,ti |
| 2 | (rheumatoid OR reumatoid OR revmatoid OR rheumatic OR reumatic OR revmatic OR rheumat$ OR reumat$ OR revmarthrit$) AND (arthrit$:ab,kw,ti OR artrit$:ab,kw,ti OR diseas$:ab,kw,ti OR condition$:ab,kw,ti OR nodule$:ab,kw,ti) |
| 3 | felty$:ab,kw,ti AND syndrome:ab,kw,ti |
| 4 | caplan$:ab,kw,ti AND syndrome:ab,kw,ti |
| 5 | sjogren$:ab,kw,ti AND syndrome:ab,kw,ti |
| 6 | sicca:ab,kw,ti AND syndrome:ab,kw,ti |
| 7 | still$ AND disease:ab,kw,ti |
| 8 | #1 OR #2 OR #3 OR #4 OR #5 OR #6 OR #7 |
| 9 | methotrexate:ab,kw,ti |
| 10 | mexate:ab,kw,ti |
| 11 | abitrexate:ab,kw,ti |
| 12 | amet?opterine:ab,kw,ti |
| 13 | abitrexate:ab,kw,ti |
| 14 | a AND met?opterine:ab,kw,ti |
| 15 | antifolan:ab,kw,ti |
| 16 | emt?exate:ab,kw,ti |
| 17 | enthexate:ab,kw,ti |
| 18 | farmitrexate:ab,kw,ti |
| 19 | folex:ab,kw,ti |
| 20 | ledertrexate:ab,kw,ti |
| 21 | methoblastin:ab,kw,ti |
| 22 | methohexate:ab,kw,ti |
| 23 | methotrate:ab,kw,ti |
| 24 | methylaminopterin:ab,kw,ti |
| 25 | metotrexat$:ab,kw,ti |
| 26 | mtx:ab,kw,ti |
| 27 | novatrex:ab,kw,ti |
| 28 | rheumatrex:ab,kw,ti |
| 29 | etanercept:ab,kw,ti |
| 30 | infliximab:ab,kw,ti |
| 31 | golimumab:ab,kw,ti |
| 32 | adalimumab:ab,kw,ti |
| 33 | tocilizumab:ab,kw,ti |
| 34 | rituximab:ab,kw,ti |
| 35 | tofacitinib:ab,kw,ti |
| 36 | baricitinib:ab,kw,ti |
| 37 | certolizumab AND pegol:ab,kw,ti |
| 38 | sarilumab:ab,kw,ti |
| 39 | abatacept:ab,kw,ti |
| 40 | filgotinib:ab,kw,ti |
| 41 | #9 OR #10 OR #11 OR #12 OR #13 OR #14 OR #15 OR #16 OR #17 OR #18 OR #19 OR #20 OR #21 OR #22 OR #23 OR #24 OR #25 OR #26 OR #27 OR #28 OR #29 OR #30 OR #31 OR #32 OR #33 OR #34 OR #35 OR #36 OR #37 OR #38 OR #39 OR #40 |
| 42 | #8 AND #41 |
| 43 | #42 AND [embase]/lim |
| 44 | #43 AND [randomized controlled trial]/lim |

##### *Supplement table 2 Search term MEDLINE via Ovid*

| No. | Search term |
| --- | --- |
| 1 | exp arthritis, rheumatoid/ |
| 2 | ((rheumatoid or reumatoid or revmatoid or rheumatic or reumatic or revmatic or rheumat$ or reumat$ or revmarthrit$) adj3 (arthrit$ or artrit$ or diseas$ or condition$ or nodule$)).tw. |
| 3 | (felty$ adj2 syndrome).tw. |
| 4 | (caplan$ adj2 syndrome).tw. |
| 5 | (sjogren$ adj2 syndrome).tw. |
| 6 | (sicca adj2 syndrome).tw. |
| 7 | still$ disease.tw. |
| 8 | 1 or 2 or 3 or 4 or 5 or 6 or 7 |
| 9 | Methotrexate/ |
| 10 | Methotrexate.tw. |
| 11 | amet?opterine.tw. |
| 12 | mexate.tw. |
| 13 | Abitrexate.tw. |
| 14 | A Met?opterine.tw. |
| 15 | Antifolan.tw. |
| 16 | Emt?exate.tw. |
| 17 | Enthexate.tw. |
| 18 | Farmitrexate.tw. |
| 19 | Folex.tw. |
| 20 | Ledertrexate.tw. |
| 21 | Methoblastin.tw. |
| 22 | Methohexate.tw. |
| 23 | Methotrate.tw. |
| 24 | Methylaminopterin.tw. |
| 25 | Metotrexate.tw. |
| 26 | Mtx.tw. |
| 27 | Novatrex.tw. |
| 28 | Rheumatrex.tw. |
| 29 | Etanercept.tw. |
| 30 | Infliximab.tw. |
| 31 | golimumab.tw. |
| 32 | Adalimumab.tw. |
| 33 | tocilizumab.tw. |
| 34 | rituximab.tw. |
| 35 | tofacitinib.tw. |
| 36 | baricitinib.tw. |
| 37 | (certolizumab and pegol).tw. |
| 38 | sarilumab.tw. |
| 39 | abatacept.tw. |
| 40 | filgotinib.tw. |
| 41 | 9 or 10 or 11 or 12 or 13 or 14 or 15 or 16 or 17 or 18 or 19 or 20 or 21 or 22 or 23 or 24 or 25 or 26 or 27 or 28 or 29 or 30 or 31 or 32 or 33 or 34 or 35 or 36 or 37 or 38 or 39 or 40 |
| 42 | 8 and 41 |
| 43 | 42 and "Randomized Controlled Trial".sa_pubt. |

##### *Supplement table 3 Search term Cochrane via Ovid*

| No. | Search term |
| --- | --- |
| 1 | exp arthritis, rheumatoid/ |
| 2 | ((rheumatoid or reumatoid or revmatoid or rheumatic or reumatic or revmatic or rheumat$ or reumat$ or revmarthrit$) adj3 (arthrit$ or artrit$ or diseas$ or condition$ or nodule$)).tw. |
| 3 | (felty$ adj2 syndrome).tw. |
| 4 | (caplan$ adj2 syndrome).tw. |
| 5 | (sjogren$ adj2 syndrome).tw. |
| 6 | (sicca adj2 syndrome).tw. |
| 7 | still$ disease.tw. |
| 8 | 1 or 2 or 3 or 4 or 5 or 6 or 7 |
| 9 | Methotrexate/ |
| 10 | Methotrexate.tw. |
| 11 | amet?opterine.tw. |
| 12 | mexate.tw. |
| 13 | Abitrexate.tw. |
| 14 | A Met?opterine.tw. |
| 15 | Antifolan.tw. |
| 16 | Emt?exate.tw. |
| 17 | Enthexate.tw. |
| 18 | Farmitrexate.tw. |
| 19 | Folex.tw. |
| 20 | Ledertrexate.tw. |
| 21 | Methoblastin.tw. |
| 22 | Methohexate.tw. |
| 23 | Methotrate.tw. |
| 24 | Methylaminopterin.tw. |
| 25 | Metotrexate.tw. |
| 26 | Mtx.tw. |
| 27 | Novatrex.tw. |
| 28 | Rheumatrex.tw. |
| 29 | Etanercept.tw. |
| 30 | Infliximab.tw. |
| 31 | golimumab.tw. |
| 32 | Adalimumab.tw. |
| 33 | tocilizumab.tw. |
| 34 | rituximab.tw. |
| 35 | tofacitinib.tw. |
| 36 | baricitinib.tw. |
| 37 | (certolizumab and pegol).tw. |
| 38 | sarilumab.tw. |
| 39 | abatacept.tw. |
| 40 | filgotinib.tw. |
| 41 | 9 or 10 or 11 or 12 or 13 or 14 or 15 or 16 or 17 or 18 or 19 or 20 or 21 or 22 or 23 or 24 or 25 or 26 or 27 or 28 or 29 or 30 or 31 or 32 or 33 or 34 or 35 or 36 or 37 or 38 or 39 or 40 |
| 42 | 8 and 41 |
| 43 | 42 and "Randomized Controlled Trial".sa_pubt. |

#### **Supplementary 2 Medications**

##### *Supplementary table 4 All treatments included in the systematic review and network meta-analysis*

| **Pharmacologic Category** | **DMARDs type** | **Interventions of interest** | **Dosing regimen** | **Abbreviation** | **Remission** | |
| --- | --- | --- | --- | --- | --- | --- |
|  |  |  |  |  | **24-26** | **48-52** |
| Conventional disease-modifying antirheumatic drugs | csDMARDs | Methotrexate (MTX) +/- csDMARDs | Oral or parenteral methotrexate monotherapy parenteral methotrexate (subcutaneous or intra-muscular) alone or in combination with csDMARDs other than MTX (i.e. hydroxychloroquine or sulfasalazin). Given the variability of dosing in clinical practice, no dose restrictions were applied to csDMARDs. | MTX | • | • |
| - | - | Placebo | Non-steroidal anti-inflammatory drugs (NSAIDs) and, if necessary, glucocorticoids at a dose of no more than 10 mg/day prednisone or equivalent. | PBO | • | • |
| Anti-TNF-alpha | Originator bDMARDs | Adalimumab | 40 mg subcutaneously every 2 weeks* | ADA (40 mg) | - | - |
|  |  | Adalimumab + MTX | 20 mg subcutaneously every week* | ADA (20 mg) + MTX |  |  |
|  |  | Adalimumab + MTX | 40 mg subcutaneously every 2 weeks | ADA (40 mg) + MTX | • | • |
|  |  | Certolizumab pegol + MTX | 200 mg subcutaneously every 2 weeks after initial dosing of 400 mg subcutaneously at 0, 2, and 4 weeks | CTZ (400 mg) + MTX | • | • |
|  |  | Etanercept + MTX | 25 mg subcutaneously twice weekly | ETA (25 mg) + MTX | • | • |
|  |  | Etanercept | 50 mg subcutaneously every week | ETA (50 mg) | • | - |
|  |  | Etanercept + MTX | 50 mg subcutaneously every week | ETA (50 mg) + MTX | • | - |
|  |  | Golimumab | 100 mg subcutaneously every 4 weeks | GOL (100 mg) | • | - |
|  |  | Golimumab + MTX | 50 mg subcutaneously every 4 weeks | GOL (50 mg) + MTX | • | - |
|  |  | Golimumab + MTX | 100 mg subcutaneously every 4 weeks | GOL (100 mg) + MTX | • | - |
|  |  | Infliximab + MTX | 3 mg/kg intravenously every 8 weeks after initial dosing at 0, 2 and 6 weeks | IFX (3 kg/mg) + MTX | • | • |
|  | Biosimilar bDMARDs | ABP 501 (ADA) + MTX | 40 mg subcutaneously every two weeks | ABP 501 (40 mg) + MTX | • | - |
|  |  | SB5 (ADA) + MTX | 40 mg subcutaneously every two weeks | SB5 40 mg) + MTX | • | - |
|  |  | LBEC0101 (ETA) + MTX | 50 mg subcutaneously every week | LBEC0101 (50 mg) +MTX | • | - |
|  |  | HD203 (ETA) + MTX | 25 mg subcutaneously twice weekly | HD203 (25 mg) + MTX | • | - |
|  |  | SB4 (ETA) + MTX | 50 mg subcutaneously every week | SB4 (50 mg) + MTX | • | - |
|  |  | CT-P13 (IFX) + MTX | 3 mg/kg intravenously every 8 weeks after initial dosing at 0, 2 and 6 weeks | CT-P13 (3 mg/kg) + MTX | • | - |
|  |  | PF-06438179/GP1111 (IFX) + MTX | 3 mg/kg intravenously every 8 weeks after initial dosing at 0, 2 and 6 weeks | PF-06438179/GP1111 (3 mg/kg) + MTX | • | - |
|  |  | SB2 (IFX) + MTX | 3 mg/kg intravenously every 8 weeks after initial dosing at 0, 2 and 6 weeks | SB2 (3 mg/kg) + MTX | • | • |
| Anti-IL-6 | Originator bDMARDs | Tocilizumab + MTX | 4 mg/kg every 4 weeks intravenously | TCZ (4 mg/kg) + MTX | • | • |
|  |  | Tocilizumab | 8 mg/kg every 4 weeks intravenously | TCZ (8 mg/kg) | • | • |
|  |  | Tocilizumab + MTX | 8 mg/kg every 4 weeks intravenously | TCZ (8 mg/kg) + MTX | • | • |
|  |  | Sarilumab^†^ | 200 mg every 2 weeks | SAR (200 mg) | - | - |
| Anti-CD20 | Originator bDMARDs | Rituximab + MTX | 1000 mg IV every 2 weeks for 2 doses | RTX (1000 mg) + MTX | • | • |
|  |  | Rituximab + MTX | 500 mg IV every 2 weeks for 2 doses | RTX (500 mg) + MTX | • | • |
|  | Biosimilar bDMARDs | CT-P10 + MTX | 1000 mg intravenously every 2 weeks for 2 doses on days 1 and 15 | CT-P10 (1000 mg) + MTX | • | • |
| Selective T-Cell Co-stimulation Blocker | Originator bDMARDs | Abatacept + MTX | <60 kg: 500 mg; ≥60 to ≤100 kg: 750 mg; >100 kg: 1000 mg intravenously on day 1, at weeks 2 and 4 and every 4 weeks thereafter | ABC (10 mg/kg) + MTX | • | - |
|  |  | Abatacept + MTX | 125 mg subcutaneously once weekly | ABC (125 mg) + MTX | • | • |
| JAK inhibitor | tsDMARDs | Tofacitinib | 5 mg orally twice daily | TOF (5 mg) | • | • |
|  |  | Tofacitinib + MTX | 5 mg orally twice daily | TOF (5 mg) + MTX | • | • |
|  |  | Tofacitinib | 10 mg orally twice daily | TOF (10 mg) | • | • |
|  |  | Tofacitinib + MTX | 10 mg orally twice daily | TOF (10 mg) + MTX | • | • |
|  |  | Baricitinib + MTX +/- csDMARDs^#^ | 2 mg orally once daily | BAR (2 mg) + MTX +/- csDMARDs | • | - |
|  |  | Baricitinib | 4 mg orally once daily | BAR (4 mg) | • | • |
|  |  | Baricitinib + MTX +/- csDMARDs | 4 mg orally once daily | BAR (4 mg) + MTX +/- csDMARDs | • | • |
|  |  | Filgotinib + MTX + csDMARDs* | 100 mg once daily | FIL (100 mg) + MTX + csDMARD | - | - |
|  |  | Filgotinib + MTX + csDMARD* | 200 mg once daily | FIL (200 mg) + MTX + csDMARD | - | - |

†Not included in network meta-analysis; *Not included in analysis for safety outcomes.

### **Results Supplement**

#### **Supplementary 3 Risk of bias assessments**

*Supplement Table 5 Risk of bias assessment for remission (DAS28) and safety on study level*

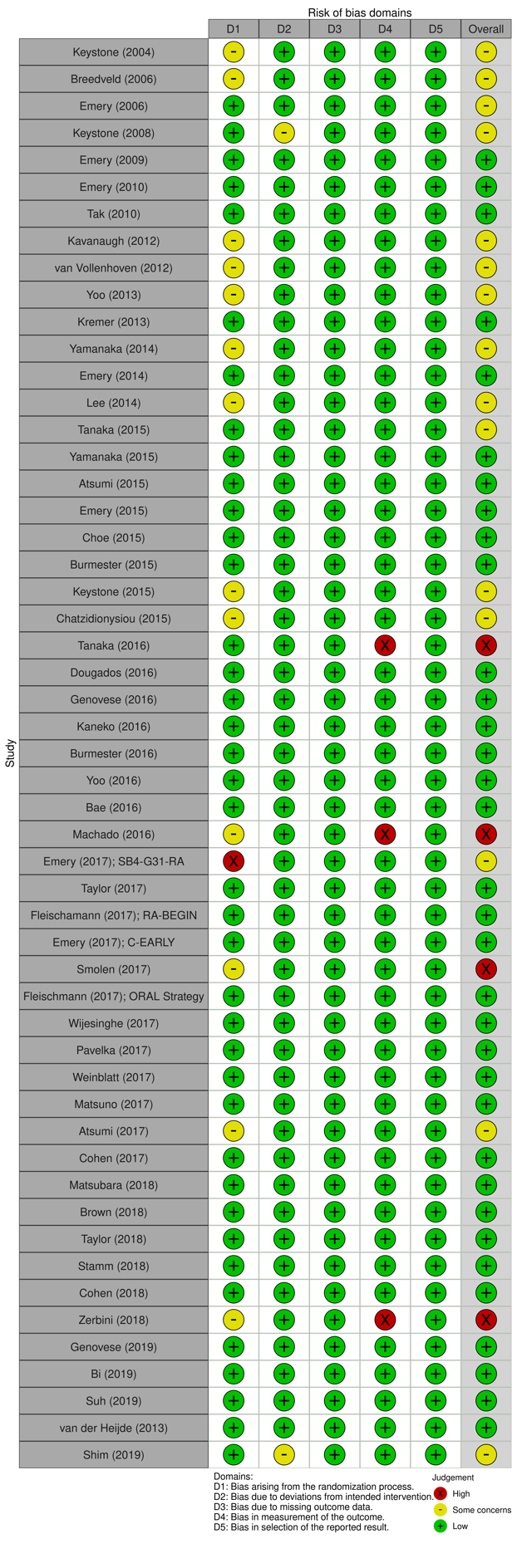

#### **Supplementary 4 Consistency assessment**

##### *Supplement table 6 Assessing inconsistency of efficacy outcomes*

| Outcomes | Timepoint | Inconsistency: p-values based on global test for inconsistency in network meta-analysis |
| --- | --- | --- |
| Remission | 24-26 weeks | 0.323 |
|  | 48-52 weeks | 0.006 |

##### *Supplement table 7 Assessing inconsistency of safety outcomes*

| Outcomes | Timepoint | Inconsistency: p-values based on global test for inconsistency in network meta-analysis |
| --- | --- | --- |
| Serious infection | 24-26 weeks | 0.401 |
|  | 48-52 weeks | 0.049 |
| Tuberculosis | 24-26 weeks | No source of inconsistency |
|  | 48-52 weeks | 0.846 |
| Herpes zoster | 24-26 weeks | 0.985 |
|  | 48-52 weeks | 0.611 |
| Cancer | 24-26 weeks | 0.278 |
|  | 48-52 weeks | 0.718 |
| Cardiovascular events | 24-26 weeks | 0.961 |
|  | 48-52 weeks | 0.397 |

**Supplementary 5**

*Supplement table 8 Remission at 24-26 weeks using inconsistency model*

| Medication (dose) | Consistency model  RR (95% CI) | Inconsistency model  RR (95% CI) |
| --- | --- | --- |
| Tofacitinib (10 mg) + MTX | **4.65** (2.98-7.27) | **11.81** (1.57-88.71) |
| Abatacept (10 mg/kg) + MTX | **3.74** (2.60-5.38) | **3.74** (2.30-6.08) |
| Tocilizumab (8 mg/kg) + MTX | **3.06** (2.27-4.12) | **3.00** (1.93-4.68) |
| Tofacitinib (10 mg) | **3.05** (2.03-4.57) | **2.80** (1.50-5.23) |
| Golimumab (50 mg) + MTX | **2.96** (2.03-4.32) | **5.12** (2.11-12.40) |
| Etanercept biosimilar LBEC0101 + MTX | **3.07** (1.87-5.05) | **3.11** (1.65-8.79) |
| Etanercept biosimilar SB4 + MTX | **2.95** (1.75-4.98) | **2.99** (1.55-5.77) |
| Etanercept (50 mg) + MTX | **2.85** (1.96-4.16) | **2.89** (1.85-4.50) |
| Adalimumab biosimilar SB5 (40 mg) + MTX | **2.81** (1.85-4.26) | **2.44** (1.27-4.68) |
| Baricitinib (4 mg) + MTX +/- csDMARDs | **2.71** (2.13-3.44) | **3.60** (2.10-6.16) |
| Adalimumab (40 mg) + MTX | **2.58** (2.03-3.28) | **2.25** (1.43-3.53) |
| Infliximab biosimilar CT-P13 + MTX | **2.62** (1.25-5.46) | 2.31 (0.96-5.54) |
| Tocilizumab (8 mg/kg) | **2.54** (1.88-3.44) | **2.60** (1.66-4.07) |
| Tofacitinib (5 mg) + MTX | **2.49** (1.70-3.65) | 5.85 (0.74-45.61) |
| Etanercept biosimilar HD203 + MTX | **2.38** (1.39-4.08) | **2.20** (1.07-4.51) |
| Etanercept (50 mg) | **2.37** (1.23-4.53) | **2.40** (1.12-5.14) |
| Infliximab biosimilar PF-06438179/GP1111 + MTX | **2.25** (1.17-4.33) | 1.93 (0.03-3.54) |
| Baricitinib (2 mg) + MTX +/- csDMARDs | **2.26** (1.49-3.43) | **2.77** (1.45-5.27) |
| Etanercept (25 mg) + MTX | **2.23** (1.64-3.04) | **2.06** (1.30-3.28) |
| Golimumab (100 mg) + MTX | **2.19** (1.47-3.27) | **3.20** (1.27-8.09) |
| Adalimumab biosimilar ABP-501 + MTX | **2.22** (1.58-3.12) | **1.93** (1.05-3.54) |
| Tocilizumab (4 mg/kg) + MTX | **2.13** (1.54-2.95) | **2.13** (1.35-3.37) |
| Baricitinib (4 mg) | **2.11** (1.42-3.14) | 1.75 (0.99-3.11) |
| Tofacitinib (5 mg) | **2.08** (1.46-2.97) | 1.88 (0.99-3.57) |
| Golimumab (100 mg) | **1.98** (1.30-3.03) | **1.79** (1.08-2.98) |
| Infliximab (3 mg/kg) + MTX | **1.95** (1.11-3.43) | 1.72 (0.89-3.34) |
| Infliximab biosimilar SB2 + MTX | 1.75 (0.89-3.47) | 1.55 (0.68-3.55) |
| Certolizumab-pegol (400 mg) + MTX | **1.75** (1.44-2.13) | **1.78** (1.30-2.44) |
| Abatacept (125 mg) + MTX | 1.15 (0.26-5.13) | 1.20 (0.25-5.75) |
| Rituximab (1000 mg) + MTX | **1.58** (1.04-2.40) | 1.65 (0.96-2.82) |
| Rituximab (500 mg) + MTX | 1.51 (0.99-2.31) | 1.59 (0.92-2.75) |
| Placebo | **1.45** (1.01-2.08) | 1.40 (0.85-2.31) |
| Rituximab biosimilar CT-P10 + MTX | 1.22 (0.60-2.51) | 1.28 (0.54-3.01) |
| MTX | Comparator | |

RR = relative risk; Significant RR (CI) are in bold.

Abatacept (125 mg) = abatacept 125 mg subcutaneously injection (SC) once weekly; Abatacept (10 mg/kg) = abatacept 10 mg/kg intravenous infusion (IV) every 4 weeks; Adalimumab (20 or 40 mg) = adalimumab 20 or 40 mg SC injection every other week; Adalimumab biosimilar ABP-501+MTX = adalimumab biosimilar 40 mg; Adalimumab biosimilar SB5 (40 mg) = SB5 Adalimumab biosimilar 40 mg every other week SC + MTX; Baricitinib (2 or 4 mg) = baricitinib 2 or 4 mg orally once daily; Certolizumab pegol (400 mg) = certolizumab pegol 400 mg SC every 4 weeks; csDMARDs = sulfasalazine and hydroxychloroquine; Etanercept (25 or 50 mg) = etanercept 25 or 50 mg SC once weekly; Etanercept biosimilar HD203+MTX = etanercept biosimilar 25 mg twice weekly SC; Etanercept biosimilar LBEC0101 + MTX = etanercept biosimilar 50 mg once weekly SC + MTX; Etanercept biosimilar SB4 + MTX = etanercept biosimilar 50 mg SC once weekly + MTX; Filgotinib (200 mg) = filgotinib 200 mg orally once daily; Golimumab (50 or 100 mg) = golimumab 50 or 100 mg SC every 4 weeks; Infliximab (3 mg/kg) = infliximab 3 mg/kg IV every 8 weeks; Infliximab biosimilar CT-P13+MTX = infliximab biosimilar 3 mg/kg + MTX; Infliximab biosimilar PF-06438179/GP1111+MTX = infliximab biosimilar 3 mg/kg + MTX; Infliximab biosimilar SB2+MTX = infliximab biosimilar 3 mg/kg + MTX; MTX = methotrexate; Rituximab (500 or 1,000 mg) = rituximab 500 or 1,000 mg IV 2 courses (15 days apart) every 24 weeks; Rituximab biosimilar CT-P10+MTX = biosimilar rituximab 1000 mg + MTX; Tocilizumab (4 or 8 mg/kg) = tocilizumab 4 or 8 mg/kg IV every 4 weeks; Tofacitinib (5 or 10 mg) = tofacitinib 5 or 10 mg orally twice daily.

*Supplement table 9 Remission at 48-52 weeks using inconsistency model*

| Medication (dose) | Consistency model  RR (95% CI) | Inconsistency model  RR (95% CI) |
| --- | --- | --- |
| Tocilizumab (8 mg/kg) + MTX | **2.41** (1.67-3.48) | **2.52** (1.94-3.28) |
| Rituximab (1000 mg) + MTX | **2.38** (1.50-3.79) | **2.38** (1.65-3.45) |
| Etanercept (25 mg) + MTX | **2.32** (1.50-3.61) | **1.93** (1.14-3.25) |
| Infliximab biosimilar SB2 (3 mg/kg) + MTX | **2.25** (1.11-4.55) | **1.94** (1.05-3.56) |
| Tocilizumab (8 mg/kg) | **2.13** (1.47-3.09) | **2.03** (1.54-2.68) |
| Infliximab (3 mg/kg) + MTX | **2.03** (1.18-3.49) | **1.75** (1.06-2.88) |
| Abatacept (125 mg) + MTX | 2.33 (0.32-16.94) | 2.33 (0.33-16.30) |
| Rituximab (500 mg) + MTX | **1.92** (1.19-3.10) | **1.92** (1.31-2.83) |
| Tofacitinib (10 mg) + MTX | 1.72 (0.84-3.52) | 1.24 (0.68-2.24) |
| Tocilizumab (4mg/kg) + MTX | **1.74** (1.17-2.60) | **1.74** (1.31-2.32) |
| Rituximab biosimilar CT-P10 + MTX | **1.71** (0.92-3.20) | **1.71** (1.02-2.88) |
| Baricitinib (4 mg) + MTX +/- csDMARDs | **1.67** (1.06-2.61) | **1.67** (1.17-2.37) |
| Certolizumab pegol (400 mg) + MTX | **1.56** (1.26-1.93) | **1.56** (1.36-1.80) |
| Tofacitinib (10 mg) | 1.51 (0.93-2.43) | **1.96** (1.27-3.03) |
| Adalimumab (40 mg) + MTX | 1.24 (0.87-1.78) | 1.01 (0.76-1.33) |
| Placebo | 1.21 (0.71-2.07) | 1.27 (0.71-2.27) |
| Baricitinib (4 mg) | 1.22 (0.74-2.01) | 1.22 (0.81-1.84) |
| Tofacitinib (5 mg) + MTX | 1.20 (0.71-2.01) | 0.86 (0.56-1.33) |
| Tofacitinib (5 mg) | 1.08 (0.71-1.63) | **1.57** (1.01-2.46) |
| MTX | Comparator | |

Significant RR (CI) are in bold.

Abatacept (125 mg) = abatacept 125 mg subcutaneously injection (SC) once weekly; Abatacept (10 mg/kg) = abatacept 10 mg/kg intravenous infusion (IV) every 4 weeks; Adalimumab (20 or 40 mg) = adalimumab 20 or 40 mg SC injection every other week; Baricitinib (2 or 4 mg) = baricitinib 2 or 4 mg orally once daily; Certolizumab pegol (400 mg) = certolizumab pegol 400 mg SC every 4 weeks; csDMARDs = sulfasalazine and hydroxychloroquine; Filgotinib (200 mg) = filgotinib 200 mg orally once daily; Etanercept (25 or 50 mg) = etanercept 25 or 50 mg SC once weekly; Golimumab (50 or 100 mg) = golimumab 50 or 100 mg SC every 4 weeks; Infliximab (3 mg/kg) = infliximab 3 mg/kg IV every 8 weeks; Infliximab biosimilar SB2+MTX = infliximab biosimilar 3 mg/kg + MTX; MTX = methotrexate; Rituximab (500 or 1,000 mg) = rituximab 500 or 1,000 mg IV 2 courses (15 days apart) every 24 weeks; Rituximab biosimilar CT-P10 + MTX = biosimilar rituximab 1000 mg + MTX; Tocilizumab (4 or 8 mg/kg) = tocilizumab 4 or 8 mg/kg IV every 4 weeks; Tofacitinib ( 5 or 10 mg) = tofacitinib 5 or 10 mg orally twice daily.

#### **Supplementary 5 Comparative effect of DMARDs**

#### **Efficacy outcomes**

##### *Supplementary table 10 league table remission at 24-26 weeks and 48-52 weeks*

| **RR (CI) of remission at 48-52 weeks** | **RR (CI) of remission at 24-26 weeks** | | | | | | | | | | | | | | | | | | | | | | | | | | | | | | | | | |
| --- | --- | --- | --- | --- | --- | --- | --- | --- | --- | --- | --- | --- | --- | --- | --- | --- | --- | --- | --- | --- | --- | --- | --- | --- | --- | --- | --- | --- | --- | --- | --- | --- | --- | --- |
|  | MTX | **2.22** | **2.81** | 1.22 | **2.25** | **2.62** | 1.75 | **2.38** | **3.07** | **2.95** | **4.65** | **2.49** | **3.05** | **2.08** | **2.71** | **2.26** | **2.11** | **1.58** | 1.51 | **3.06** | **2.54** | **2.13** | **1.95** | **1.98** | **2.19** | **2.96** | **2.37** | **2.85** | **2.23** | **1.75** | **2.58** | **1.15** | **3.74** | **1.45** |
|  |  | (1.58-3.12) | (1.85-4.26) | (0.60-2.51) | (1.17-4.33) | (1.25-5.46) | (0.89-3.47) | (1.39-4.08) | (1.87-5.05) | (1.75-4.98) | (2.98-7.27) | (1.70-3.65) | (2.03-4.57) | (1.46-2.97) | (2.13-3.44) | (1.49-3.43) | (1.42-3.14) | (1.04-2.40) | (0.99-2.31) | (2.27-4.12) | (1.88-3.44) | (1.54-2.95) | (1.11-3.43) | (1.30-3.03) | (1.47-3.27) | (2.03-4.32) | (1.23-4.53) | (1.96-4.16) | (1.64-3.04) | (1.44-2.13) | (2.03-3.28) | (0.26-5.13) | (2.60-5.38) | (1.01-2.08) |
|  | N/A | MTX + ABP501 (40 mg) | 1.26 | 0.55 | 1.01 | 1.18 | 0.79 | 1.07 | 1.39 | 1.33 | **2.1** | 1.12 | 1.37 | 0.94 | 1.22 | 1.02 | 0.95 | 0.71 | 0.68 | 1.38 | 1.15 | 0.96 | 0.88 | 0.89 | 0.99 | 1.33 | 1.07 | 1.29 | 1.01 | 0.79 | 1.16 | 0.52 | **1.69** | 0.65 |
|  |  |  | (0.83-1.93) | (0.25-1.22) | (0.49-2.12) | (0.52-2.66) | (0.37-1.70) | (0.57-2.03) | (0.76-2.53) | (0.71-2.48) | (1.32-3.34) | (0.75-1.68) | (0.86-2.20) | (0.62-1.41) | (0.86-1.73) | (0.62-1.68) | (0.58-1.55) | (0.41-1.22) | (0.40-1.18) | (0.88-2.17) | (0.73-1.81) | (0.60-1.54) | (0.45-1.70) | (0.52-1.54) | (0.58-1.67) | (0.80-2.22) | (0.51-2.22) | (0.77-2.14) | (0.64-1.59) | (0.53-1.17) | (0.91-1.49) | (0.11-2.40) | (1.03-2.78) | (0.40-1.07) |
|  | N/A | N/A | MTX + SB5  (40 mg) | **0.44** | 0.8 | 0.93 | 0.63 | 0.85 | 1.1 | 1.05 | 1.66 | 0.89 | 1.09 | 0.74 | 0.96 | 0.81 | 0.75 | 0.56 | **0.54** | 1.09 | 0.91 | 0.76 | 0.69 | 0.71 | 0.78 | 1.06 | 0.84 | 1.02 | 0.8 | **0.62** | 0.92 | 0.41 | 1.33 | 0.52 |
|  |  |  |  | (0.19-1.00) | (0.37-1.74) | (0.40-2.17) | (0.28-1.39) | (0.43-1.68) | (0.57-2.10) | (0.54-2.05) | (0.98-2.80) | (0.56-1.42) | (0.64-1.84) | (0.46-1.19) | (0.63-1.48) | (0.46-1.40) | (0.44-1.30) | (0.31-1.02) | (0.30-0.98) | (0.65-1.82) | (0.54-1.52) | (0.45-1.29) | (0.34-1.40) | (0.39-1.28) | (0.44-1.39) | (0.60-1.85) | (0.39-1.83) | (0.58-1.78) | (0.47-1.34) | (0.39-0.99) | (0.65-1.30) | (0.09-1.94) | (0.77-2.32) | (0.30-0.90) |
|  | **1.71** | N/A | N/A | MTX + CT-P10 (1000 mg) | 1.84 | 2.14 | 1.43 | 1.95 | **2.51** | 2.41 | **3.8** | 2.04 | **2.49** | 1.7 | **2.21** | 1.85 | 1.72 | 1.29 | 1.24 | **2.5** | 2.08 | 1.74 | 1.59 | 1.62 | 1.79 | **2.42** | 1.93 | **2.33** | 1.82 | 1.43 | 2.11 | 0.94 | **3.06** | 1.18 |
|  | (1.02-2.88) |  |  |  | (0.70-4.86) | (0.76-5.98) | (0.53-3.86) | (0.79-4.78) | (1.05-6.02) | (0.99-5.86) | (1.63-8.86) | (0.90-4.60) | (1.09-5.68) | (0.76-3.79) | (1.04-4.72) | (0.81-4.24) | (0.76-3.92) | (0.72-2.31) | (0.62-2.45) | (1.15-5.44) | (0.95-4.53) | (0.79-3.83) | (0.64-3.97) | (0.70-3.73) | (0.79-4.07) | (1.07-5.45) | (0.73-5.10) | (1.03-5.25) | (0.83-3.99) | (0.68-3.01) | (0.99-4.50) | (0.20-4.42) | (1.37-6.84) | (0.53-2.65) |
|  | N/A | N/A | N/A | N/A | MTX + PF-06438179/GP1111 (3mg) | 1.16 | 0.78 | 1.06 | 1.37 | 1.31 | 2.07 | 1.11 | 1.35 | 0.92 | 1.2 | 1.01 | 0.94 | 0.7 | 0.67 | 1.36 | 1.13 | 0.95 | 0.87 | 0.88 | 0.97 | 1.31 | 1.05 | 1.27 | 0.99 | 0.78 | 1.15 | 0.51 | 1.66 | 0.64 |
|  |  |  |  |  |  | (0.65-2.07) | (0.47-1.29) | (0.46-2.45) | (0.60-3.10) | (0.57-3.03) | (0.94-4.56) | (0.52-2.36) | (0.63-2.92) | (0.44-1.95) | (0.60-2.41) | (0.46-2.18) | (0.44-2.02) | (0.32-1.52) | (0.31-1.47) | (0.66-2.79) | (0.55-2.32) | (0.46-1.96) | (0.62-1.21) | (0.40-1.92) | (0.45-2.09) | (0.62-2.80) | (0.42-2.64) | (0.60-2.70) | (0.48-2.03) | (0.39-1.54) | (0.57-2.30) | (0.10-2.61) | (0.79-3.51) | (0.31-1.34) |
|  | N/A | N/A | N/A | N/A | N/A | MTX + CT-P13  (3 mg/kg) | 0.67 | 0.91 | 1.18 | 1.13 | 1.78 | 0.95 | 1.16 | 0.8 | 1.03 | 0.87 | 0.81 | 0.6 | 0.58 | 1.17 | 0.97 | 0.82 | 0.75 | 0.76 | 0.84 | 1.13 | 0.9 | 1.09 | 0.85 | 0.67 | 0.99 | 0.44 | 1.43 | 0.55 |
|  |  |  |  |  |  |  | (0.36-1.23) | (0.37-2.25) | (0.48-2.86) | (0.46-2.78) | (0.75-4.21) | (0.42-2.19) | (0.50-2.70) | (0.35-1.80) | (0.48-2.25) | (0.37-2.02) | (0.35-1.86) | (0.26-1.41) | (0.25-1.35) | (0.53-2.59) | (0.44-2.15) | (0.36-1.82) | (0.46-1.20) | (0.32-1.77) | (0.36-1.94) | (0.49-2.59) | (0.34-2.42) | (0.48-2.49) | (0.39-1.88) | (0.31-1.43) | (0.46-2.14) | (0.08-2.33) | (0.63-3.25) | (0.25-1.25) |
|  | **1.94** | N/A | N/A | 1.13 | N/A | N/A | MTX + SB2  (3mg/kg) | 1.36 | 1.75 | 1.68 | **2.65** | 1.42 | 1.74 | 1.19 | 1.54 | 1.29 | 1.2 | 0.9 | 0.86 | 1.74 | 1.45 | 1.22 | 1.11 | 1.13 | 1.25 | 1.69 | 1.35 | 1.63 | 1.27 | 1 | 1.47 | 0.66 | 2.13 | 0.83 |
|  | (1.05-3.56) |  |  | (0.51-2.52) |  |  |  | (0.57-3.22) | (0.75-4.08) | (0.71-3.97) | (1.17-6.00) | (0.65-3.11) | (0.79-3.84) | (0.55-2.56) | (0.75-3.18) | (0.58-2.87) | (0.55-2.65) | (0.40-2.00) | (0.39-1.93) | (0.83-3.67) | (0.69-3.06) | (0.57-2.59) | (0.76-1.63) | (0.51-2.52) | (0.57-2.75) | (0.77-3.68) | (0.53-3.46) | (0.75-3.55) | (0.61-2.67) | (0.49-2.03) | (0.71-3.03) | (0.13-3.39) | (0.99-4.62) | (0.39-1.77) |
|  | N/A | N/A | N/A | N/A | N/A | N/A | N/A | MTX + HD203  (25 mg) | 1.29 | 1.24 | 1.95 | 1.05 | 1.28 | 0.87 | 1.14 | 0.95 | 0.89 | 0.66 | 0.64 | 1.28 | 1.07 | 0.9 | 0.82 | 0.83 | 0.92 | 1.24 | 0.99 | 1.2 | 0.94 | 0.74 | 1.08 | 0.48 | 1.57 | 0.61 |
|  |  |  |  |  |  |  |  |  | (0.62-2.69) | (0.58-2.63) | (0.97-3.93) | (0.54-2.03) | (0.65-2.51) | (0.46-1.67) | (0.63-2.05) | (0.48-1.88) | (0.45-1.73) | (0.33-1.31) | (0.32-1.26) | (0.69-2.38) | (0.58-1.98) | (0.48-1.68) | (0.38-1.77) | (0.42-1.65) | (0.47-1.80) | (0.64-2.40) | (0.43-2.31) | (0.62-2.31) | (0.60-1.46) | (0.41-1.31) | (0.60-1.96) | (0.10-2.37) | (0.82-3.01) | (0.37-1.01) |
|  | N/A | N/A | N/A | N/A | N/A | N/A | N/A | N/A | MTX + LBEC0101  (50 mg) | 0.96 | 1.51 | 0.81 | 0.99 | 0.68 | 0.88 | 0.74 | 0.69 | **0.51** | **0.49** | 0.99 | 0.83 | 0.69 | 0.63 | 0.64 | 0.71 | 0.96 | 0.77 | 0.93 | 0.73 | 0.57 | 0.84 | 0.38 | 1.22 | 0.47 |
|  |  |  |  |  |  |  |  |  |  | (0.59-1.56) | (0.78-2.95) | (0.43-1.52) | (0.52-1.88) | (0.37-1.25) | (0.51-1.53) | (0.39-1.41) | (0.36-1.30) | (0.27-0.98) | (0.26-0.95) | (0.56-1.78) | (0.46-1.48) | (0.38-1.25) | (0.30-1.34) | (0.34-1.24) | (0.38-1.35) | (0.52-1.80) | (0.41-1.43) | (0.67-1.28) | (0.40-1.30) | (0.33-0.97) | (0.48-1.46) | (0.08-1.81) | (0.66-2.25) | (0.26-0.87) |
|  | N/A | N/A | N/A | N/A | N/A | N/A | N/A | N/A | N/A | MTX + SB4  (50 mg) | 1.58 | 0.85 | 1.03 | 0.71 | 0.92 | 0.77 | 0.72 | 0.53 | 0.51 | 1.04 | 0.86 | 0.72 | 0.66 | 0.67 | 0.74 | 1 | 0.8 | 0.97 | 0.76 | 0.59 | 0.88 | 0.39 | 1.27 | **0.49** |
|  |  |  |  |  |  |  |  |  |  |  | (0.79-3.14) | (0.44-1.62) | (0.53-2.00) | (0.37-1.33) | (0.52-1.63) | (0.39-1.50) | (0.37-1.38) | (0.27-1.04) | (0.26-1.01) | (0.57-1.89) | (0.47-1.58) | (0.39-1.34) | (0.31-1.43) | (0.34-1.32) | (0.38-1.43) | (0.53-1.91) | (0.42-1.52) | (0.67-1.39) | (0.41-1.39) | (0.34-1.04) | (0.49-1.56) | (0.08-1.90) | (0.67-2.40) | (0.26-0.93) |
|  | 1.24 | N/A | N/A | 0.72 | N/A | N/A | 0.64 | N/A | N/A | N/A | MTX + TOF  (10 mg) | **0.54** | 0.65 | **0.45** | **0.58** | **0.49** | **0.45** | **0.34** | **0.33** | 0.66 | **0.55** | **0.46** | **0.42** | **0.43** | **0.47** | 0.64 | 0.51 | 0.61 | **0.48** | **0.38** | **0.56** | 0.25 | 0.8 | **0.31** |
|  | (0.68-2.24) |  |  | (0.33-1.59) |  |  | (0.27-1.50) |  |  |  |  | (0.40-0.71) | (0.39-1.09) | (0.29-0.70) | (0.37-0.92) | (0.27-0.87) | (0.26-0.80) | (0.18-0.63) | (0.18-0.60) | (0.38-1.12) | (0.32-0.94) | (0.26-0.80) | (0.20-0.86) | (0.23-0.79) | (0.26-0.86) | (0.35-1.14) | (0.23-1.12) | (0.34-1.10) | (0.28-0.83) | (0.23-0.61) | (0.37-0.82) | (0.05-1.18) | (0.45-1.43) | (0.18-0.55) |
|  | 0.86 | N/A | N/A | 0.5 | N/A | N/A | 0.45 | N/A | N/A | N/A | 0.7 | MTX + TOF  (5 mg) | 1.22 | 0.83 | 1.09 | 0.91 | 0.85 | 0.63 | 0.61 | 1.23 | 1.02 | 0.85 | 0.78 | 0.79 | 0.88 | 1.19 | 0.95 | 1.14 | 0.89 | 0.7 | 1.04 | 0.46 | 1.5 | **0.58** |
|  | (0.56-1.33) |  |  | (0.26-0.99) |  |  | (0.21-0.94) |  |  |  | (0.46-1.05) |  | (0.78-1.91) | (0.58-1.20) | (0.73-1.62) | (0.53-1.54) | (0.50-1.43) | (0.36-1.11) | (0.34-1.07) | (0.76-1.99) | (0.63-1.66) | (0.52-1.41) | (0.40-1.54) | (0.45-1.41) | (0.51-1.53) | (0.69-2.03) | (0.45-2.02) | (0.67-1.95) | (0.55-1.46) | (0.46-1.08) | (0.75-1.43) | (0.10-2.16) | (0.89-2.54) | (0.34-0.98) |
|  | **1.96** | N/A | N/A | 1.15 | N/A | N/A | 1.01 | N/A | N/A | N/A | 1.59 | **2.28** | TOF  (10 mg) | **0.68** | 0.89 | 0.74 | 0.69 | **0.52** | **0.5** | 1 | 0.83 | 0.7 | 0.64 | 0.65 | 0.72 | 0.97 | 0.78 | 0.94 | 0.73 | **0.57** | 0.85 | 0.38 | 1.23 | **0.48** |
|  | (1.27-3.03) |  |  | (0.58-2.26) |  |  | (0.48-2.14) |  |  |  | (0.76-3.32) | (1.24-4.19) |  | (0.51-0.92) | (0.57-1.38) | (0.42-1.30) | (0.40-1.20) | (0.29-0.93) | (0.28-0.89) | (0.61-1.66) | (0.50-1.38) | (0.42-1.18) | (0.32-1.28) | (0.36-1.17) | (0.41-1.27) | (0.56-1.69) | (0.36-1.67) | (0.54-1.63) | (0.44-1.22) | (0.37-0.90) | (0.57-1.26) | (0.08-1.78) | (0.71-2.12) | (0.28-0.82) |
|  | **1.57** | N/A | N/A | 0.92 | N/A | N/A | 0.81 | N/A | N/A | N/A | 1.27 | 1.83 | 0.8 | TOF  (5 mg) | 1.3 | 1.09 | 1.01 | 0.76 | 0.73 | 1.47 | 1.22 | 1.02 | 0.94 | 0.95 | 1.05 | 1.42 | 1.14 | 1.37 | 1.07 | 0.84 | 1.24 | 0.55 | **1.8** | 0.7 |
|  | (1.01-2.46) |  |  | (0.46-1.83) |  |  | (0.38-1.73) |  |  |  | (0.60-2.68) | (0.98-3.40) | (0.61-1.06) |  | (0.88-1.91) | (0.65-1.83) | (0.61-1.69) | (0.44-1.31) | (0.42-1.26) | (0.92-2.34) | (0.77-1.95) | (0.63-1.66) | (0.48-1.82) | (0.55-1.66) | (0.62-1.80) | (0.85-2.39) | (0.54-2.39) | (0.8-2.30) | (0.67-1.72) | (0.56-1.26) | (0.90-1.72) | (0.12-2.57) | (1.08-2.99) | (0.42-1.16) |
|  | **1.67** | N/A | N/A | 0.97 | N/A | N/A | 0.86 | N/A | N/A | N/A | 1.35 | **1.93** | 0.85 | 1.06 | BAR 4 mg + MTX (+/- csDMARDs) | 0.84 | 0.78 | **0.58** | 0.56 | 1.13 | 0.94 | 0.79 | 0.72 | 0.73 | 0.81 | 1.09 | 0.87 | 1.05 | 0.82 | **0.65** | 0.95 | 0.43 | 1.38 | **0.54** |
|  | (1.17-2.37) |  |  | (0.52-1.83) |  |  | (0.43-1.74) |  |  |  | (0.67-2.69) | (1.11-3.37) | (0.48-1.48) | (0.60-1.87) |  | (0.58-1.21) | (0.54-1.12) | (0.36-0.94) | (0.34-0.91) | (0.77-1.66) | (0.64-1.38) | (0.53-1.18) | (0.39-1.33) | (0.45-1.19) | (0.51-1.29) | (0.70-1.71) | (0.44-1.75) | (0.67-1.65) | (0.56-1.22) | (0.47-0.88) | (0.74-1.23) | (0.09-1.93) | (0.90-2.14) | (0.35-0.83) |
|  | N/A | N/A | N/A | N/A | N/A | N/A | N/A | N/A | N/A | N/A | N/A | N/A | N/A | N/A | N/A | BAR (2 mg) + MTX (+/- csDMARDs) | 0.93 | 0.7 | 0.67 | 1.35 | 1.12 | 0.94 | 0.86 | 0.88 | 0.97 | 1.31 | 1.04 | 1.26 | 0.99 | 0.77 | 1.14 | 0.51 | 1.65 | 0.64 |
|  |  |  |  |  |  |  |  |  |  |  |  |  |  |  |  |  | (0.56-1.56) | (0.39-1.26) | (0.37-1.21) | (0.81-2.25) | (0.67-1.88) | (0.56-1.59) | (0.43-1.74) | (0.48-1.59) | (0.54-1.72) | (0.75-2.29) | (0.48-2.26) | (0.72-2.21) | (0.59-1.65) | (0.49-1.22) | (0.74-1.76) | (0.11-2.40) | (0.95-2.87) | (0.37-1.11) |
|  | 1.22 | N/A | N/A | 0.71 | N/A | N/A | 0.63 | N/A | N/A | N/A | 0.99 | 1.42 | 0.62 | 0.78 | 0.73 | N/A | BAR 4 mg | 0.75 | 0.72 | 1.45 | 1.2 | 1.01 | 0.92 | 0.94 | 1.04 | 1.4 | 1.12 | 1.35 | 1.06 | 0.83 | 1.22 | 0.55 | **1.77** | 0.69 |
|  | (0.81-1.84) |  |  | (0.37-1.39) |  |  | (0.30-1.32) |  |  |  | (0.48-2.04) | (0.78-2.57) | (0.34-1.13) | (0.42-1.43) | (0.51-1.05) |  |  | (0.42-1.33) | (0.40-1.28) | (0.88-2.38) | (0.73-1.99) | (0.60-1.69) | (0.46-1.84) | (0.53-1.68) | (0.59-1.83) | (0.81-2.43) | (0.52-2.40) | (0.78-2.34) | (0.64-1.75) | (0.53-1.29) | (0.80-1.87) | (0.12-2.56) | (1.04-3.04) | (0.40-1.18) |
|  | **2.38** | N/A | N/A | 1.39 | N/A | N/A | 1.23 | N/A | N/A | N/A | 1.93 | **2.77** | 1.21 | 1.51 | 1.43 | **1.95** | N/A | MTX + RTX (1000 mg) | 0.96 | **1.94** | 1.61 | 1.35 | 1.24 | 1.26 | 1.39 | 1.88 | 1.5 | **1.81** | 1.42 | 1.11 | **1.64** | 0.73 | **2.38** | 0.92 |
|  | (1.65-3.45) |  |  | (0.97-2.01) |  |  | (0.60-2.51) |  |  |  | (0.96-3.89) | (1.57-4.88) | (0.69-2.15) | (0.85-2.71) | (0.86-2.39) | (1.12-3.39) |  |  | (0.67-1.37) | (1.16-3.25) | (0.96-2.70) | (0.80-2.30) | (0.61-2.50) | (0.69-2.28) | (0.78-2.48) | (1.07-3.31) | (0.69-3.26) | (1.03-3.18) | (0.84-2.38) | (0.70-1.77) | (1.01-2.66) | (0.17-3.06) | (1.36-4.14) | (0.53-1.60) |
|  | **1.92** | N/A | N/A | 1.12 | N/A | N/A | 0.99 | N/A | N/A | N/A | 1.56 | **2.23** | 0.98 | 1.22 | 1.15 | 1.57 | 0.81 | N/A | MTX + RTX (500 mg) | **2.02** | 1.68 | 1.41 | 1.29 | 1.31 | 1.45 | **1.96** | 1.56 | **1.88** | 1.48 | 1.16 | **1.71** | 0.76 | **2.47** | 0.96 |
|  | (1.31-2.83) |  |  | (0.71-1.78) |  |  | (0.48-2.04) |  |  |  | (0.77-3.16) | (1.25-3.98) | (0.55-1.75) | (0.68-2.21) | (0.68-1.95) | (0.89-2.77) | (0.61-1.07) |  |  | (1.20-3.40) | (1.00-2.83) | (0.83-2.40) | (0.64-2.61) | (0.72-2.39) | (0.81-2.60) | (1.11-3.46) | (0.72-3.40) | (1.07-3.33) | (0.87-2.49) | (0.72-1.85) | (1.05-2.78) | (0.17-3.34) | (1.42-4.33) | (0.55-1.67) |
|  | **2.52** | N/A | N/A | 1.47 | N/A | N/A | 1.3 | N/A | N/A | N/A | **2.04** | **2.92** | 1.28 | 1.6 | 1.51 | 2.06 | 1.06 | 1.31 | N/A | MTX + TCZ (8 mg/kg) | 0.83 | **0.7** | 0.64 | 0.65 | 0.72 | 0.97 | 0.77 | 0.93 | 0.88 | **0.57** | 0.84 | 0.38 | 1.22 | **0.47** |
|  | (1.94-3.28) |  |  | (0.82-2.64) |  |  | (0.67-2.52) |  |  |  | (1.06-3.90) | (1.76-4.83) | (0.77-2.13) | (0.95-2.69) | (0.97-2.35) | (1.26-3.36) | (0.67-1.66) | (0.82-2.09) |  |  | (0.72-0.96) | (0.57-0.85) | (0.34-1.21) | (0.39-1.09) | (0.44-1.18) | (0.60-1.57) | (0.38-1.58) | (0.58-1.51) | (0.57-1.35) | (0.40-0.82) | (0.58-1.24) | (0.08-1.73) | (0.77-1.96) | (0.30-0.76) |
|  | **2.03** | N/A | N/A | 1.19 | N/A | N/A | 1.05 | N/A | N/A | N/A | 1.64 | **2.36** | 1.03 | 1.29 | 1.22 | 1.66 | 0.85 | 1.06 | **0.81** | N/A | TCZ (8 mg/kg) | 0.84 | 0.77 | 0.78 | 0.86 | 1.17 | 0.93 | 1.12 | 0.73 | **0.69** | 1.02 | 0.45 | 1.47 | **0.57** |
|  | (1.54-2.68) |  |  | (0.66-2.14) |  |  | (0.54-2.05) |  |  |  | (0.85-3.16) | (1.42-3.93) | (0.62-1.73) | (0.76-2.18) | (0.78-1.91) | (1.01-2.73) | (0.54-1.35) | (0.66-1.70) | (0.67-0.97) |  |  | (0.68-1.04) | (0.40-1.46) | (0.46-1.31) | (0.52-1.42) | (0.72-1.89) | (0.45-1.91) | (0.69-1.82) | (0.48-1.12) | (0.48-0.99) | (0.69-1.49) | (0.10-2.08) | (0.92-2.36) | (0.36-0.91) |
|  | **1.74** | N/A | N/A | 1.02 | N/A | N/A | 0.9 | N/A | N/A | N/A | 1.41 | **2.02** | 0.89 | 1.11 | 1.05 | 1.43 | 0.73 | 0.91 | **0.69** | 0.86 | N/A | MTX + TCZ 4 mg/kg | 0.91 | 0.93 | 1.03 | 1.39 | 1.11 | 1.34 | 1.05 | 0.82 | 1.21 | 0.54 | **1.76** | 0.68 |
|  | (1.31-2.32) |  |  | (0.56-1.84) |  |  | (0.46-1.76) |  |  |  | (0.73-2.73) | (1.21-3.39) | (0.53-1.49) | (0.65-1.88) | (0.6-1.65) | (0.87-2.35) | (0.46-1.17) | (0.56-1.47) | **(0.57-0.85)** | (0.69-1.06) |  |  | (0.48-1.75) | (0.55-1.58) | (0.61-1.72) | (0.84-2.28) | (0.54-2.29) | (0.81-2.20) | (0.67-1.64) | (0.56-1.20) | (0.81-1.81) | (0.12-2.49) | (1.08-2.85) | (0.42-1.10) |
|  | **1.75** | N/A | N/A | 1.02 | N/A | N/A | 0.9 | N/A | N/A | N/A | 1.41 | **2.03** | 0.89 | 1.11 | 1.05 | 1.43 | 0.73 | 0.91 | 0.69 | 0.86 | 1 | N/A | MTX + IFX (3mg/kg) | 1.02 | 1.12 | 1.52 | 1.21 | 1.46 | 1.15 | 0.9 | 1.33 | 0.59 | 1.92 | 0.74 |
|  | (1.06-2.88) |  |  | (0.50-2.10) |  |  | (0.64-1.28) |  |  |  | (0.65-3.07) | (1.05-3.92) | (0.46-1.72) | (0.57-2.17) | (0.57-1.93) | (0.75-2.73) | (0.39-1.36) | (0.48-1.71) | (0.40-1.22) | (0.49-1.52) | (0.57-1.78) |  |  | (0.50-2.06) | (0.56-2.24) | (0.77-3.00) | (0.51-2.87) | (0.74-2.88) | (0.61-2.16) | (0.49-1.63) | (0.72-2.45) | (0.12-2.92) | (0.98-3.76) | (0.39-1.44) |
|  | N/A | N/A | N/A | N/A | N/A | N/A | N/A | N/A | N/A | N/A | N/A | N/A | N/A | N/A | N/A | N/A | N/A | N/A | N/A | N/A | N/A | N/A | N/A | GOL (100 mg) | 1.11 | **1.49** | 1.19 | 1.44 | 1.13 | 0.88 | 1.3 | 0.58 | **1.89** | 0.73 |
|  |  |  |  |  |  |  |  |  |  |  |  |  |  |  |  |  |  |  |  |  |  |  |  |  | (0.78-1.57) | (1.07-2.08) | (0.55-2.59) | (0.82-2.54) | (0.67-1.90) | (0.55-1.41) | (0.80-2.12) | (0.12-2.74) | (1.08-3.30) | (0.42-1.27) |
|  | N/A | N/A | N/A | N/A | N/A | N/A | N/A | N/A | N/A | N/A | N/A | N/A | N/A | N/A | N/A | N/A | N/A | N/A | N/A | N/A | N/A | N/A | N/A | N/A | MTX + GOL (100 mg) | **1.35** | 1.08 | 1.3 | 1.02 | 0.8 | 1.18 | 0.53 | **1.71** | 0.66 |
|  |  |  |  |  |  |  |  |  |  |  |  |  |  |  |  |  |  |  |  |  |  |  |  |  |  | (1.02-1.80) | (0.50-2.32) | (0.75-2.25) | (0.62-1.69) | (0.51-1.25) | (0.74-1.88) | (0.11-2.47) | (1.00-2.93) | (0.39-1.13) |
|  | N/A | N/A | N/A | N/A | N/A | N/A | N/A | N/A | N/A | N/A | N/A | N/A | N/A | N/A | N/A | N/A | N/A | N/A | N/A | N/A | N/A | N/A | N/A | N/A | N/A | MTX  + GOL (50 mg) | 0.8 | 0.96 | 0.75 | **0.59** | 0.87 | 0.39 | 1.26 | **0.49** |
|  |  |  |  |  |  |  |  |  |  |  |  |  |  |  |  |  |  |  |  |  |  |  |  |  |  |  | (0.38-1.70) | (0.56-1.64) | (0.46-1.23) | (0.39-0.91) | (0.56-1.36) | (0.08-1.82) | (0.75-2.14) | (0.29-0.83) |
|  | N/A | N/A | N/A | N/A | N/A | N/A | N/A | N/A | N/A | N/A | N/A | N/A | N/A | N/A | N/A | N/A | N/A | N/A | N/A | N/A | N/A | N/A | N/A | N/A | N/A | N/A | ETA  (50 mg) | 1.21 | 0.94 | 0.74 | 1.09 | 0.49 | 1.58 | 0.61 |
|  |  |  |  |  |  |  |  |  |  |  |  |  |  |  |  |  |  |  |  |  |  |  |  |  |  |  |  | (0.71-2.05) | (0.46-1.94) | (0.38-1.46) | (0.55-2.18) | (0.10-2.48) | (0.75-3.33) | (0.29-1.29) |
|  | N/A | N/A | N/A | N/A | N/A | N/A | N/A | N/A | N/A | N/A | N/A | N/A | N/A | N/A | N/A | N/A | N/A | N/A | N/A | N/A | N/A | N/A | N/A | N/A | N/A | N/A | N/A | MTX + ETA  (50 mg) | 0.78 | **0.61** | 0.91 | 0.4 | 1.31 | **0.51** |
|  |  |  |  |  |  |  |  |  |  |  |  |  |  |  |  |  |  |  |  |  |  |  |  |  |  |  |  |  | (0.48-1.27) | (0.40-0.94) | (0.58-1.42) | (0.09-1.89) | (0.78-2.22) | (0.30-0.86) |
|  | **1.93** | N/A | N/A | 1.12 | N/A | N/A | 0.99 | N/A | N/A | N/A | 1.56 | **2.23** | 0.98 | 1.22 | 1.16 | N/A | 1.58 | 0.81 | 1 | 0.76 | 0.95 | 1.1 | 1.1 | N/A | N/A | N/A | N/A | N/A | MTX + ETA  (25 mg) | 0.78 | 1.16 | 0.52 | 1.68 | **0.65** |
|  | (1.14-3.25) |  |  | (0.54-2.35) |  |  | (0.45-2.22) |  |  |  | (0.71-3.44) | (1.14-4.39) | (0.50-1.93) | (0.61-2.44) | (0.61-2.17) |  | (0.81-3.06) | (0.43-1.53) | (0.52-1.92) | (0.43-1.37) | (0.52-1.71) | (0.61-2.00) | (0.54-2.27) |  |  |  |  |  |  | (0.55-1.13) | (0.78-1.71) | (0.11-2.37) | (1.04-2.70) | (0.50-0.84) |
|  | **1.56** | N/A | N/A | 0.91 | N/A | N/A | 0.81 | N/A | N/A | N/A | 1.26 | **1.81** | 0.8 | 0.99 | 0.94 | N/A | 1.28 | 0.66 | 0.81 | **0.62** | 0.77 | 0.9 | 0.89 | N/A | N/A | N/A | N/A | N/A | 0.81 | MTX + CZP  (400 mg) | **1.47** | 0.66 | **2.14** | 0.83 |
|  | (1.36-1.80) |  |  | (0.53-1.57) |  |  | (0.43-1.51) |  |  |  | (0.69-2.33) | (1.15-2.85) | (0.50-1.25) | (0.62-1.59) | (0.64-1.37) |  | (0.83-1.97) | (0.44-0.97) | (0.54-1.23) | (0.46-0.84) | (0.56-1.05) | (0.65-1.23) | (0.53-1.50) |  |  |  |  |  | (0.47-1.39) |  | (1.08-2.01) | (0.15-2.97) | (1.42-3.23) | (0.55-1.25) |
|  | 1.01 | N/A | N/A | 0.59 | N/A | N/A | 0.52 | N/A | N/A | N/A | 0.82 | 1.17 | **0.51** | 0.64 | **0.60** | N/A | 0.82 | 0.42 | **0.52** | **0.40** | **0.50** | **0.58** | 0.58 | N/A | N/A | N/A | N/A | N/A | 0.52 | 0.64 | MTX + ADA  (40 mg) | 0.45 | 1.45 | **0.56** |
|  | (0.76-1.33) |  |  | (0.33-1.06) |  |  | (0.27-1.02) |  |  |  | (0.48-1.38) | (0.84-1.62) | (0.31-0.86) | (0.38-1.08) | (0.39-0.95) |  | (0.50-1.35) | (0.27-0.67) | (0.33-0.84) | (0.27-0.59) | (0.34-0.73) | (0.39-0.86) | (0.33-1.02) |  |  |  |  |  | (0.29-0.95) | (0.47-0.88) |  | (0.10-2.02) | (0.94-2.24) | (0.36-0.87) |
|  | 2.33 | N/A | N/A | 1.36 | N/A | N/A | 1.2 | N/A | N/A | N/A | 1.88 | 2.7 | 1.18 | 1.48 | 1.4 | N/A | 1.9 | 0.98 | 1.21 | 0.92 | 1.14 | 1.33 | 1.33 | N/A | N/A | N/A | N/A | N/A | 1.21 | 1.49 | 2.31 | MTX + ABC (125 mg) | 3.25 | 1.26 |
|  | (0.32-16.95) |  |  | (0.19-9.51) |  |  | (0.16-9.23) |  |  |  | (0.25-14.40) | (0.37-19.81) | (0.16-8.70) | (0.20-10.89) | (0.19-10.09) |  | (0.26-13.92) | (0.14-6.59) | (0.18-8.35) | (0.13-6.59) | (0.16-8.17) | (0.19-9.54) | (0.18-9.92) |  |  |  |  |  | (0.16-9.07) | (0.21-10.48) | (0.32-16.49) |  | (0.70-15.08) | (0.27-5.84) |
|  | N/A | N/A | N/A | N/A | N/A | N/A | N/A | N/A | N/A | N/A | N/A | N/A | N/A | N/A | N/A | N/A | N/A | N/A | N/A | N/A | N/A | N/A | N/A | N/A | N/A | N/A | N/A | N/A | N/A | N/A | N/A | N/A | MTX + ABC (10 mg/kg) | **0.39** |
|  |  |  |  |  |  |  |  |  |  |  |  |  |  |  |  |  |  |  |  |  |  |  |  |  |  |  |  |  |  |  |  |  |  | (0.23-0.65) |
|  | 1.21 | N/A | N/A | 0.74 | N/A | N/A | 0.65 | N/A | N/A | N/A | 1.02 | 1.47 | 0.64 | 0.8 | 0.76 | N/A | 1.04 | 0.53 | 0.66 | **0.50** | 0.62 | 0.73 | 0.72 | N/A | N/A | N/A | N/A | N/A | 0.66 | 0.81 | 1.26 | 0.54 | N/A | PBO |
|  | (0.71-2.07) |  |  | (0.34-1.62) |  |  | (0.28-1.52) |  |  |  | (0.45-2.36) | (0.71-3.03) | (0.31-1.33) | (0.39-1.68) | (0.38-1.50) |  | (0.51-2.12) | (0.27-1.06) | (0.33-1.33) | (0.27-0.95) | (0.33-1.19) | (0.38-1.39) | (0.34-1.56) |  |  |  |  |  | (0.41-1.05) | (0.44-1.48) | (0.66-2.40) | (0.07-4.15) |  |  |

##### Significant RR (CI) are in bold. ABC (125 mg) = abatacept 125 mg subcutaneously injection (SC) once weekly; ABP-501 (40 mg) = adalimumab biosimilar 40 mg; ADA (40 mg) = adalimumab 40 mg SC injection every other week; BAR (2 or 4 mg) = baricitinib 2 or 4 mg orally once daily; CT-P10 (1000 mg) = biosimilar rituximab 1000 mg + MTX; CT-P13 (3 mg/kg) = infliximab biosimilar 3 mg/kg + MTX; CZP (400 mg) = certolizumab pegol 400 mg SC every 4 weeks; csDMARDs = sulfasalazine and hydroxychloroquine; GOL (50 or 100 mg) = golimumab 50 or 100 mg SC every 4 weeks; HD203 (25 mg) = etanercept biosimilar 25 mg twice weekly SC; LBEC0101 (50 mg) = etanercept biosimilar 50 mg once weekly SC + MTX; MTX = methotrexate; PBO = placebo; PF-06438179/GP1111 (3 mg/kg) = infliximab biosimilar 3 mg/kg + MTX; RTX (500 or 1,000 mg) = rituximab 500 or 1,000 mg IV 2 courses (15 days apart) every 24 weeks; SB2 (3 mg/kg) = infliximab biosimilar 3 mg/kg + MTX; SB4 (50 mg) = etanercept biosimilar 50 mg SC once weekly + MTX; SB5 (40 mg) = SB5 adalimumab biosimilar 40 mg every other week SC + MTX; TCZ (4 or 8 mg/kg) = tocilizumab 4 or 8 mg/kg IV every 4 weeks; TOF (5 or 10 mg) = tofacitinib 5 or 10 mg orally twice daily.

##### *Supplementary figure 1 SUCRA graph for remission at 24-26 weeks*

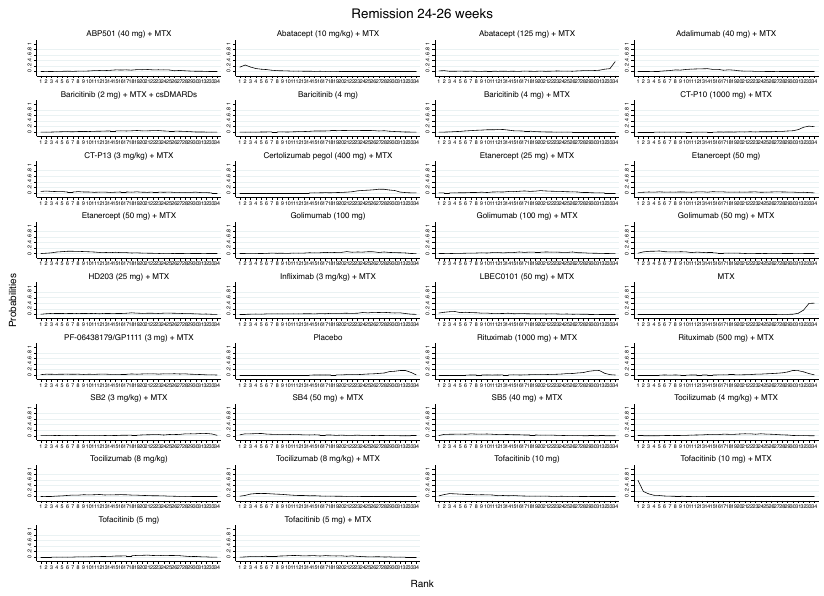

Abatacept (125 mg) = abatacept 125 mg subcutaneously injection (SC) once weekly; Abatacept (10 mg/kg) = abatacept 10 mg/kg intravenous infusion (IV) every 4 weeks; ABP-501 (40 mg) = adalimumab biosimilar 40 mg; Adalimumab (20 or 40 mg) = adalimumab 20 or 40 mg SC injection every other week; Baricitinib (2 or 4 mg) = baricitinib 2 or 4 mg orally once daily; Certolizumab pegol (400 mg) = certolizumab pegol 400 mg SC every 4 weeks; csDMARDs = sulfasalazine and hydroxychloroquine; CT-P10 (1000 mg) = biosimilar rituximab 1000 mg + MTX; CT-P13 (3 mg/kg) = infliximab biosimilar 3 mg/kg + MTX; Etanercept (25 or 50 mg) = etanercept 25 or 50 mg SC once weekly; Golimumab (50 or 100 mg) = golimumab 50 or 100 mg SC every 4 weeks; HD203 (25 mg) = etanercept biosimilar 25 mg twice weekly SC;

Infliximab (3 mg/kg) = infliximab 3 mg/kg IV every 8 weeks; LBEC0101 (50 mg) = etanercept biosimilar 50 mg once weekly SC + MTX; MTX = methotrexate; PBO = placebo; PF-06438179/GP1111 (3 mg/kg) = infliximab biosimilar 3 mg/kg + MTX; Rituximab (500 or 1,000 mg) = rituximab 500 or 1,000 mg IV 2 courses (15 days apart) every 24 weeks; SB2 (3 mg/kg) = infliximab biosimilar 3 mg/kg + MTX; SB4 (50 mg) = etanercept biosimilar 50 mg SC once weekly + MTX; SB5 (40 mg) = SB5 adalimumab biosimilar 40 mg every other week SC + MTX; Tocilizumab (4 or 8 mg/kg) = tocilizumab 4 or 8 mg/kg IV every 4 weeks; Tofacitinib (5 or 10 mg) = tofacitinib 5 or 10 mg orally twice daily.

##### *Supplementary figure 2 SUCRA graph for remission at 48-52 weeks*

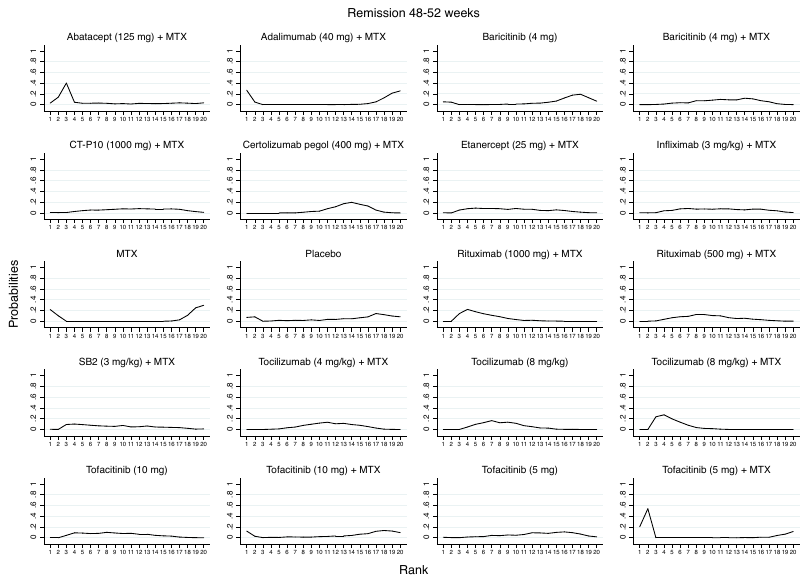

Abatacept (125 mg) = abatacept 125 mg subcutaneously injection (SC) once weekly; Adalimumab (40 mg) = adalimumab 40 mg SC injection every other week; Baricitinib (4 mg) = baricitinib 4 mg orally once daily; Certolizumab pegol (400 mg) = certolizumab pegol 400 mg SC every 4 weeks; csDMARDs = sulfasalazine and hydroxychloroquine; CT-P10 (1000 mg) = biosimilar rituximab 1000 mg + MTX; Etanercept (25 mg) = etanercept 25 mg SC once weekly; Infliximab (3 mg/kg) = infliximab 3 mg/kg IV every 8 weeks; MTX = methotrexate; PBO = placebo; Rituximab (500 or 1,000 mg) = rituximab 500 or 1,000 mg IV 2 courses (15 days apart) every 24 weeks; SB2 (3 mg/kg) = infliximab biosimilar 3 mg/kg + MTX; Tocilizumab (4 or 8 mg/kg) = tocilizumab 4 or 8 mg/kg IV every 4 weeks; Tofacitinib (5 or 10 mg) = tofacitinib 5 or 10 mg orally twice daily.

**Safety outcomes**

##### *Supplementary table 11 Serious infection at 24-26 weeks*

| Ranking | Medication (dose) | Number of study (patients) | RR (95% CI) |
| --- | --- | --- | --- |
| 1 | Rituximab (500 mg) + MTX | 2 (291) | 0.26 (0.04-1.55) |
| 2 | Certolizumab pegol (400 mg) + MTX | 1 (316) | 4.72 (0.26-85.34) |
| 3 | Filgotinib (200 mg) + MTX + csDMARDs | 1 (148) | 0.50 (0.04-5.64) |
| 4 | Etanercept biosimilar SB5 (50 mg) + MTX | 1 (271) | 0.69 (0.05-9.40) |
| 5 | Baricitinib (2 mg) + MTX +/- csDMARDs | 2 (403) | 0.67 (0.24-1.82) |
| 6 | MTX | 11 (2,495) | Comparator |
| 7 | Baricitinib (4 mg) | 1 (159) | 0.79 (0.15-4.09) |
| 8 | Filgotinib (100 mg) + MTX + csDMARDs | 1 (153) | 1.45 (0.24-8.94) |
| 9 | Rituximab (1,000 mg) + MTX | 2 (362) | 0.90 (0.28-2.86) |
| 10 | Golimumab (100 mg) + MTX | 2 (248) | 2.33 (0.62-8.81) |
| 11 | Golimumab (100 mg) | 2 (292) | 2.37 (0.67-8.41) |
| 12 | Golimumab (50 mg) + MTX | 2 (248) | 1.10 (0.26-4.63) |
| 13 | Baricitinib (4 mg) + MTX +/- csDMARDs | 4 (1,106) | 1.08 (0.56-2.10) |
| 14 | Adalimumab (40 mg) + MTX | 11 (1,118) | 1.37 (0.53-3.58) |

Adalimumab (40 mg) = adalimumab 40 mg SC injection every other week; Baricitinib (2 or 4 mg) = baricitinib 2 or 4 mg orally once daily;

Certolizumab pegol (400 mg) = certolizumab pegol 400 mg SC every 4 weeks; csDMARDs = sulfasalazine and hydroxychloroquine; Filgotinib (200 mg) = filgotinib 200 mg orally once daily; Golimumab (50 or 100 mg) = golimumab 50 or 100 mg SC every 4 weeks;

MTX = methotrexate; Rituximab (500 or 1,000 mg) = rituximab 500 or 1,000 mg IV 2 courses (15 days apart) every 24 weeks; SB5 (40 mg) = adalimumab biosimilar SB5 40 mg every other week SC.

##### *Supplementary table 12 Serious infection at 48-52 weeks*

| Ranking | Medication (dose) | Number of study (patients) | RR (95% CI) |
| --- | --- | --- | --- |
| 1 | Rituximab (500 mg) + MTX | 1 (249) | 0.46 (0.16-1.37) |
| 2 | Rituximab biosimilar CT-P10 + MTX | 1 (161) | 0.40 (0.05-3.06) |
| 3 | Rituximab (1000 mg) + MTX | 3 (501) | 0.61 (0.22-1.68) |
| 4 | Tofacitinib (10 mg) + MTX | 1 (316) | 6.74 (0.36-124.86) |
| 5 | Certolizumab pegol (400 mg) + MTX | 3 (973) | 0.80 (0.41-1.54) |
| 6 | MTX | 12 (2,426) | Comparator |
| 7 | Abatacept (125 mg) + MTX | 1 (41) | 1.20 (0.09-16.42) |
| 8 | Adalimumab (20 mg) + MTX | 1 (212) | 1.03 (0.18-5.78) |
| 9 | Baricitinib (4 mg) | 1 (159) | 1.41 (0.44-4.52) |
| 10 | Baricitinib (4 mg) + MTX +/- csDMARDs | 2 (702) | 1.47 (0.53-4.08) |
| 11 | Tocilizumab (8 mg/kg) | 2 (407) | 1.46 (0.48-4.39) |
| 12 | Tocilizumab (8 mg/kg) + MTX | 2 (409) | 1.52 (0.52-4.49) |
| 13 | Tocilizumab (4 mg/kg) + MTX | 1 (290) | 1.83 (0.60-5.55) |
| 14 | Tofacitinib (5 mg) | 1 (386) | 1.95 (0.37-10.28) |
| 15 | Adalimumab (40 mg) + MTX | 6 (1,373) | 1.94 (0.64-5.89) |
| 16 | Tofacitinib (5 mg) + MTX | 2 (699) | 3.32 (0.68-16.17) |

Abatacept (125 mg) = abatacept 125 mg subcutaneously injection (SC) once weekly; Adalimumab (40 mg) = adalimumab 40 mg SC injection every other week; Baricitinib (4 mg) = baricitinib 4 mg orally once daily; Certolizumab pegol (400 mg) = certolizumab pegol 400 mg SC every 4 weeks; CT-P10 (1,000 mg) = rituximab biosimilar CT-P10 1,000 mg; MTX = methotrexate; csDMARDs = sulfasalazine and hydroxychloroquine; Rituximab (500 or 1,000 mg) = rituximab 500 or 1,000 mg IV 2 courses (15 days apart) every 24 weeks; Tocilizumab (4 or 8 mg/kg) = tocilizumab 4 or 8 mg/kg IV every 4 weeks; Tofacitinib (5 or 10 mg) = tofacitinib 5 or 10 mg orally twice daily.

##### *Supplementary table 13: Tuberculosis at 24-26 weeks*

| Ranking | Medication (dose) | Number of study (patients) | RR (95% CI) |
| --- | --- | --- | --- |
| 1 | MTX | 4 (939) | Comparator |
| 2 | Golimumab (100 mg) + MTX | 1 (159) | 1.01 (0.02-50.40) |
| 3 | Golimumab (100 mg) | 1 (159) | 1.01 (0.02-50.40) |
| 4 | Filgotinib (100 mg) + MTX + csDMARDs | 1 (153) | 0.97 (0.02-48.45) |
| 5 | Filgotinib (200 mg) + MTX + csDMARDs | 1 (148) | 1.00 (0.02-50.07) |
| 6 | Adalimumab (40 mg) + MTX | 1 (515) | 3.01 (0.12-73.76) |
| 7 | Golimumab (50 mg) + MTX | 1 (159) | 3.02 (0.12-73.55) |
| 8 | Certolizumab pegol (400 mg) + MTX | 1 (316) | 3.26 (0.18-60.17) |

Adalimumab (20 or 40 mg) = adalimumab 20 or 40 mg SC injection every other week; Certolizumab pegol (400 mg) = certolizumab pegol 400 mg SC every 4 weeks; csDMARDs = sulfasalazine and hydroxychloroquine; Golimumab (50 or 100 mg) = golimumab 50 or 100 mg SC every 4 weeks; Filgotinib (100 or 200 mg) = filgotinib 100 or 200 mg orally once daily; MTX = methotrexate.

##### *Supplementary table 14: Tuberculosis at 48-52 weeks*

| Ranking | Medication (dose) | Number of study (patients) | RR (95% CI) |
| --- | --- | --- | --- |
| 1 | Adalimumab (40 mg) | 1 (274) | 0.65 (0.02-17.00) |
| 2 | Baricitinib (4 mg) + MTX + csDMARDs | 2 (702) | 0.65 (0.05-7.99) |
| 3 | Adalimumab (20 mg) + MTX | 1 (212) | 0.65 (0.02-16.99) |
| 4 | Tofacitinib (5 mg) | 1 (386) | 0.77 (0.02-31.63) |
| 5 | MTX | 10 (2,049) | Comparator |
| 6 | Certolizumab pegol (400 mg) + MTX | 3 (973) | 0.98 (0.12-7.97) |
| 7 | Baricitinib (4 mg) | 1 (159) | 1.07 (0.03-39.92) |
| 8 | Adalimumab biosimilar SB5 (40 mg) + MTX | 1 (271) | 1.78 (0.03-113.06) |
| 9 | Adalimumab (40 mg) + MTX | 8 (1,932) | 1.76 (0.44-7.02) |
| 10 | Tofacitinib (5 mg) + MTX | 2 (582) | 3.35 (0.27-40.92) |
| 11 | Tofacitinib (10 mg) + MTX | 1 (201) | 7.43 (0.74-74.35) |

Adalimumab (20 or 40 mg) = adalimumab 20 or 40 mg SC injection every other week; Baricitinib (4 mg) = baricitinib 4 mg orally once daily; Certolizumab pegol (400 mg) = certolizumab pegol 400 mg SC every 4 weeks; csDMARDs = sulfasalazine and hydroxychloroquine; MTX = methotrexate; SB5 = adalimumab biosimilar SB5 40 mg every other week SC; Tofacitinib (5 or 10 mg) = tofacitinib 5 or 10 mg orally twice daily.

##### *Supplementary table 15: Herpes zoster at 24-26 weeks*

| Ranking | Medication (dose) | Number of study (patients) | RR (95% CI) |
| --- | --- | --- | --- |
| 1 | MTX | 5 (1,250) | Comparator |
| 2 | Baricitinib (2 mg) + MTX +/- csDMARDs | 2 (403) | 2.47 (0.69-8.84) |
| 3 | Adalimumab (40 mg) + MTX | 1 (330) | 2.97 (0.75-11.81) |
| 4 | Baricitinib (4 mg) + MTX +/- csDMARDs | 4 (1,106) | **3.52** (1.38-9.02) |
| 5 | Filgotinib (100 mg) + MTX + csDMARDs | 1 (153) | 4.84 (0.23-99.93) |
| 6 | Filgotinib (200 mg) + MTX + csDMARDs | 1 (148) | 5.00 (0.24-103.27) |
| 7 | Baricitinib (4 mg) | 1 (159) | 4.55 (0.88-23.57) |

Significant RR (CI) are in bold. Adalimumab (40 mg) = adalimumab 40 mg SC injection every other week; Baricitinib (2 or 4 mg) = baricitinib 2 or 4 mg orally once daily; csDMARDs = sulfasalazine and hydroxychloroquine; Filgotinib (100 or 200 mg) = filgotinib 100 or 200 mg orally once daily; MTX = methotrexate.

##### *Supplementary table 16: Herpes zoster at 48-52 weeks*

| Ranking | Medication (dose) | Number of study (patients) | RR (95% CI) |
| --- | --- | --- | --- |
| 1 | MTX | 4 (994) | Comparator |
| 2 | Tofacitinib (5 mg) | 1 (386) | 2.64 (0.44-15.81) |
| 3 | Adalimumab (20 mg) + MTX | 1 (212) | 1.00 (0.04-26.04) |
| 4 | Adalimumab (40 mg) + MTX | 5 (1,203) | 3.15 (0.82-12.13) |
| 5 | Baricitinib (4 mg) | 1 (159) | 3.90 (0.87-17.54) |
| 6 | Baricitinib (4 mg) + MTX +/- csDMARDs | 2 (702) | **4.20** (1.22-14.48) |
| 7 | Tofacitinib (5 mg) + MTX | 1 (378) | **5.38** (1.00-28.91) |

Significant RR (CI) are in bold. Adalimumab (40 mg) = adalimumab 40 mg SC injection every other week; Baricitinib (4 mg) = baricitinib 4 mg orally once daily; csDMARDs = sulfasalazine and hydroxychloroquine; MTX = methotrexate; Tofacitinib (5 mg) = tofacitinib 5 mg orally twice daily.

##### *Supplementary table 17: Cancer at 24-26 weeks*

| Ranking | Medication (dose) | Number of study (patients) | RR (95% CI) |
| --- | --- | --- | --- |
| 1 | Filgotinib (100 mg) + MTX + csDMARDs | 1 (153) | 0.97 (0.02-54.11) |
| 2 | Baricitinib (4 mg) | 1 (159) | 0.63 (0.02-17.98) |
| 3 | Filgotinib (200 mg) + MTX + csDMARDs | 1 (148) | 1.00 (0.02-55.92) |
| 4 | Certolizumab pegol (400 mg) + MTX | 1 (316) | 1.09 (0.04-30.35) |
| 5 | Adalimumab biosimilar ABP50 (40 mg) + MTX | 1 (264) | 1.17 (0.03-45.61) |
| 6 | Baricitinib (2 mg) + MTX +/- csDMARDs | 2 (403) | 0.58 (0.05-7.26) |
| 7 | MTX | 12 (2,511) | Comparator |
| 8 | Golimumab (100 mg) | 2 (292) | 0.64 (0.09-4.45) |
| 9 | Rituximab (500 mg) + MTX | 2 (291) | 3.57 (0.44-28.89) |
| 10 | Golimumab (50 mg) + MTX | 2 (248) | 0.71 (0.10-4.97) |
| 11 | Golimumab (100 mg) + MTX | 2 (248) | 1.08 (0.13-9.05) |
| 12 | Adalimumab (40 mg) + MTX | 4 (1,380) | 1.18 (0.13-10.72) |
| 13 | Rituximab (1,000 mg) + MTX | 2 (362) | 1.79 (0.20-15.75) |
| 14 | Baricitinib (4 mg) + MTX +/- csDMARDs | 4 (1,106) | 1.95 (0.46-8.26) |

ABP501 (40 mg) = adalimumab biosimilar ABP50 40 mg every 2 weeks; Adalimumab (40 mg) = adalimumab 40 mg SC injection every other week; Baricitinib (2 or 4 mg) = baricitinib 2 or 4 mg orally once daily; Certolizumab pegol (400 mg) = certolizumab pegol 400 mg SC every 4 weeks; csDMARDs = sulfasalazine and hydroxychloroquine; Filgotinib (100 or 200 mg) = filgotinib 100 or 200 mg orally once daily; Golimumab (50 or 100 mg) = golimumab 50 or 100 mg SC every 4 weeks; MTX = methotrexate; Rituximab (1,000 mg) = rituximab 1,000 mg IV 2 courses (15 days apart) every 24 weeks.

##### *Supplementary table 18: Cancer at 48-52 weeks*

| Ranking | Medication (dose) | Number of study (patients) | RR (95% CI) |
| --- | --- | --- | --- |
| 1 | Abatacept (125 mg) + MTX | 1 (41) | 0.06 (0.00-2.97) |
| 2 | Tofacitinib (5 mg) | 1 (386) | 6.63 (0.62-70.53) |
| 3 | Rituximab (1,000 mg) + MTX | 2 (290) | 0.20 (0.02-1.69) |
| 4 | Abatacept (10 mg/kg) + MTX | 1 (203) | 4.98 (0.24-102.99) |
| 5 | Adalimumab (20 mg) + MTX | 1 (212) | 0.27 (0.01-6.94) |
| 6 | Certolizumab pegol (400 mg) + MTX | 1 (159) | 2.96 (0.12-72.17) |
| 7 | MTX | 9 (2,066) | Comparator |
| 8 | Rituximab (500 mg) + MTX | 1 (249) | 0.40 (0.08-2.04) |
| 9 | Tocilizumab (8 mg/kg) | 2 (407) | 0.49 (0.09-2.73) |
| 10 | Tocilizumab (8 mg/kg) + MTX | 2 (409) | 0.59 (0.08-4.51) |
| 11 | Baricitinib (4 mg) + MTX +/- csDMARDs | 2 (702) | 3.63 (0.60-21.99) |
| 12 | Baricitinib (4 mg) | 1 (159) | 2.55 (0.20-32.84) |
| 13 | Tofacitinib (5 mg) + MTX | 3 (903) | 2.22 (0.29-16.77) |
| 14 | Tofacitinib (10 mg) + MTX | 2 (517) | 1.74 (0.21-14.66) |
| 15 | Tocilizumab (4 mg/kg) + MTX | 1 (290) | 1.33 (0.30-5.88) |
| 16 | Adalimumab (40 mg) + MTX | 5 (1,299) | 2.14 (0.42-11.08) |

Abatacept (125 mg) = abatacept 125 mg subcutaneously injection (SC) once weekly; Abatacept (10 mg/kg) = abatacept 10 mg/kg intravenous infusion (IV) every 4 weeks; Adalimumab (20 or 40 mg) = adalimumab 20 or 40 mg SC injection every other week; Baricitinib (4 mg) = baricitinib 4 mg orally once daily; Certolizumab pegol (400 mg) = certolizumab pegol 400 mg SC every 4 weeks; csDMARDs = sulfasalazine and hydroxychloroquine; MTX = methotrexate; Rituximab (500 or 1,000 mg) = rituximab 500 or 1,000 mg IV 2 courses (15 days apart) every 24 weeks; Tocilizumab (4 or 8 mg/kg) = tocilizumab 4 or 8 mg/kg IV every 4 weeks; Tofacitinib (5 or 10 mg) = tofacitinib 5 or 10 mg orally twice daily.

##### *Supplementary table 19: Cardiovascular events at 24-26 weeks*

| Ranking | Medication (dose) | Number of study (patients) | RR (95% CI) |
| --- | --- | --- | --- |
| 1 | Baricitinib (4 mg) | 1 (159) | 4.61 (0.33-64.28) |
| 2 | Certolizumab pegol (400 mg) + MTX | 1 (316) | 0.12 (0.00-2.95) |
| 3 | Baricitinib (2 mg) + MTX +/- csDMARDs | 2 (403) | 0.30 (0.03-2.85) |
| 4 | Filgotinib (100 mg) + MTX + csDMARDs | 1 (153) | 0.33 (0.01-8.12) |
| 5 | Adalimumab biosimilar ABP501 (40 mg) + MTX | 1 (264) | 0.45 (0.02-11.06) |
| 6 | Filgotinib (200 mg) + MTX + csDMARDs | 1 (148) | 0.97 (0.10-9.20) |
| 7 | MTX | 8 (2,053) | Comparator |
| 8 | Rituximab (1,000 mg) + MTX | 1 (170) | 1.77 (0.53-5.94) |
| 9 | Adalimumab (40 mg) + MTX | 3 (1,107) | 0.91 (0.11-7.60) |
| 10 | Baricitinib (4 mg) + MTX +/- csDMARDs | 4 (1,106) | 1.33 (0.29-6.01) |
| 11 | Rituximab (500 mg) + MTX | 1 (167) | 1.29 (0.35-4.71) |

ABP501 (40 mg) = adalimumab biosimilar ABP501 40 mg every 2 weeks; Adalimumab (40 mg) = adalimumab 40 mg SC injection every other week; Baricitinib (2 or 4 mg) = baricitinib 2 or 4 mg orally once daily; Certolizumab pegol (400 mg) = certolizumab pegol 400 mg SC every 4 weeks; csDMARDs = sulfasalazine and hydroxychloroquine; Filgotinib (100 or 200 mg) = filgotinib 100 or 200 mg orally once daily; MTX = methotrexate; Rituximab (500 or 1,000 mg) = rituximab 500 or 1,000 mg IV 2 courses (15 days apart) every 24 weeks.

##### *Supplementary table 20: Cardiovascular events at 48-52 weeks*

| Ranking | Medication (dose) | Number of study (patients) | RR (95% CI) |
| --- | --- | --- | --- |
| 1 | Rituximab (1000 mg) + MTX | 1 (250) | 1.44 (0.84-2.48) |
| 2 | Tocilizumab (4 mg/kg) + MTX | 1 (290) | 2.49 (0.49-12.74) |
| 3 | MTX | 5 (1,344) | Comparator |
| 4 | Adalimumab (40 mg) + MTX | 2 (534) | 1.65 (0.31-8.89) |
| 5 | Baricitinib (4 mg) | 1 (159) | 1.07 (0.15-7.70) |
| 6 | Baricitinib (4 mg) + MTX +/- csDMARDs | 2 (702) | 1.21 (0.19-7.52) |
| 7 | Rituximab (500 mg) + MTX | 1 (249) | 1.10 (0.62-1.96) |
| 8 | Tofacitinib (5 mg) + MTX | 2 (525) | 0.69 (0.07-6.62) |
| 9 | Tocilizumab (8 mg/kg) | 1 (292) | 0.49 (0.05-5.43) |
| 10 | Tocilizumab (8 mg/kg) + MTX | 1 (291) | 0.50 (0.05-5.45) |
| 11 | Tofacitinib (10 mg) + MTX | 2 (517) | 0.70 (0.09-5.38) |

Adalimumab (40 mg) = adalimumab 40 mg SC injection every other week; Baricitinib (4 mg) = baricitinib 4 mg orally once daily; csDMARDs = sulfasalazine and hydroxychloroquine; MTX = methotrexate; Rituximab (500 or 1,000 mg) = rituximab 500 or 1,000 mg IV 2 courses (15 days apart) every 24 weeks; Tocilizumab (4 or 8 mg/kg) = tocilizumab 4 or 8 mg/kg IV every 4 weeks; Tofacitinib (5 or 10 mg) = tofacitinib 5 or 10 mg orally twice daily.

**Comparative effect of treatments**

**Efficacy**

*Supplementary table 20 Serious infection*

| **RR (CI) of serious infection at 24-26 weeks** | **RR (CI) of serious infection at 48-52 weeks** | | | | | | | | | | | | | | | | | | | | | | |
| --- | --- | --- | --- | --- | --- | --- | --- | --- | --- | --- | --- | --- | --- | --- | --- | --- | --- | --- | --- | --- | --- | --- | --- |
|  | MTX | 0.4 | 6.74 | 3.32 | 1.95 | 1.47 | 1.41 | 0.61 | 0.46 | 1.52 | 1.46 | 1.83 | 0.8 | 1.94 | 1.03 | 1.2 | N/A | N/A | N/A | N/A | N/A | N/A | N/A |
|  |  | (0.05-3.06) | (0.36-124.86) | (0.68-16.17) | (0.37-10.28) | (0.53-4.08) | (0.44-4.52) | (0.22-1.68) | (0.16-01.37) | (0.52-4.49) | (0.48-4.39) | (0.60-5.55) | (0.41-1.54) | (0.64-5.89) | (0.18-5.78) | (0.09-16.42) |  |  |  |  |  |  |  |
|  | N/A | MTX +  CT-P10 | 16.79 | 8.26 | 4.86 | 3.67 | 3.52 | 1.53 | 1.15 | 3.8 | 3.62 | 4.55 | 1.98 | 4.83 | 2.57 | 2.98 | N/A | N/A | N/A | N/A | N/A | N/A | N/A |
|  |  |  | (0.48-588.09) | (0.63-108.63) | (0.35-67.05) | (0.38-35.60) | (0.34-36.57) | (0.26-8.90) | (0.14-9.52) | (0.38-37.91) | (0.36-36.60) | (0.45-46.09) | (0.23-16.78) | (0.48-48.92) | (0.18-36.83) | (0.15-59.36) |  |  |  |  |  |  |  |
|  | N/A | N/A | MTX + TOF (10 mg) | 0.49 | 0.29 | 0.22 | 0.21 | 0.09 | 0.07 | 0.23 | 0.22 | 0.27 | 0.12 | 0.29 | 0.15 | 0.18 | N/A | N/A | N/A | N/A | N/A | N/A | N/A |
|  |  |  |  | (0.04-5.71) | (0.02-4.30) | (0.01-3.85) | (0.01-4.29) | (0.00-1.99) | (0.00-1.54) | (0.01-5.13) | (0.01-4.88) | (0.01-6.15) | (0.01-2.39) | (0.02-4.28) | (0.01-2.92) | (0.00-8.95) |  |  |  |  |  |  |  |
|  | N/A | N/A | N/A | MTX + TOF (5 mg) | 0.59 | 0.44 | 0.43 | 0.18 | **0.14** | 0.46 | 0.44 | 0.55 | 0.24 | 0.58 | 0.31 | 0.36 | N/A | N/A | N/A | N/A | N/A | N/A | N/A |
|  |  |  |  |  | (0.19-1.82) | (0.10-1.97) | (0.07-2.48) | (0.03-1.21) | (0.02-0.95) | (0.07-3.17) | (0.06-3.01) | (0.08-3.81) | (0.04-1.37) | (0.19-1.81) | (0.06-1.61) | (0.02-7.69) |  |  |  |  |  |  |  |
|  | N/A | N/A | N/A | N/A | TOF (5 mg) | 0.76 | 0.72 | 0.31 | 0.24 | 0.78 | 0.75 | 0.94 | 0.41 | 0.99 | 0.53 | 0.61 | N/A | N/A | N/A | N/A | N/A | N/A | N/A |
|  |  |  |  |  |  | (0.16-3.65) | (0.12-4.54) | (0.04-2.20) | (0.03-1.72) | (0.11-5.76) | (0.10-5.48) | (0.13-6.92) | (0.07-2.51) | (0.29-3.43) | (0.09-2.95) | (0.03-13.65) |  |  |  |  |  |  |  |
|  | 1.08 | N/A | N/A | N/A | N/A | BAR 4 mg  + MTX  (+/- csDMARDs) | 0.96 | 0.42 | 0.31 | 1.03 | 0.99 | 1.24 | 0.54 | 1.32 | 0.7 | 0.81 | N/A | N/A | N/A | N/A | N/A | N/A | N/A |
|  | (0.56-2.10) |  |  |  |  |  | (0.29-3.16) | (0.10-1.74) | (0.07-1.38) | (0.23-4.62) | (0.22-4.42) | (0.27-5.59) | (0.16-1.86) | (0.50-3.47) | (0.14-3.37) | (0.05-13.47) |  |  |  |  |  |  |  |
|  | 0.79 | N/A | N/A | N/A | N/A | 0.73 | BAR  (4 mg) | 0.43 | 0.33 | 1.08 | 1.03 | 1.29 | 0.56 | 1.37 | 0.73 | 0.85 | N/A | N/A | N/A | N/A | N/A | N/A | N/A |
|  | (0.15-4.09) |  |  |  |  | (0.14-3.71) |  | (0.09-2.02) | (0.07-1.60) | (0.22-5.33) | (0.21-5.12) | (0.26-6.46) | (0.15-2.18) | (0.35-5.32) | (0.12-4.59) | (0.05-14.87) |  |  |  |  |  |  |  |
|  | 0.9 | N/A | N/A | N/A | N/A | 0.83 | 1.13 | MTX + RTX  (1000 mg) | 0.75 | 2.49 | 2.37 | 2.98 | 1.3 | 3.17 | 1.68 | 1.95 | N/A | N/A | N/A | N/A | N/A | N/A | N/A |
|  | (0.28-2.86) |  |  |  |  | (0.22-3.16) | (0.15-8.46) |  | (0.23-2.42) | (0.57-10.91) | (0.53-10.60) | (0.66-13.37) | (0.39-4.34) | (0.71-14.18) | (0.23-12.38) | (0.17-21.89) |  |  |  |  |  |  |  |
|  | 0.26 | N/A | N/A | N/A | N/A | 0.24 | 0.32 | 0.29 | MTX + RTX (500 mg) | 3.3 | 3.15 | 3.96 | 1.72 | 4.2 | 2.23 | 2.59 | N/A | N/A | N/A | N/A | N/A | N/A | N/A |
|  | (0.04-1.55) |  |  |  |  | (0.04-1.61) | (0.03-3.69) | (0.05-1.80) |  | (0.71-15.28) | (0.67-14.83) | (0.84-18.71) | (0.48-6.14) | (0.89-19.85) | (0.29-17.11) | (0.18-37.96) |  |  |  |  |  |  |  |
|  | N/A | N/A | N/A | N/A | N/A | N/A | N/A | N/A | N/A | MTX + TCZ  (8 mg/kg) | 0.95 | 1.2 | 0.52 | 1.27 | 0.68 | 0.78 | N/A | N/A | N/A | N/A | N/A | N/A | N/A |
|  |  |  |  |  |  |  |  |  |  |  | (0.43-2.10) | (0.47-3.06) | (0.15-1.85) | (0.27-6.11) | (0.09-5.28) | (0.05-13.34) |  |  |  |  |  |  |  |
|  | N/A | N/A | N/A | N/A | N/A | N/A | N/A | N/A | N/A | N/A | TCZ  (8 mg/kg) | 1.26 | 0.55 | 1.33 | 0.71 | 0.82 | N/A | N/A | N/A | N/A | N/A | N/A | N/A |
|  |  |  |  |  |  |  |  |  |  |  |  | (0.48-3.29) | (0.15-1.98) | (0.28-6.36) | (0.09-5.46) | (0.05-14.11) |  |  |  |  |  |  |  |
|  | N/A | N/A | N/A | N/A | N/A | N/A | N/A | N/A | N/A | N/A | N/A | MTX +  TCZ  (4 mg/kg) | 0.44 | 1.06 | 0.56 | 0.65 | N/A | N/A | N/A | N/A | N/A | N/A | N/A |
|  |  |  |  |  |  |  |  |  |  |  |  |  | (0.12-1.59) | (0.22-5.11) | (0.07-4.38) | (0.04-11.26) |  |  |  |  |  |  |  |
|  | 4.72 | N/A | N/A | N/A | N/A | 4.36 | 5.95 | 5.27 | 18.32 | N/A | N/A | N/A | MTX +  CZP  (400 mg) | 2.44 | 1.29 | 1.5 | N/A | N/A | N/A | N/A | N/A | N/A | N/A |
|  | (0.26-85.34) |  |  |  |  | (0.22-84.97) | (0.21-166.05) | (0.23-119.30) | (0.61-552.22) |  |  |  |  | (0.65-9.20) | (0.20-8.51) | (0.10-22.40) |  |  |  |  |  |  |  |
|  | 1.37 | N/A | N/A | N/A | N/A | 1.27 | 1.73 | 1.53 | 5.33 | N/A | N/A | N/A | 0.29 | MTX + ADA (40 mg) | 0.53 | 0.62 | N/A | N/A | N/A | N/A | N/A | N/A | N/A |
|  | (0.53-3.58) |  |  |  |  | (0.41-3.96) | (0.26-11.53) | (0.34-6.84) | (0.69-40.94) |  |  |  | (0.01-6.15) |  | (0.16-1.75) | (0.04-10.60) |  |  |  |  |  |  |  |
|  | N/A | N/A | N/A | N/A | N/A | N/A | N/A | N/A | N/A | N/A | N/A | N/A | N/A | N/A | ADA (40 mg) | 1.16 | N/A | N/A | N/A | N/A | N/A | N/A | N/A |
|  |  |  |  |  |  |  |  |  |  |  |  |  |  |  |  | (0.05-26.70) |  |  |  |  |  |  |  |
|  | N/A | N/A | N/A | N/A | N/A | N/A | N/A | N/A | N/A | N/A | N/A | N/A | N/A | N/A | N/A | MTX + ABC (125 mg) | N/A | N/A | N/A | N/A | N/A | N/A | N/A |
|  | 0.5 | N/A | N/A | N/A | N/A | 2.16 | 1.59 | 1.79 | 0.51 | N/A | N/A | N/A | 9.43 | 2.74 | N/A | N/A | FIL (200 mg) + MTX +csDMARDs | N/A | N/A | N/A | N/A | N/A | N/A |
|  | (0.04-5.64) |  |  |  |  | (0.18-26.66) | (0.08-29.58) | (0.12-26.30) | (0.03-10.48) |  |  |  | (0.22-411.30) | (0.20-37.14) |  |  |  |  |  |  |  |  |  |
|  | 1.45 | N/A | N/A | N/A | N/A | 0.75 | 0.55 | 0.62 | 0.18 | N/A | N/A | N/A | 3.25 | 0.95 | N/A | N/A | 2.9 | FIL (100 mg)  + MTX +csDMARDs | N/A | N/A | N/A | N/A | N/A |
|  | (0.24-8.94) |  |  |  |  | (0.11-5.16) | (0.05-6.33) | (0.07-5.34) | (0.01-2.28) |  |  |  | (0.11-99.29) | (0.12-7.39) |  |  | (0.29-28.55) |  |  |  |  |  |  |
|  | 0.69 | N/A | N/A | N/A | N/A | 1.57 | 1.15 | 1.3 | 0.37 | N/A | N/A | N/A | 6.82 | 1.99 | N/A | N/A | 1.38 | 0.48 | MTX + SB5 (40 mg) | N/A | N/A | N/A | N/A |
|  | (0.05-9.40) |  |  |  |  | (0.11-22.87) | (0.05-24.95) | (0.07-22.43) | (0.02-8.87) |  |  |  | (0.14-336.58) | (0.18-22.49) |  |  | (0.04-48.64) | (0.02-11.46) |  |  |  |  |  |
|  | 0.67 | N/A | N/A | N/A | N/A | 0.62 | 1.19 | 1.34 | 0.39 | N/A | N/A | N/A | 7.08 | 2.06 | N/A | N/A | 1.33 | 0.46 | 0.96 | BAR (2 mg) + MTX (+/- csDMARDs) | N/A | N/A | N/A |
|  | (0.24-1.82) |  |  |  |  | (0.23-1.67) | (0.19-7.63) | (0.29-6.26) | (0.05-3.02) |  |  |  | (0.33-151.93) | (0.53-8.08) |  |  | (0.10-18.35) | (0.06-3.67) | (0.06-15.61) |  |  |  |  |
|  | 2.37 | N/A | N/A | N/A | N/A | 2.19 | 2.99 | 2.65 | **9.22** | N/A | N/A | N/A | 1.99 | 0.58 | N/A | N/A | 4.74 | 1.63 | 3.43 | 3.56 | GOL (100 mg) (+/- PBO) | N/A | N/A |
|  | (0.67-8.41) |  |  |  |  | (0.52-9.25) | (0.37-23.94) | (0.48-14.67) | (1.02-82.95) |  |  |  | (0.08-46.86) | (0.13-2.64) |  |  | (0.31-72.94) | (0.18-14.98) | (0.20-60.05) | (0.71-17.92) |  |  |  |
|  | 2.33 | N/A | N/A | N/A | N/A | 2.15 | 2.94 | 2.6 | 9.05 | N/A | N/A | N/A | 2.02 | 0.59 | N/A | N/A | 4.66 | 1.61 | 3.37 | 3.5 | 0.98 | MTX + GOL (100 mg) | N/A |
|  | (0.62-8.81) |  |  |  |  | (0.49-9.53) | (0.35-24.35) | (0.45-15.20) | (0.97-84.42) |  |  |  | (0.08-49.00) | (0.12-3.00) |  |  | (0.29-73.86) | (0.17-15.28) | (0.18-62.63) | (0.66-18.55) | (0.29-3.29) |  |  |
|  | 1.1 | N/A | N/A | N/A | N/A | 1.02 | 1.39 | 1.23 | 4.28 | N/A | N/A | N/A | 4.28 | 1.24 | N/A | N/A | 2.2 | 0.76 | 1.6 | 1.66 | 0.46 | 0.47 | MTX + GOL (50 mg) |
|  | (0.26-4.63) |  |  |  |  | (0.21-4.91) | (0.16-12.27) | (0.19-7.83) | (0.43-42.50) |  |  |  | (0.17-108.36) | (0.21-7.26) |  |  | (0.13-36.82) | (0.07-7.70) | (0.08-32.04) | (0.29-9.57) | (0.12-1.79) | (0.13-1.70) |  |

Significant RR (CI) are in bold. ABC (125 mg) = abatacept 125 mg subcutaneously injection (SC) once weekly; ADA (40 mg) = adalimumab 40 mg SC injection every other week; BAR (2 or 4 mg) = baricitinib 2 or 4 mg orally once daily; CZP (400 mg) = certolizumab pegol 400 mg SC every 4 weeks; csDMARDs = sulfasalazine and hydroxychloroquine; CT-P10 (1,000 mg) = rituximab biosimilar CT-P10 1,000 mg; FIL (100 or 200 mg) = filgotinib 100 or 200 mg orally once daily; GOL (50 or 100 mg) = golimumab 50 or 100 mg SC every 4 weeks; MTX = methotrexate; SB5 (40 mg) = adalimumab biosimilar SB5 40 mg every other week SC; TCZ (4 or 8 mg/kg) = tocilizumab 4 or 8 mg/kg IV every 4 weeks, RTX (500 or 1,000 mg) = rituximab 500 or 1,000 mg IV 2 courses (15 days apart) every 24 weeks; TOF (5 or 10 mg) = tofacitinib 5 or 10 mg orally twice daily.

##### *Supplementary table 21 Tuberculosis*

| **RR (CI) of Tuberculosis at 24-26 weeks** | **RR (CI) of Tuberculosis at 48-52 weeks** | | | | | | | | | | | | | | | |
| --- | --- | --- | --- | --- | --- | --- | --- | --- | --- | --- | --- | --- | --- | --- | --- | --- |
|  | MTX | 0.65 | 1.78 | 7.43 | 3.35 | 0.77 | 0.65 | 1.07 | 0.98 | 1.76 | 0.65 | N/A | N/A | N/A | N/A | N/A |
|  |  | (0.02,16.99) | (0.03-113.06) | (0.74-74.35) | (0.27-40.92) | (0.02-31.63) | (0.05-7.99) | (0.03-39.92) | (0.12-7.97) | (0.44-7.02) | (0.02,17.00) |  |  |  |  |  |
|  | N/A | MTX + ADA 20 mg | 2.73 | 11.43 | 5.16 | 1.19 | 1.00 | 1.65 | 1.51 | 2.71 | 1.00 | N/A | N/A | N/A | N/A | N/A |
|  |  |  | (0.02,407.08) | (0.26,498.82) | (0.11,237.97) | (0.01,130.83) | (0.02,52.01) | (0.01,201.19) | (0.03,73.03) | (0.12,61.07) | (0.01,79.56) |  |  |  |  |  |
|  | N/A | N/A | MTX + SB5 (40 mg) | 4.18 | 1.89 | 0.44 | 0.36 | 0.6 | 0.55 | 0.99 | 0.37 | N/A | N/A | N/A | N/A | N/A |
|  |  |  |  | (0.05-377.95) | (0.02-175.27) | (0.00-85.72) | (0.00-38.84) | (0.00-137.24) | (0.01-57.88) | (0.02-49.85) | (0.00,54.47) |  |  |  |  |  |
|  | N/A | N/A | N/A | MTX + TOF (10 mg) | 0.45 | 0.1 | 0.09 | 0.14 | 0.13 | 0.24 | 0.09 | N/A | N/A | N/A | N/A | N/A |
|  |  |  |  |  | (0.04-5.67) | (0.00-4.63) | (0.00-2.25) | (0.00-9.89) | (0.01-2.97) | (0.03-2.20) | (0.00,3.82) |  |  |  |  |  |
|  | N/A | N/A | N/A | N/A | MTX + TOF (5 mg) | 0.23 | 0.19 | 0.32 | 0.29 | 0.53 | 0.19 | N/A | N/A | N/A | N/A | N/A |
|  |  |  |  |  |  | (0.01-4.71) | (0.01-5.50) | (0.00-24.06) | (0.01-7.66) | (0.05-5.15) | (0.00,8.96) |  |  |  |  |  |
|  | N/A | N/A | N/A | N/A | N/A | TOF (5 mg) | 0.84 | 1.39 | 1.27 | 2.28 | 0.84 | N/A | N/A | N/A | N/A | N/A |
|  |  |  |  |  |  |  | (0.01-62.81) | (0.01-230.05) | (0.02-89.94) | (0.07-78.96) | (0.01,92.31) |  |  |  |  |  |
|  | N/A | N/A | N/A | N/A | N/A | N/A | BAR (4 mg) + MTX (+/- csDMARDs) | 1.66 | 1.52 | 2.72 | 1.00 | N/A | N/A | N/A | N/A | N/A |
|  |  |  |  |  |  |  |  | (0.04-61.55) | (0.06-39.85) | (0.21-34.50) | (0.02,52.29) |  |  |  |  |  |
|  | N/A | N/A | N/A | N/A | N/A | N/A | N/A | BAR (4 mg) | 0.92 | 1.64 | 0.6 | N/A | N/A | N/A | N/A | N/A |
|  |  |  |  |  |  |  |  |  | (0.01-59.65) | (0.04-70.12) | (0.00,73.67) |  |  |  |  |  |
|  | 3.26 | N/A | N/A | N/A | N/A | N/A | N/A | N/A | MTX + CZP (400 mg) | 1.79 | 0.66 | N/A | N/A | N/A | N/A | N/A |
|  | (0.18-60.17) |  |  |  |  |  |  |  |  | (0.15-22.01) | (0.01,31.91) |  |  |  |  |  |
|  | 3.01 | N/A | N/A | N/A | N/A | N/A | N/A | N/A | 0.92 | MTX + ADA (40 mg) | 0.37 | N/A | N/A | N/A | N/A | N/A |
|  | (0.12-73.76) |  |  |  |  |  |  |  | (0.01-69.82) |  | (0.02,8.29) |  |  |  |  |  |
|  | N/A | N/A | N/A | N/A | N/A | N/A | N/A | N/A | N/A | N/A | ADA (40 mg) | N/A | N/A | N/A | N/A | N/A |
|  | 1 | N/A | N/A | N/A | N/A | N/A | N/A | N/A | 3.26 | 3.01 | N/A | FIL (200 mg) + MTX + csDMARDs | N/A | N/A | N/A | N/A |
|  | (0.02-50.07) |  |  |  |  |  |  |  | (0.02-429.37) | (0.02-471.78) |  |  |  |  |  |  |
|  | 0.97 | N/A | N/A | N/A | N/A | N/A | N/A | N/A | 3.37 | 3.11 | N/A | 0.97 | FIL (100 mg) + MTX +csDMARDs | N/A | N/A | N/A |
|  | (0.02-48.45) |  |  |  |  |  |  |  | (0.03-443.82) | (0.02-487.65) |  | (0.02-48.45) |  |  |  |  |
|  | 1.01 | N/A | N/A | N/A | N/A | N/A | N/A | N/A | 3.24 | 2.99 | N/A | 1.01 | 1.04 | GOL  (100 mg)  (+/- PBO) | N/A | N/A |
|  | (0.02-50.40) |  |  |  |  |  |  |  | (0.02-426.87) | (0.02-469.02) |  | (0.00-254.90) | (0.00-263.47) |  |  |  |
|  | 1.01 | N/A | N/A | N/A | N/A | N/A | N/A | N/A | 3.24 | 2.99 | N/A | 1.01 | 1.04 | 1 | MTX + GOL (100 mg) | N/A |
|  | (0.02-50.40) |  |  |  |  |  |  |  | (0.02-426.87) | (0.02-469.02) |  | (0.00-254.90) | (0.00-263.47) | (0.02-50.09) |  |  |
|  | 3.02 | N/A | N/A | N/A | N/A | N/A | N/A | N/A | 1.08 | 1 | N/A | 3.02 | 3.12 | 3 | 3 | MTX +  GOL (50 mg) |
|  | (0.12-73.55) |  |  |  |  |  |  |  | (0.01-81.55) | (0.01-91.56) |  | (0.02-471.36) | (0.02-487.21) | (0.12-73.09) | (0.12-73.09) |  |

ADA (20 or 40 mg) = adalimumab 20 or 40 mg SC injection every other week; BAR (4 mg) = baricitinib 4 mg orally once daily; CZP (400 mg) = certolizumab pegol 400 mg SC every 4 weeks; csDMARDs = sulfasalazine and hydroxychloroquine; FIL (100 or 200 mg) = filgotinib 100 or 200 mg orally once daily; GOL (100 mg) = golimumab 100 mg SC every 4 weeks; MTX = methotrexate; SB5 (40 mg) = adalimumab biosimilar SB5 40 mg every other week SC; TOF (5 or 10 mg) = tofacitinib 5 or 10 mg orally twice daily.

*Supplementary table 22 Herpes zoster*

| **RR (CI) of Herpes zoster at 24-26 weeks** | **RR (CI) of Herpes zoster at 48-52 weeks** | | | | | | | | | |
| --- | --- | --- | --- | --- | --- | --- | --- | --- | --- | --- |
|  | MTX | **5.38** | 2.64 | **4.20** | 3.9 | 3.15 | 1.00 | N/A | N/A | N/A |
|  |  | (1.00-28.91) | (0.44-15.81) | (1.22-14.48) | (0.87-17.54) | (0.82-12.13) | (0.04-26.04) |  |  |  |
|  | N/A | MTX + TOF (5 mg) | 0.49 | 0.78 | 0.72 | 0.58 | 0.19 | N/A | N/A | N/A |
|  |  |  | (0.17-1.42) | (0.20-3.10) | (0.12-4.48) | (0.21-1.59) | (0.01-4.90) |  |  |  |
|  | N/A | N/A | TOF (5 mg) | 1.59 | 1.48 | 1.19 | 0.38 | N/A | N/A | N/A |
|  |  |  |  | (0.35-7.22) | (0.22-10.15) | (0.37-3.88) | (0.01-10.62) |  |  |  |
|  | **3.52** | N/A | N/A | BAR (4 mg) + MTX (+/- csDMARDs) | 0.93 | 0.75 | 0.24 | N/A | N/A | N/A |
|  | (1.38-9.02) |  |  |  | (0.26-3.28) | (0.29-1.93) | (0.01-5.91) |  |  |  |
|  | 4.55 | N/A | N/A | 1.29 | BAR (4 mg) | 0.81 | 0.26 | N/A | N/A | N/A |
|  | (0.88-23.57) |  |  | (0.29-5.81) |  | (0.18-3.71) | (0.01-7.69) |  |  |  |
|  | 2.97 | N/A | N/A | 0.84 | 0.65 | MTX + ADA  (40 mg) | 0.32 | N/A | N/A | N/A |
|  | (0.75-11.81) |  |  | (0.26-2.77) | (0.10-4.32) |  | (0.01-7.17) |  |  |  |
|  | N/A | N/A | N/A | N/A | N/A | N/A | ADA (40 mg) | N/A | N/A | N/A |
|  | 5.00 | N/A | N/A | 0.70 | 0.91 | 0.59 | N/A | FIL (200 mg) + MTX +csDMARDs | N/A | N/A |
|  | (0.24-103.26) |  |  | (0.03-16.78) | (0.03-28.55) | (0.02-16.55) |  |  |  |  |
|  | 4.84 | N/A | N/A | 0.73 | 0.94 | 0.61 | N/A | 0.97 | FIL (100 mg) + MTX +csDMARDs | N/A |
|  | (0.23-99.93) |  |  | (0.03-17.35) | (0.03-29.51) | (0.02-17.11) |  | (0.17-5.50) |  |  |
|  | 2.47 | N/A | N/A | 0.70 | 1.84 | 1.20 | N/A | 0.49 | 0.51 | BAR (2 mg) + MTX (+/- csDMARDs) |
|  | (0.69-8.84) |  |  | (0.25-1.94) | (0.31-11.07) | (0.26-5.62) |  | (0.02-13.20) | (0.02-13.65) |  |

Significant RR (CI) are in bold. ADA (40 mg) = adalimumab 40 mg SC injection every other week; BAR (4 mg) = baricitinib 4 mg orally once daily; csDMARDs = sulfasalazine and hydroxychloroquine; FIL (100 or 200 mg) = filgotinib 100 or 200 mg orally once daily; MTX = methotrexate; TCZ (4 or 8 mg/kg) = tocilizumab 4 or 8 mg/kg IV every 4 weeks; RTX (500 or 1,000 mg) = rituximab 500 or 1,000 mg IV 2 courses (15 days apart) every 24 weeks; TOF (5 or 10 mg) = tofacitinib 5 or 10 mg orally twice daily.

##### *Supplementary table 23 Cancer*

| **RR (CI) of cancer 24-26 weeks** |  |  |  |  |  |  |  |  |  |  |  |  |  |  |  |  |  |  |  |  |  |  |  |
| --- | --- | --- | --- | --- | --- | --- | --- | --- | --- | --- | --- | --- | --- | --- | --- | --- | --- | --- | --- | --- | --- | --- | --- |
|  | **RR (CI) of cancer at 48-52 weeks** | | | | | | | | | | | | | | | | | | | | | | |
|  | MTX | 1.74 | 2.22 | 6.63 | 3.63 | 2.55 | 0.2 | 0.4 | 0.59 | 0.49 | 1.33 | 2.96 | 2.14 | 0.27 | 0.06 | 4.98 | N/A | N/A | N/A | N/A | N/A | N/A | N/A |
|  |  | (0.21-14.66) | (0.29-16.77) | (0.62-70.53) | (0.60-21.99) | (0.20-32.84) | (0.02-1.69) | (0.08-2.04) | (0.08-4.51) | (0.09-2.73) | (0.30-5.88) | (0.12-72.17) | (0.42-11.08) | (0.01-6.94) | (0.00-2.97) | (0.24-102.99) |  |  |  |  |  |  |  |
|  | N/A | MTX + TOF 10 mg | 1.28 | 3.82 | 2.09 | 1.47 | 0.11 | 0.23 | 0.34 | 0.28 | 0.76 | 1.71 | 1.23 | 0.15 | 0.04 | 2.86 | N/A | N/A | N/A | N/A | N/A | N/A | N/A |
|  |  |  | (0.43-3.79) | (0.40-36.07) | (0.16-26.65) | (0.06-37.05) | (0.01-2.35) | (0.02-3.37) | (0.02-6.49) | (0.02-4.37) | (0.06-10.30) | (0.04-79.33) | (0.22-6.85) | (0.01-4.41) | (0.00-2.98) | (0.07-116.48) |  |  |  |  |  |  |  |
|  | N/A | N/A | MTX + TOF 5 mg | 2.99 | 1.64 | 1.15 | 0.09 | 0.18 | 0.27 | 0.22 | 0.6 | 1.34 | 0.97 | 0.12 | 0.03 | 2.24 | N/A | N/A | N/A | N/A | N/A | N/A | N/A |
|  |  |  |  | (0.37-24.20) | (0.14-19.01) | (0.05-26.99) | (0.00-1.71) | (0.01-2.42) | (0.02-4.70) | (0.02-3.15) | (0.05-7.38) | (0.03-58.53) | (0.20-4.60) | (0.00-3.21) | (0.00-2.22) | (0.06-85.75) |  |  |  |  |  |  |  |
|  | N/A | N/A | N/A | TOF 5 mg | 0.55 | 0.38 | **0.03** | 0.06 | 0.09 | 0.07 | 0.2 | 0.45 | 0.32 | 0.04 | 0.01 | 0.75 | N/A | N/A | N/A | N/A | N/A | N/A | N/A |
|  |  |  |  |  | (0.04-8.12) | (0.01-11.16) | (0.00-0.73) | (0.00-1.07) | (0.00-2.02) | (0.00-1.38) | (0.01-3.28) | (0.01-23.76) | (0.05-1.91) | (0.00-1.21) | (0.00-0.88) | (0.02-35.05) |  |  |  |  |  |  |  |
|  | 1.95 | N/A | N/A | N/A | BAR 4 mg + MTX (+/- csDMARDs) | 0.7 | **0.05** | 0.11 | 0.16 | 0.14 | 0.37 | 0.82 | 0.59 | 0.07 | 0.02 | 1.37 | N/A | N/A | N/A | N/A | N/A | N/A | N/A |
|  | (0.46-8.26) |  |  |  |  | (0.05-9.05) | (0.00-0.90) | (0.01-1.25) | (0.01-2.47) | (0.01-1.63) | (0.04-3.79) | (0.02-31.92) | (0.07-4.70) | (0.00-2.48) | (0.00-1.23) | (0.04-46.57) |  |  |  |  |  |  |  |
|  | 0.63 | N/A | N/A | N/A | 0.32 | BAR 4 mg | 0.08 | 0.16 | 0.23 | 0.19 | 0.52 | 1.16 | 0.84 | 0.1 | 0.03 | 1.95 | N/A | N/A | N/A | N/A | N/A | N/A | N/A |
|  | (0.02-17.98) |  |  |  | (0.01-7.32) |  | (0.00-2.20) | (0.01-3.26) | (0.01-6.10) | (0.01-4.20) | (0.03-10.06) | (0.02-69.59) | (0.05-15.33) | (0.00-6.03) | (0.00-2.54) | (0.04-103.05) |  |  |  |  |  |  |  |
|  | 1.79 | N/A | N/A | N/A | 0.92 | 2.85 | MTX + RTX 1000 mg | 2.01 | 2.98 | 2.48 | 6.67 | 14.87 | 10.77 | 1.34 | 0.33 | 24.98 | N/A | N/A | N/A | N/A | N/A | N/A | N/A |
|  | (0.20-15.75) |  |  |  | (0.06-13.26) | (0.05-161.42) |  | (0.18-22.00) | (0.16-56.84) | (0.16-38.34) | (0.49-90.38) | (0.32-694.48) | (0.73-159.76) | (0.03-66.05) | (0.01-7.76) | (0.61-1019.87) |  |  |  |  |  |  |  |
|  | 3.57 | N/A | N/A | N/A | 1.83 | 5.67 | 1.99 | MTX + RTX 500 mg | 1.48 | 1.23 | 3.32 | 7.41 | 5.36 | 0.67 | 0.16 | 12.44 | N/A | N/A | N/A | N/A | N/A | N/A | N/A |
|  | (0.44-28.89) |  |  |  | (0.14-24.02) | (0.11-301.65) | (0.29-13.62) |  | (0.11-20.03) | (0.12-13.11) | (0.37-30.20) | (0.21-267.07) | (0.53-54.24) | (0.02-25.50) | (0.00-8.62) | (0.40-388.28) |  |  |  |  |  |  |  |
|  | N/A | N/A | N/A | N/A | N/A | N/A | N/A | N/A | MTX + TCZ 8 mg/kg | 0.83 | 2.24 | 4.99 | 3.61 | 0.45 | 0.11 | 8.38 | N/A | N/A | N/A | N/A | N/A | N/A | N/A |
|  |  |  |  |  |  |  |  |  |  | (0.13-5.43) | (0.32-15.68) | (0.11-219.07) | (0.27-49.06) | (0.01-20.85) | (0.00-8.29) | (0.22-320.99) |  |  |  |  |  |  |  |
|  | N/A | N/A | N/A | N/A | N/A | N/A | N/A | N/A | N/A | TCZ 8 mg/kg | 2.69 | 6.01 | 4.35 | 0.54 | 0.13 | 10.09 | N/A | N/A | N/A | N/A | N/A | N/A | N/A |
|  |  |  |  |  |  |  |  |  |  |  | (0.54-13.56) | (0.16-224.99) | (0.41-46.63) | (0.01-21.47) | (0.00-8.69) | (0.31-327.61) |  |  |  |  |  |  |  |
|  | N/A | N/A | N/A | N/A | N/A | N/A | N/A | N/A | N/A | N/A | MTX + TCZ 4 mg/kg | 2.23 | 1.61 | 0.2 | 0.05 | 3.74 | N/A | N/A | N/A | N/A | N/A | N/A | N/A |
|  |  |  |  |  |  |  |  |  |  |  |  | (0.07-75.53) | (0.18-14.80) | (0.01-7.22) | (0.00-2.96) | (0.13-109.52) |  |  |  |  |  |  |  |
|  | 1.09 | N/A | N/A | N/A | 0.56 | 1.73 | 0.61 | 0.66 | N/A | N/A | N/A | MTX + CZP (400 mg) | 0.72 | 0.09 | 0.02 | 1.68 | N/A | N/A | N/A | N/A | N/A | N/A | N/A |
|  | (0.04-30.35) |  |  |  | (0.01-20.95) | (0.02-194.14) | (0.01-32.27) | (0.03-14.74) |  |  |  |  | (0.02-26.25) | (0.00-8.63) | (0.00-3.19) | (0.02-137.07) |  |  |  |  |  |  |  |
|  | 1.18 | N/A | N/A | N/A | 0.61 | 1.88 | 0.3 | 0.33 | N/A | N/A | N/A | 1.09 | MTX + ADA 40 mg | 0.12 | 0.03 | 2.32 | N/A | N/A | N/A | N/A | N/A | N/A | N/A |
|  | (0.13-10.72) |  |  |  | (0.05-6.99) | (0.04-93.42) | (0.01-15.51) | (0.02-6.96) |  |  |  | (0.02-58.89) |  | (0.01-2.28) | (0.00-1.94) | (0.07-72.82) |  |  |  |  |  |  |  |
|  | N/A | N/A | N/A | N/A | N/A | N/A | N/A | N/A | N/A | N/A | N/A | N/A | N/A | ADA 40 mg | 0.24 | 18.63 | N/A | N/A | N/A | N/A | N/A | N/A | N/A |
|  |  |  |  |  |  |  |  |  |  |  |  |  |  |  | (0.00-36.93) | (0.22-1593.67) |  |  |  |  |  |  |  |
|  | N/A | N/A | N/A | N/A | N/A | N/A | N/A | N/A | N/A | N/A | N/A | N/A | N/A | N/A | MTX + ABC 125 mg | 76.75 | N/A | N/A | N/A | N/A | N/A | N/A | N/A |
|  |  |  |  |  |  |  |  |  |  |  |  |  |  |  |  | (0.58-10107.89) |  |  |  |  |  |  |  |
|  | N/A | N/A | N/A | N/A | N/A | N/A | N/A | N/A | N/A | N/A | N/A | N/A | N/A | N/A | N/A | MTX + ABC 10 mg/kg | N/A | N/A | N/A | N/A | N/A | N/A | N/A |
|  | 1 | N/A | N/A | N/A | 1.95 | 0.63 | 1.79 | 3.57 | N/A | N/A | N/A | 1.09 | 1.18 | N/A | N/A | N/A | FIL 200 mg + MTX +csDMARDs | N/A | N/A | N/A | N/A | N/A | N/A |
|  | (0.02-55.92) |  |  |  | (0.03-140.38) | (0.00-118.52) | (0.02-173.71) | (0.04-332.84) |  |  |  | (0.01-201.64) | (0.01-116.34) |  |  |  |  |  |  |  |  |  |  |
|  | 0.97 | N/A | N/A | N/A | 2.02 | 0.65 | 1.85 | 3.69 | N/A | N/A | N/A | 1.12 | 1.22 | N/A | N/A | N/A | 0.97 | FIL 100 mg + MTX +csDMARDs | N/A | N/A | N/A | N/A | N/A |
|  | (0.02-54.11) |  |  |  | (0.03-145.10) | (0.00-122.51) | (0.02-179.55) | (0.04-344.04) |  |  |  | (0.01-208.42) | (0.01-120.26) |  |  |  | (0.02-54.11) |  |  |  |  |  |  |
|  | 1.17 | N/A | N/A | N/A | 1.66 | 0.54 | 1.53 | 3.04 | N/A | N/A | N/A | 0.93 | 1.01 | N/A | N/A | N/A | 1.17 | 1.21 | MTX + ABP501 (40 mg) | N/A | N/A | N/A | N/A |
|  | (0.03-45.61) |  |  |  | (0.04-75.04) | (0.00-70.52) | (0.02-108.56) | (0.04-206.64) |  |  |  | (0.01-130.27) | (0.05-18.70) |  |  |  | (0.01-270.37) | (0.01-279.46) |  |  |  |  |  |
|  | 0.58 | N/A | N/A | N/A | 0.3 | 1.08 | 3.07 | 6.12 | N/A | N/A | N/A | 1.86 | 2.03 | N/A | N/A | N/A | 0.58 | 0.6 | 0.5 | BAR 2 mg + MTX (+/- csDMARDs) | N/A | N/A | N/A |
|  | (0.05-7.26) |  |  |  | (0.03-2.85) | (0.02-49.70) | (0.11-89.76) | (0.23-166.06) |  |  |  | (0.03-121.11) | (0.08-51.26) |  |  |  | (0.01-67.38) | (0.01-69.65) | (0.01-38.73) |  |  |  |  |
|  | 0.64 | N/A | N/A | N/A | 0.33 | 1.01 | 0.36 | 0.18 | N/A | N/A | N/A | 1.71 | 1.86 | N/A | N/A | N/A | 0.64 | 0.66 | 0.54 | 1.09 | GOL 100 mg (+/- PBO) | N/A | N/A |
|  | (0.09-4.45) |  |  |  | (0.03-3.55) | (0.02-47.54) | (0.02-6.63) | (0.01-3.12) |  |  |  | (0.04-80.63) | (0.10-34.90) |  |  |  | (0.01-55.60) | (0.01-57.47) | (0.01-34.03) | (0.05-25.66) |  |  |  |
|  | 1.08 | N/A | N/A | N/A | 0.55 | 1.71 | 0.6 | 0.3 | N/A | N/A | N/A | 1.01 | 1.1 | N/A | N/A | N/A | 1.08 | 1.11 | 0.92 | 1.85 | 1.69 | MTX + GOL 100 mg | N/A |
|  | (0.13-9.05) |  |  |  | (0.04-7.32) | (0.03-91.52) | (0.03-12.50) | (0.02-5.94) |  |  |  | (0.02-52.38) | (0.05-23.53) |  |  |  | (0.01-102.23) | (0.01-105.67) | (0.01-63.37) | (0.07-50.53) | (0.18-15.70) |  |  |
|  | 0.71 | N/A | N/A | N/A | 0.36 | 1.12 | 0.4 | 0.2 | N/A | N/A | N/A | 1.54 | 1.67 | N/A | N/A | N/A | 0.71 | 0.73 | 0.6 | 1.21 | 1.11 | 0.66 | MTX + GOL 50 mg |
|  | (0.10-4.97) |  |  |  | (0.03-3.91) | (0.02-52.53) | (0.02-7.44) | (0.01-3.49) |  |  |  | (0.03-72.60) | (0.09-31.36) |  |  |  | (0.01-61.98) | (0.01-64.07) | (0.01-37.87) | (0.05-28.34) | (0.14-8.97) | (0.07-6.12) |  |

Significant RR (CI) are in bold. ABC (125 mg) = abatacept 125 mg subcutaneously injection (SC) once weekly; ABP501 (40 mg) = adalimumab biosimilar ABP50 40 mg every 2 weeks; ADA (40 mg) = adalimumab 40 mg SC injection every other week; BAR (2 or 4 mg) = baricitinib 2 or 4 mg orally once daily; csDMARDs = sulfasalazine and hydroxychloroquine; CT-P10 (1,000 mg) = rituximab biosimilar CT-P10 1,000 mg; CZP (400 mg) = certolizumab pegol 400 mg SC every 4 weeks; FIL (100 or 200 mg) = filgotinib 100 or 200 mg orally once daily; GOL (50 or 100 mg) = golimumab 50 or 100 mg SC every 4 weeks; MTX = methotrexate; RTX (500 or 1,000 mg) = rituximab 500 or 1,000 mg IV 2 courses (15 days apart) every 24 weeks; TCZ (4 or 8 mg/kg) = tocilizumab 4 or 8 mg/kg IV every 4 weeks; TOF (5 or 10 mg) = tofacitinib 5 or 10 mg orally twice daily.

##### *Supplementary table 24 Cardiovascular events*

| **RR (CI) of cardiovascular events 24-26 weeks** | **RR (CI) of cardiovascular events at 48-52 weeks** | | | | | | | | | | | | | | | | |
| --- | --- | --- | --- | --- | --- | --- | --- | --- | --- | --- | --- | --- | --- | --- | --- | --- | --- |
|  | MTX | 0.70 | 0.69 | 1.21 | 1.07 | 1.44 | 1.1 | 0.5 | 0.49 | 2.49 | 1.65 | N/A | N/A | N/A | N/A | N/A | N/A |
|  |  | (0.09-5.38) | (0.07-6.62) | (0.19-7.52) | (0.15-7.70) | (0.84-2.48) | (0.62-1.96) | (0.05-5.45) | (0.05-5.43) | (0.49-12.74) | (0.31-8.89) |  |  |  |  |  |  |
|  | N/A | MTX + TOF  (10 mg) | 0.98 | 1.72 | 1.53 | 2.06 | 1.57 | 0.71 | 0.71 | 3.56 | 2.36 | N/A | N/A | N/A | N/A | N/A | N/A |
|  |  |  | (0.18-5.35) | (0.18-16.25) | (0.10-24.27) | (0.25-17.03) | (0.19-13.10) | (0.03-16.49) | (0.03-16.44) | (0.26-48.52) | (0.49-11.30) |  |  |  |  |  |  |
|  | N/A | N/A | MTX + TOF (5 mg) | 1.75 | 1.55 | 2.10 | 1.60 | 0.72 | 0.72 | 3.62 | 2.4 | N/A | N/A | N/A | N/A | N/A | N/A |
|  |  |  |  | (0.15-20.27) | (0.08-29.20) | (0.20-21.50) | (0.15-16.52) | (0.03-19.46) | (0.03-19.39) | (0.22-58.92) | (0.38-15.22) |  |  |  |  |  |  |
|  | 1.33 | N/A | N/A | BAR (4 mg) + MTX (+/- csDMARDs) | 0.89 | 1.20 | 0.91 | 0.41 | 0.41 | 2.06 | 1.37 | N/A | N/A | N/A | N/A | N/A | N/A |
|  | (0.29-6.01) |  |  |  | (0.07-10.55) | (0.18-8.07) | (0.13-6.21) | (0.02-8.38) | (0.02-8.35) | (0.18-23.96) | (0.24-7.93) |  |  |  |  |  |  |
|  | 4.61 | N/A | N/A | 3.47 | BAR (4 mg) | 1.35 | 1.03 | 0.46 | 0.46 | 2.33 | 1.54 | N/A | N/A | N/A | N/A | N/A | N/A |
|  | (0.33-64.28) |  |  | (0.25-48.39) |  | (0.17-10.46) | (0.13-8.05) | (0.02-10.34) | (0.02-10.31) | (0.18-30.17) | (0.13-18.70) |  |  |  |  |  |  |
|  | 1.77 | N/A | N/A | 1.33 | 0.38 | MTX + RTX  (1000 mg) | 0.76 | 0.34 | 0.34 | 1.73 | 1.14 | N/A | N/A | N/A | N/A | N/A | N/A |
|  | (0.53-5.94) |  |  | (0.19-9.23) | (0.02-6.97) |  | (0.45-1.29) | (0.03-4.01) | (0.03-3.99) | (0.31-9.63) | (0.20-6.70) |  |  |  |  |  |  |
|  | 1.29 | N/A | N/A | 0.97 | 0.28 | 0.73 | MTX + RTX  (500 mg) | 0.45 | 0.45 | 2.26 | 1.50 | N/A | N/A | N/A | N/A | N/A | N/A |
|  | (0.35-4.71) |  |  | (0.13-7.10) | (0.01-5.26) | (0.24-2.25) |  | (0.04-5.31) | (0.04-5.29) | (0.40-12.80) | (0.25-8.90) |  |  |  |  |  |  |
|  | N/A | N/A | N/A | N/A | N/A | N/A | N/A | MTX + TCZ  (8 mg/kg) | 1.00 | 5.02 | 3.33 | N/A | N/A | N/A | N/A | N/A | N/A |
|  |  |  |  |  |  |  |  |  | (0.06-15.86) | (0.59-42.68) | (0.18-62.12) |  |  |  |  |  |  |
|  | N/A | N/A | N/A | N/A | N/A | N/A | N/A | N/A | TCZ  (8 mg/kg) | 5.03 | 3.34 | N/A | N/A | N/A | N/A | N/A | N/A |
|  |  |  |  |  |  |  |  |  |  | (0.59-42.83) | (0.18-62.34) |  |  |  |  |  |  |
|  | N/A | N/A | N/A | N/A | N/A | N/A | N/A | N/A | N/A | MTX + TCZ (4 mg/kg) | 0.66 | N/A | N/A | N/A | N/A | N/A | N/A |
|  |  |  |  |  |  |  |  |  |  |  | (0.06-6.91) |  |  |  |  |  |  |
|  | N/A | N/A | N/A | N/A | N/A | N/A | N/A | N/A | N/A | N/A | MTX + ADA (40 mg) | N/A | N/A | N/A | N/A | N/A | N/A |
|  | 0.33 | N/A | N/A | 3.98 | 13.84 | 5.31 | 3.86 | N/A | N/A | N/A | N/A | FIL (200 mg) + MTX +csDMARDs | N/A | N/A | N/A | N/A | N/A |
|  | (0.01-8.12) |  |  | (0.12-136.19) | (0.22-868.33) | (0.17-161.43) | (0.12-121.19) |  |  |  |  |  |  |  |  |  |  |
|  | 0.97 | N/A | N/A | 1.37 | 4.77 | 1.83 | 1.33 | N/A | N/A | N/A | N/A | 2.9 | FIL (100 mg) + MTX +csDMARDs | N/A | N/A | N/A | N/A |
|  | (0.10-9.20) |  |  | (0.09-20.66) | (0.15-152.56) | (0.14-23.59) | (0.10-17.90) |  |  |  |  | (0.12-70.69) |  |  |  |  |  |
|  | 0.45 | N/A | N/A | 2.95 | 10.24 | 3.93 | 2.86 | N/A | N/A | N/A | N/A | 1.35 | 0.47 | MTX + ABP501 (40 mg) | N/A | N/A | N/A |
|  | (0.02-11.06) |  |  | (0.11-81.37) | (0.18-589.56) | (0.13-120.42) | (0.09-90.40) |  |  |  |  | (0.01-124.22) | (0.01-23.32) |  |  |  |  |
|  | 0.30 | N/A | N/A | 0.23 | 15.25 | 5.85 | 4.26 | N/A | N/A | N/A | N/A | 0.91 | 0.31 | 0.67 | BAR (2 mg) + MTX (+/- csDMARDs) | N/A | N/A |
|  | (0.03-2.85) |  |  | (0.02-2.15) | (0.57-410.11) | (0.46-74.95) | (0.32-56.87) |  |  |  |  | (0.02-44.93) | (0.01-7.51) | (0.01-30.34) |  |  |  |
|  | 0.12 | N/A | N/A | 0.09 | 0.03 | 0.07 | 0.09 | N/A | N/A | N/A | N/A | 0.36 | 0.12 | 0.27 | 0.4 | MTX + CZP  (400 mg) | N/A |
|  | (0.00-2.95) |  |  | (0.00-3.11) | (0.00-1.65) | (0.00-2.08) | (0.00-2.95) |  |  |  |  | (0.00-33.17) | (0.00-6.22) | (0.00-24.70) | (0.01-19.82) |  |  |
|  | 0.91 | N/A | N/A | 0.68 | 0.20 | 0.51 | 0.71 | N/A | N/A | N/A | N/A | 2.72 | 0.94 | 2.02 | 3.00 | 7.51 | MTX + ADA  (40 mg) |
|  | (0.11-7.60) |  |  | (0.07-6.79) | (0.01-5.18) | (0.04-5.91) | (0.06-8.50) |  |  |  |  | (0.06-126.05) | (0.04-20.74) | (0.18-22.09) | (0.15-58.19) | (0.16-347.73) |  |

ABC (125 mg) = abatacept 125 mg subcutaneously injection (SC) once weekly; ABP501 (40 mg) = adalimumab biosimilar ABP50 40 mg every 2 weeks; ADA (40 mg) = adalimumab 40 mg SC injection every other week; BAR (2 or 4 mg) = baricitinib 2 or 4 mg orally once daily; csDMARDs = sulfasalazine and hydroxychloroquine; CT-P10 (1,000 mg) = rituximab biosimilar CT-P10 1,000 mg; CZP (400 mg) = certolizumab pegol 400 mg SC every 4 weeks; FIL (100 or 200 mg) = filgotinib 100 or 200 mg orally once daily; GOL (50 or 100 mg); golimumab 50 or 100 mg SC every 4 weeks; MTX = methotrexate; RTX (500 or 1,000 mg) = rituximab 500 or 1,000 mg IV 2 courses (15 days apart) every 24 weeks; TCZ (4 or 8 mg/kg); tocilizumab 4 or 8 mg/kg IV every 4 weeks, TOF (5 or 10 mg) = tofacitinib 5 or 10 mg orally twice daily.

##### *Supplementary figure 3 SUCRA serious infection at 24-26 weeks*

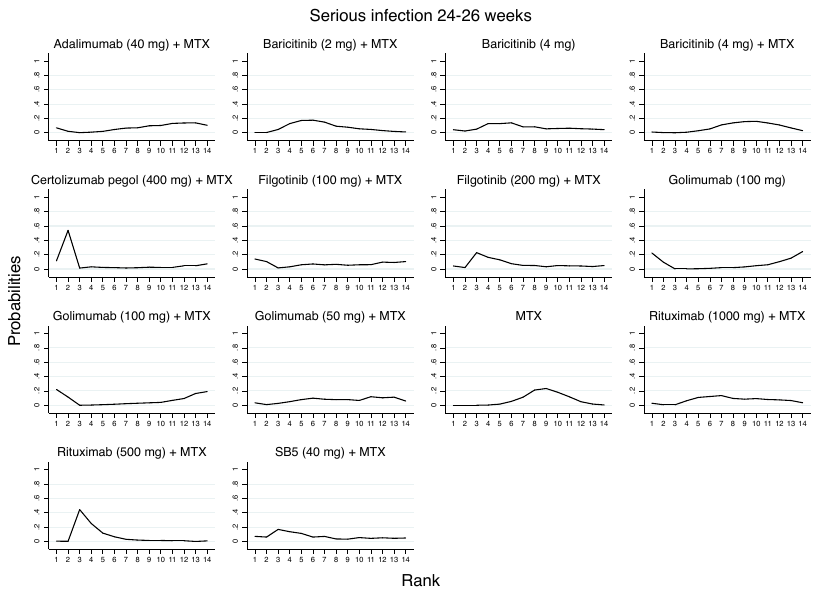

Adalimumab (40 mg) = adalimumab 40 mg SC injection every other week Baricitinib (2 or 4 mg) = baricitinib 2 or 4 mg orally once daily; Certolizumab pegol (400 mg) = certolizumab pegol 400 mg SC every 4 weeks; Filgotinib (200 mg) = filgotinib 200 mg orally once daily; Golimumab (50 or 100 mg) = golimumab 50 or 100 mg SC every 4 weeks; MTX = methotrexate; Rituximab (500 or 1,000 mg) = rituximab 500 or 1,000 mg IV 2 courses (15 days apart) every 24 weeks; SB5 (40 mg) = adalimumab biosimilar SB5 40 mg every other week SC.

##### *Supplementary figure 4 SUCRA serious infection at 48-52 weeks*

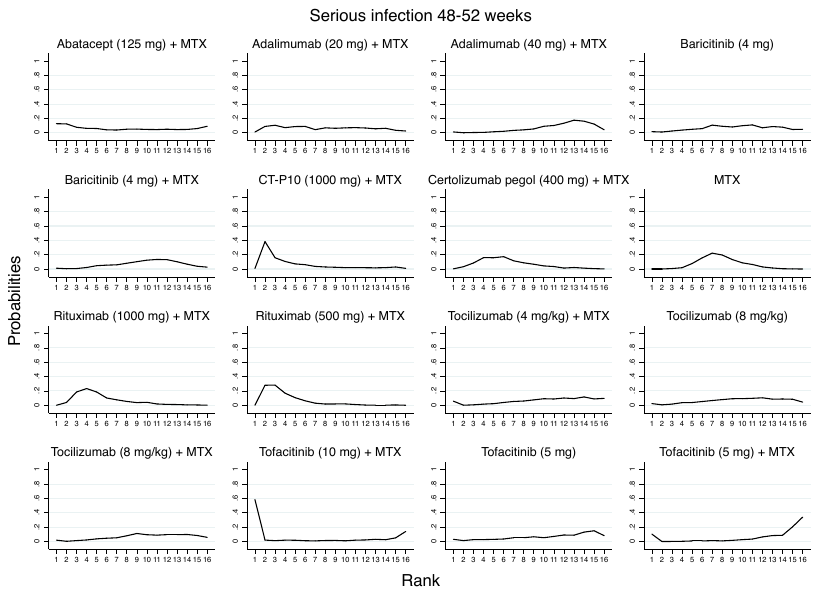

Abatacept (125 mg) = abatacept 125 mg subcutaneously injection (SC) once weekly; Adalimumab (20 or 40 mg) = adalimumab 20 or 40 mg SC injection every other week; Baricitinib (4 mg) = baricitinib 4 mg orally once daily; Certolizumab pegol (400 mg) = certolizumab pegol 400 mg SC every 4 weeks; CT-P10 (1,000 mg) = rituximab biosimilar CT-P10 1,000 mg; MTX = methotrexate; Rituximab (500 or 1,000 mg) = rituximab 500 or 1,000 mg IV 2 courses (15 days apart) every 24 weeks; Tocilizumab (4 or 8 mg/kg) = tocilizumab 4 or 8 mg/kg IV every 4 weeks; Tofacitinib (5 or 10 mg) = tofacitinib 5 or 10 mg orally twice daily.

##### *Supplementary figure 5 SUCRA tuberculosis at 24-26 weeks*

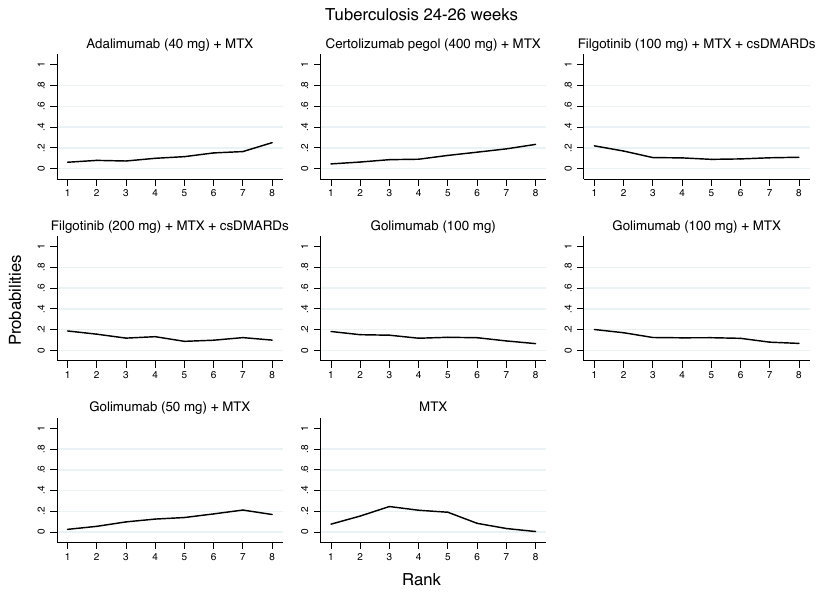

Adalimumab (40 mg) = adalimumab 40 mg SC injection every other week; Certolizumab pegol (400 mg) = certolizumab pegol 400 mg SC every 4 weeks; csDMARDs = sulfasalazine and hydroxychloroquine; Filgotinib (100 or 200 mg) = filgotinib 100 or 200 mg orally once daily; Golimumab (50 or 100 mg) = golimumab 50 or 100 mg SC every 4 weeks; MTX = methotrexate.

##### *Supplementary figure 6 SUCRA tuberculosis at 48-52 weeks*

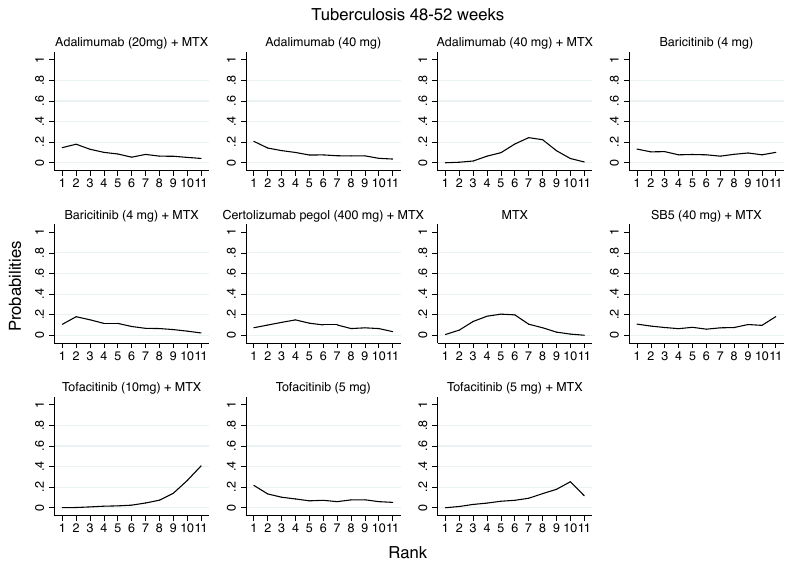

Adalimumab (20 or 40 mg) = adalimumab 20 or 40 mg SC injection every other week; Baricitinib (4 mg) = baricitinib 4 mg orally once daily;

Certolizumab pegol (400 mg) = certolizumab pegol 400 mg SC every 4 weeks; MTX = methotrexate; SB5 = adalimumab biosimilar SB5 40 mg every other week SC; Tofacitinib (5 or 10 mg) = tofacitinib 5 or 10 mg orally twice daily.

##### *Supplementary figure 7 SUCRA herpes zoster at 24-26 weeks*

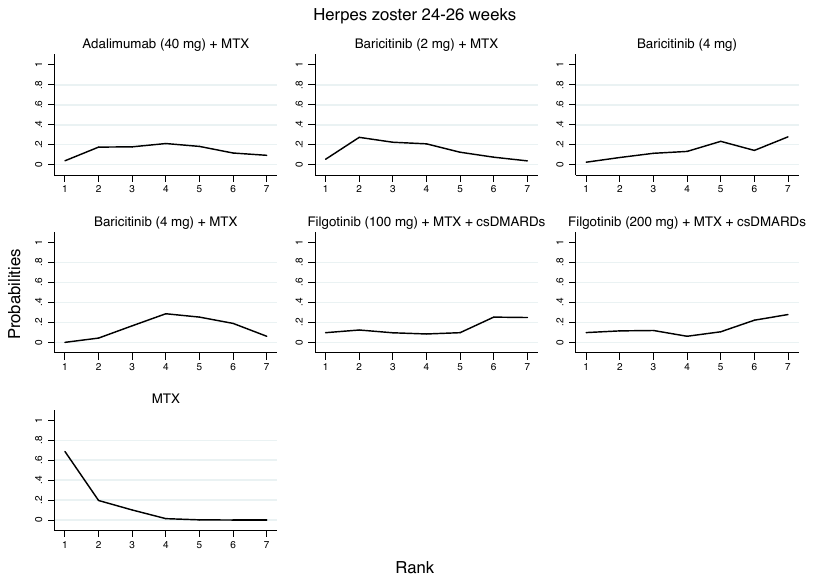

Adalimumab (40 mg) = adalimumab 40 mg SC injection every other week; Baricitinib (2 or 4 mg) = baricitinib 2 or 4 mg orally once daily; Filgotinib (100 or 200 mg) = filgotinib 100 or 200 mg orally once daily; MTX = methotrexate.

##### *Supplementary figure 8 SUCRA herpes zoster at 48-52 weeks*

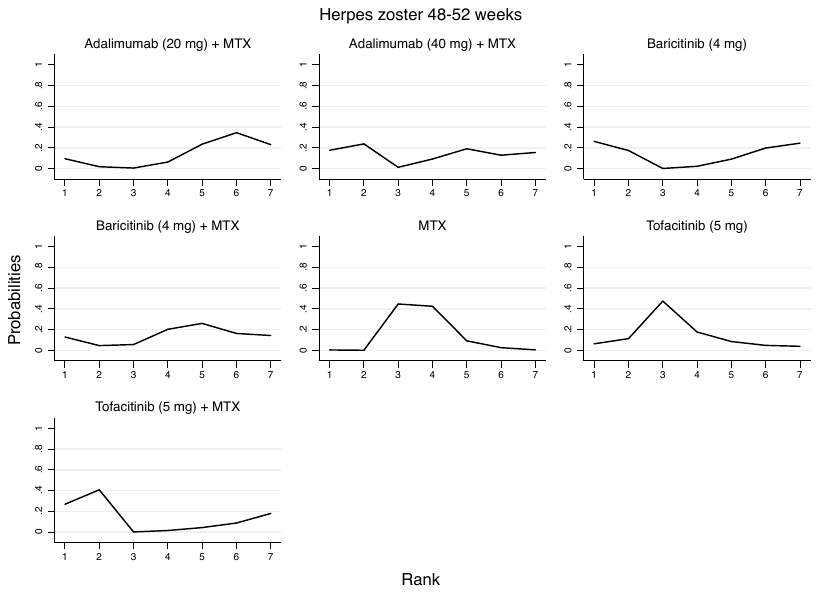

Adalimumab (20 or 40 mg) = adalimumab 20 or 40 mg SC injection every other week; Baricitinib (4 mg) = baricitinib 4 mg orally once daily; MTX = methotrexate; Tofacitinib (5 mg) = tofacitinib 5 mg orally twice daily.

##### *Supplementary figure 9 SUCRA cancer at 24-26 weeks*

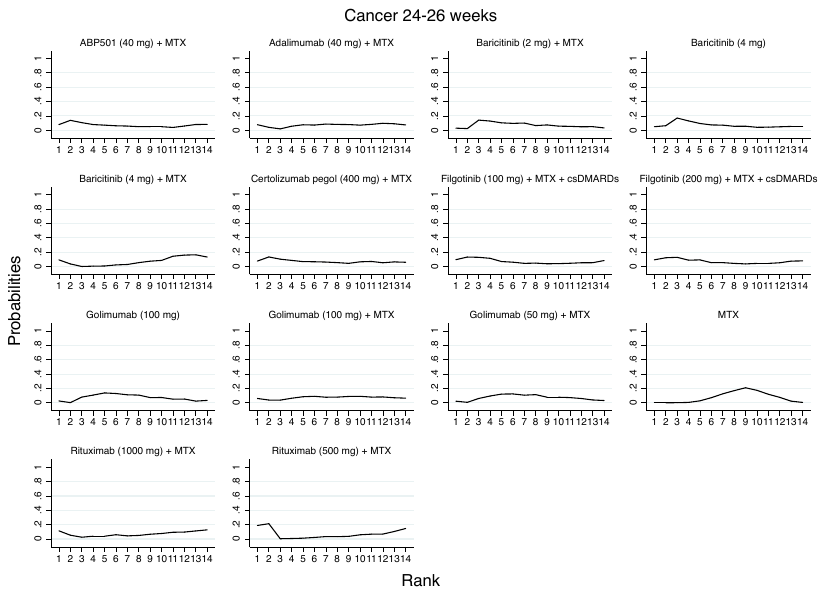

ABP501 (40 mg) = adalimumab biosimilar ABP50 40 mg every 2 weeks; Adalimumab (40 mg) = adalimumab 40 mg SC injection every other week; Baricitinib (2 or 4 mg) = baricitinib 2 or 4 mg orally once daily; Certolizumab pegol (400 mg) = certolizumab pegol 400 mg SC every 4 weeks; Filgotinib (100 or 200 mg) = filgotinib 100 or 200 mg orally once daily; Golimumab (50 or 100 mg) = golimumab 50 or 100 mg SC every 4 weeks; MTX = methotrexate; Rituximab (500 or 1,000 mg) = rituximab 500 or 1,000 mg IV 2 courses (15 days apart) every 24 weeks.

##### *Supplementary figure 10 SUCRA cancer at 48-52 weeks*

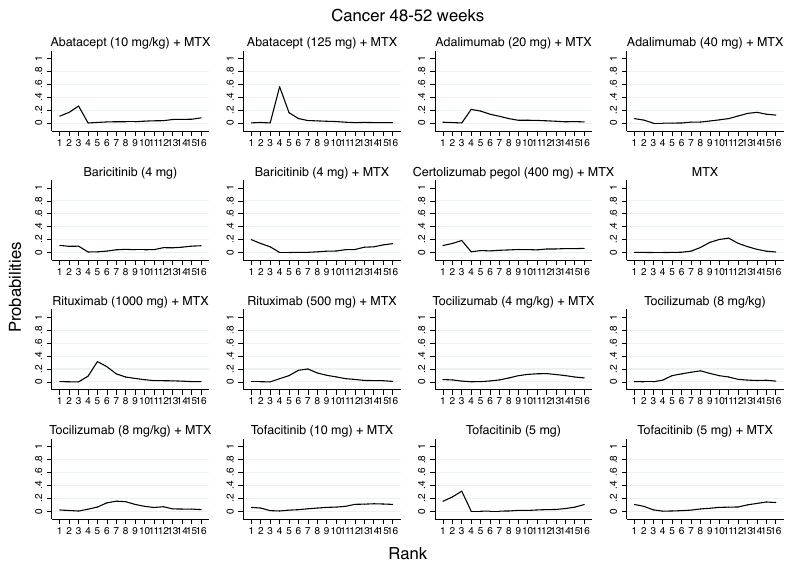

Abatacept (10 mg/kg) = abatacept 10 mg/kg intravenous infusion (IV) every 4 weeks; Abatacept (125 mg) = abatacept 125 mg subcutaneously injection (SC) once weekly; Adalimumab (20 or 40 mg) = adalimumab 20 or 40 mg SC injection every other week; Baricitinib (4 mg) = baricitinib 4 mg orally once daily; Certolizumab pegol (400 mg) = certolizumab pegol 400 mg SC every 4 weeks; MTX = methotrexate; Rituximab (500 or 1,000 mg) = rituximab 500 or 1,000 mg IV 2 courses (15 days apart) every 24 weeks; Tocilizumab (4 or 8 mg/kg) = tocilizumab 4 or 8 mg/kg IV every 4 weeks; Tofacitinib (5 or 10 mg) = tofacitinib 5 or 10 mg orally twice daily.

*Supplementary figure 11 SUCRA cardiovascular events at 24-26 weeks*

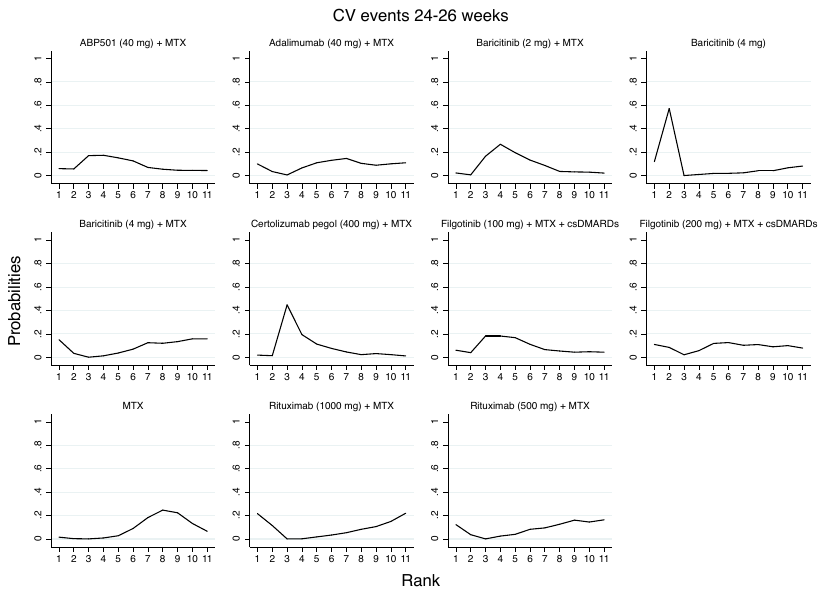

ABP501 (40 mg) = adalimumab biosimilar ABP501 40 mg every 2 weeks; Adalimumab (40 mg) = adalimumab 40 mg SC injection every other week; Baricitinib (2 or 4 mg) = baricitinib 2 or 4 mg orally once daily; Certolizumab pegol (400 mg) = certolizumab pegol 400 mg SC every 4 weeks; MTX = methotrexate; Filgotinib (100 or 200 mg) = filgotinib 100 or 200 mg orally once daily; Rituximab (500 or 1,000 mg) = rituximab 500 or 1,000 mg IV 2 courses (15 days apart) every 24 weeks.

.

##### *Supplementary figure 12 SUCRA cardiovascular events at 48-52 weeks*

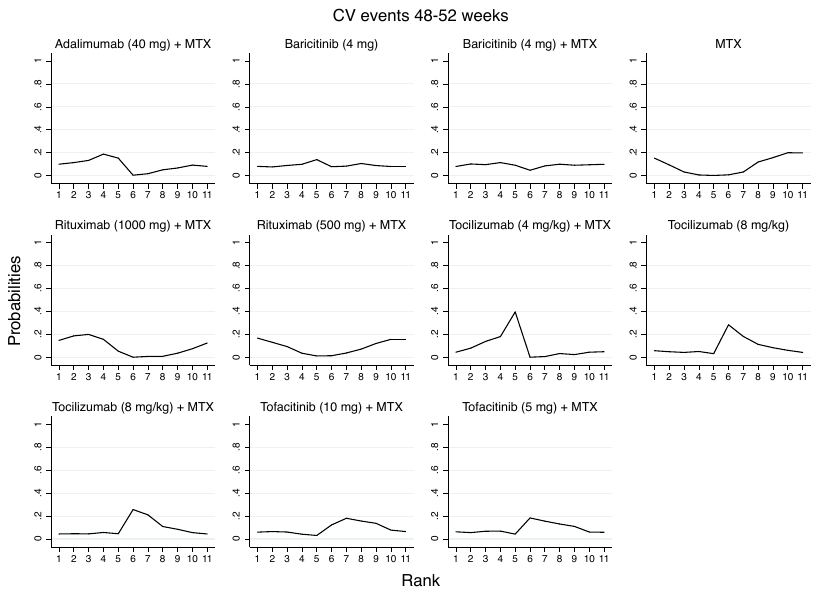

Adalimumab (40 mg) = adalimumab 40 mg SC injection every other week; Baricitinib (4 mg) = baricitinib 4 mg orally once daily; MTX = methotrexate; Rituximab (500 or 1,000 mg) = rituximab 500 or 1,000 mg IV 2 courses (15 days apart) every 24 weeks; Tocilizumab (4 or 8 mg/kg) = tocilizumab 4 or 8 mg/kg IV every 4 weeks; Tofacitinib (5 or 10 mg) = tofacitinib 5 or 10 mg orally twice daily.
